# FlahyRecovery is an AI & gene expression based immune profiling biomarker for decision support in precision oncology

**DOI:** 10.64898/2026.11.12.26367994

**Authors:** Jagjit Singh, Pragya Singh, Christopher E Mason, Sonia

**Affiliations:** Flahy, USA & India; Department of Physiology and Biophysics, Weill Cornell Medicine, New York, NY, USA

**Keywords:** Artificial Intelligence (AI), Tumor Microenvironment, Next-Generation Sequencing, Subtyping, Algorithm, Metrics, Precision Medicine

## Abstract

**Background:** Accurate characterization of the tumor microenvironment (TME) is crucial for inferring tumor biology, subsequent tumor behavior, and predicting therapeutic response. Current characterization methods are inadequate in capturing the complexity of immune signaling networks, hindering robust clinical decision making. To address this, we have developed FlahyRecovery – an artificial intelligence (AI) and gene expression based immune profiling biomarker**, implemented via a supervised classification machine learning model,** that integrates response prediction and prognostic insights to personalize treatment design, particularly immunotherapy.

**Methods:** Gene expression data of 7424 tumor specimens across 14 cancers was annotated specimen wise into three immune subtypes: Elevated Immunity (Hot) with high cytotoxic T-cell infiltration, Suppressed Immunity (Cold) marked by immune exclusion and stromal dominance, and Moderate Immunity (Intermediate) exhibiting mixed features. Following annotation validation, an AI model was trained and tested on the annotated data to classify new tumor specimens into the three immune subtypes – Elevated Immunity (Hot), Suppressed Immunity (Cold) and Moderate Immunity (Intermediate) based on the expression of a panel of 1275 genes involved in immune cell signaling and immune signal transduction.

**Results:** In a multi-tumor validation cohort (N=1337), the subtyping model showed a sensitivity of 79%, a specificity of 89%, and a one-vs-rest AUC of 0.93. Elevated Immunity tumors correlated with prolonged survival and immunotherapy response, while Suppressed Immunity tumors correlated with treatment resistance and need for combination therapy.

**Conclusion:** Artificial intelligence can be effectively used to process tumor biology digitized via next-generation sequencing, to accurately mark the TME from an immunological perspective. Its clinical utility mechanisms andying patients who can benefit from immunotherapies, identifying treatment resistance mechanisms, and informing combination treatment regimes.

## INTRODUCTION

The tumor microenvironment (TME) is the ecosystem surrounding the tumor. This ecosystem is a heterogeneous collection of infiltrating resident and host cells [1]. The main components of the TME are cancer cells, immune cells, stromal cells, blood vessels, and the extracellular matrix. It has now been established that the tumor microenvironment plays an active role in cancer progression instead of being a silent bystander [2]. Furthermore, studies have proven that the cell-type composition of the TME differs across individuals, as the tumor cells have distinct cell type populations, and the proportions of these cell type populations differ across individuals resulting in differing states of the TME. Over the past decade, numerous studies and recent reviews have established that the cellular and molecular composition of the tumor microenvironment (TME), including immune cells, stromal elements, vasculature, and extracellular matrix, plays a decisive role in tumor progression, metastatic spread, and response to systemic therapies, particularly immune checkpoint blockade [1–3]. TMEs can be classified along an “immune hot” to “immune cold” spectrum, where cytotoxic T cell– inflamed, interferon γ–rich, antigen presentation–competent microenvironments tend to be associated with improved survival and higher likelihood of benefit from PD 1/PD L1 or CTLA 4 inhibition, whereas immune excluded or immunosuppressive TMEs with abundant regulatory T cells, myeloid derived suppressor cells, and cancer associated fibroblasts are enriched among non responders [4–6]. These observations have been reproduced across multiple solid tumors, including breast cancer, lung cancer, melanoma, gastric cancer, hepatocellular carcinoma, pancreatic cancer, and cervical cancer, supporting the general principle that immune based stratification into immune high, immune intermediate, and immune low states captures clinically meaningful heterogeneity in prognosis and therapeutic benefit [7–9]. Therefore, there is a clinical need to capture the TME and leverage that for precise clinical management of the tumor.

Since 2010, clinicians and researchers have pointed towards the existence of tumor subtypes, with microarray & next-generation sequencing enabling researchers and clinicians to digitize this biology at a high resolution to refine the subtypes for improved clinical utility. The increased accessibility of next-generation sequencing has enabled the large-scale sequencing of the tumor transcriptome profile. Since 2010, numerous studies have suggested the existence of tumor subtypes, and these subtypes have been refined over time, particularly in relation to immune cell signaling and signal transduction. Multiple studies have validated the presence of immune subtypes in different solid tumors [10–14]. Importantly, multiple clinical and analytical retrospective studies have found that patients exhibiting elevated expression of specific immune-associated pathways show better prognoses, overall survival rates, and higher responses to immunotherapy, compared to those in whom these pathways are suppressed (**Table 1** and **Table 2**). For each cancer, three immune subtypes are defined: EI (Elevated Immunity) or Immune-High (“hot”), MI (Moderate Immunity) or Immune-Moderate, and SI (Suppressed Immunity) or Immune-Low (“cold”). Tumors classified as EI (e.g., BC-ImH, SubA, ImH, IS3, Subtype2, C1 for glioma, C2 for pancreatic and cervical cancer, C3 for prostate cancer, and IM for triple negative breast cancer in **Table 1** and **Table 2**) exhibit elevated immune cell signaling and signal transduction in the TME. These “hot” tumors are associated with better prognosis and higher five-year OS rates across most cancer types, with exceptions observed in renal cell carcinoma, prostate cancer, and glioma, where Immune-High subtypes do not correspond to the highest survival (**Figure 1** and **Figure 2**). In contrast, SI or Immune-Low (“cold”) subtypes (e.g., BC-ImL, SubB, ImL, IS1, Subtype1, C3 for glioma, C1 for pancreatic and cervical cancer, C1 for prostate cancer, and BLIS for triple negative breast cancer) are characterised by suppressed immune cell signaling, low immune infiltration, and a less active TME, which is associated with poorer prognosis and lower five-year OS rates (**Figure 1** and **Figure 2**).

**Table 1:** Five-year overall survival (OS) rates across immune subtypes—Immunity-High (Im-H), Immunity-Moderate (Im-M), and Immunity-Low (Im-L) for various cancer types.

| Cancer Types | Treatment | Subtypes | Overall Survival at 5 Years | Number of Cancer Specimens (n) | Reference |
|---|---|---|---|---|---|
| Breast Cancer | Hormone and Chemotherapy | BC-ImH<br>BC-ImM<br>BC-ImL | BC-ImH:~84%<br>BC-ImM:~76%<br>BC-ImL:~80% | BC-ImH(n=411)<br>BC-ImM(n=1146)<br>BC-ImL(n=535) | [15] |
| Non-Small Cell Lung Cancer | Chemotherapy, Radiation therapy | SubA<br>SubB | LUAD:<br>SubA:~50%<br>SubB:~30%<br>LUSC:<br>SubA:~58%<br>SubB:~48% | LUAD:<br>SubA(n=314)<br>SubB(n=178)<br>LUSC:<br>SubA(n=256)<br>SubB(n=239) | [16] |
| Head and Neck Squamous Cell Cancer | Chemotherapy, Radiation therapy | Im-H<br>Im-M<br>Im-L | Im-H:~60%<br>Im-M:~43%<br>Im-L:~40% | Im-H(n=161)<br>Im-M(n=234)<br>Im-L(n=127) | [13] |
| Gastric Cancer | Chemotherapy, Targeted therapy, Radiation therapy | IS1<br>IS2<br>IS3 | IS1:~41%<br>IS2:~45%<br>IS3:~64% | IS1(n=252)<br>IS2(n=102)<br>IS3(n=139) | [17] |
| Cutaneous Melanoma | Chemotherapy, Targeted therapy | Im-H<br>Im-M<br>Im-L | Im-H:~70%<br>Im-M:~60%<br>Im-L:~38% | Im-H(n=44)<br>Im-M (n=135)<br>Im-L(n=275) | [11] |
| Renal Cell Carcinoma | Chemotherapy | Subtype1<br>Subtype2 | Subtype1:~75%<br>Subtype2:~62% | Subtype1(n=301)<br>Subtype2(n=229) | [21] |
| Hepatocellular Carcinoma | - | Im-H<br>Im-M<br>Im-L | Im-H+M:~76%<br>Im-L:~52% | Im-H+M(n=343)<br>Im-L(n=231) | [18] |
| Diffuse Glioma | Chemotherapy, Radiation therapy | C1<br>C2<br>C3 | C1:~8%<br>C2:~42%<br>C3:~62% | C1(n=97)<br>C2(n=397)<br>C3(n=178) | [22] |
| Pancreatic Cancer | Chemotherapy | C1<br>C2 | C1:~25%<br>C2:~38% | C1(n=97)<br>C2(n=82) | [19] |
| Cervical Cancer | Chemotherapy, Radiation therapy | C1<br>C2<br>C3 | C1:~45%<br>C2:~78%<br>C3:~75% | C1(n=96)<br>C2(n=101)<br>C3(n=94) | [20] |
| Prostate Cancer | Chemotherapy | C1<br>C2<br>C3 | C1:~88%<br>C2:~78%<br>C3:~62% | C1(n=133)<br>C2(n=92)<br>C3(n=88) | [56] |

Abbreviation IS: Immune Subtype; LUAD: Lung Adenocarcinoma; LUSC: Lung Squamous Cell Carcinoma.

**Table 2:** Objective Response Rates (ORRs) across immune subtypes in Triple-Negative Breast Cancer (TNBC), Renal Cell Carcinoma (RCC), and Cutaneous Melanoma.

| **Cancer Types** | **Treatment** | **Subtypes** | **Objective Response Rates** | **Number of Cancer Specimens (n)** | **Reference** |
|---|---|---|---|---|---|
| Triple Negative Breast Cancer | Chemotherapy,<br>Immunotherapy | IM<br>BLIS | IM:<br>Arm C: 43.5%<br>BLIS:<br>Arm D:20.0%<br>Arm E:28.3% | IM(n=46)<br>BLIS<br>Arm D(n=10)<br>Arm E(n=46) | [57] |
| Renal Cell Carcinoma | Atezolizumab | Subtype1<br>Subtype2 | Subtype1:16%<br>Subtype2:29% | (n=348) | [21] |
| Cutaneous Melanoma | Immune Checkpoint Inhibitor Therapy | Im-H<br>Im-M<br>Im-L | ICI-R:<br>Im-H:44.4%<br>Im-M:31.3%<br>Im-L:18.6%<br>ICI-M:<br>Im-H:42.9%<br>Im-M:57.9%<br>Im-L:8% | ICI-R<br>Im-H(n=67)<br>Im-M(n=45)<br>Im-L(n=70)<br>ICI-M(n=65) | [11] |

As summarised in Figure 1A, tumors with high immune activity are generally associated with higher five-year overall survival (OS) across the majority of cancer types examined, whereas Immune-Low tumors consistently show poorer survival outcomes. Although the magnitude of this difference varies across cancers, Im-L tumors consistently exhibit poorer survival, highlighting cancer-specific relationships between immune activity and clinical outcome. Exceptions to this trend are observed in renal cell carcinoma, prostate cancer, and glioma, where Immune-High subtypes do not correspond to the highest survival rates (Figure 2A). This overall trend is further supported by the pooled survival distributions presented in Figure 1B, which show a higher median five-year OS for Immune-High tumors compared with Immune-Moderate and Immune-Low subtypes, highlighting the prognostic value of immune-based tumor stratification across studies.

**Figure 1C** shows the estimated survival curves from previously published Kaplan–Meier analyses across different cancer types. This summary curve shows that the Im-H subtype has a higher mean survival compared with Im-L across the compiled datasets. For example, Yao et al. (Figure 3A) reported Kaplan–Meier curves of overall survival using the METABRIC, GSE24450 data, which display that the Im-H subtype exhibits better overall survival than Im-L in breast cancer [15]. Li et al. (Figures 1D and 2D, TCGA LUAD and LUSC) showed that subtype A(Im-H) demonstrates significantly better survival than subtype B(Im-L) [16]. Song et al. (TCGA HNSCC) showed in Figure 3 the Kaplan–Meier survival curve that Immunity-H(Im-H) patients have better overall survival than Immunity-L(Im-L) [13]. Chen, Yan et al. (TCGA-GC and GEO cohorts) reported (Figure 3H, I) that IS3(Im-H) exhibits superior overall survival relative to IS1(Im-L) and IS2(Im-M) [17]. Qian Liu et al. (TCGA melanoma) showed in Figure 3A a Kaplan-Meier curve that Im-H has a better prognosis than Im-L [11]. Zhuang et al. (TCGA and LIRI_JP hepatocellular carcinoma) in Figures 3I and J demonstrated that Immunity-H and Immunity-M (Im-H and Im-M) had better survival than Immunity-L(Im-L) [18]. Du et al. Figures 3 F and G (TCGA and GSE62452 pancreatic cancer) reported that C2(Im-H) has longer survival than C1(Im-L) [19]. Deng et al. (TCGA-CESC) showed that the C2(Im-H) subtype has the best survival relative to C1(Im-L) in Figure (1A) [20].

**Figure 2C** presents the average survival curve calculated by aggregating survival probabilities extracted from previously published studies. This summary curve shows that the Im-L subtype has a higher mean survival compared with Im-H across the compiled datasets. The supporting studies include Wang et al. (six combined ccRCC datasets GSE15641, GSE36895, GSE40435, GSE46699, GSE53757, and TCGA) showing that subtype 1 (Im-L) has better overall survival than subtype 2 (Im-H) in Figure(1C) [21]. Zhou et al. (Figure 1 E and F TCGA and GSE16011 glioma) reported that subtype C3 (Im-L) has longer median survival than subtype C1 (Im-H) [22]. Song et al. (Figure 7A) showing that Cluster III (Im-L) has better overall survival than Clusters I (Im-H) and II (Im-M) [13]. These published OS Kaplan–Meier curves form the basis for the averaged survival trends presented in **Figures 1C and 2C**. The average survival curve is calculated as the arithmetic mean of the approximate survival probabilities across all cancer types. Each original study used multiple datasets and reported Kaplan–Meier curves of overall survival analysis with censor marks at patient-level event times. **Figures 1C and 2C** are based on summary-level survival probabilities rather than patient-level time-to-event data, censoring times cannot be displayed, and the curve shows regular steps corresponding to the extracted time intervals.

Furthermore, the studies have reported that the EI subtype demonstrates the highest objective response rate (ORR), followed by MI and SI subtypes (EI > MI > SI) in triple negative breast cancer (TNBC), renal cell carcinoma, and cutaneous melanoma, indicating a positive association between immune activity and therapeutic response (**Table 2**). **Figure 3** illustrates Objective Response Rate (ORR) by immune subtype in TNBC, renal cell carcinoma, and cutaneous melanoma. The elevated immunity tumor has a significantly better prognosis under immunotherapy than the suppressed immunity and the moderate immunity tumor for the 14 cancer types that FlahyRecovery currently covers.

The Elevated Immunity tumor has the best overall survival regardless of treatment for Breast Cancer, Non-Small Cell Lung Cancer, Gastric Cancer, Head & Neck Cancer, Skin Cancer, Colon Cancer, Esophageal Cancer, Bladder Cancer, Liver Cancer, Pancreatic Cancer, and Cervical Cancer versus tumors of Suppressed Immunity (SI) and Moderate Immunity (MI) subtypes (**Figure 1**). The Suppressed Immunity tumor has the best overall survival for Kidney Cancer, Brain Cancer, and Prostate Cancer (**Figure 2**). However, post-immunotherapy-treatment of elevated immunity tumors, EI shows better prognosis than SI irrespective of the cancer type. FlahyRecovery builds on this work, wherein we process the gene expression of a panel of 1275 genes involved in the immune cell signaling and signal transduction, and subtype the patient’s tumor into Elevated Immunity, Moderate Immunity, and Suppressed Immunity with high accuracy and precision.

To this effect we have developed a subtyping model. The subtyping model comprises of a trained Random Forest (RF) machine-learning algorithm, a robust algorithm widely employed in the biomedical field. Originally conceptualized in 1995, RF began gaining traction in biomedical research during the early 2000s, as demonstrated by multiple studies [23–27]. Over the past two decades, the RF model’s architecture and training mechanisms have been refined to improve specificity and sensitivity. During the model training, the subtyping model constructs decision trees, each based on a random subset of the 1275 genes. These individual trees act as estimators of the immune subtype. The FlahyRecovery subtyping model then processes the decisions of these trees and assigns a final subtype according to the majority vote across all estimators/trees. Randomly sub-setting features (i.e., genes) reduces correlation among the individual decision trees, creating a more diverse ensemble and minimizing the risk of overfitting to noise or spurious signals. Furthermore, this approach prevents strong predictors from overshadowing other potentially informative genes, thereby capturing subtle patterns and interactions in the data. As a result, the model is significantly more robust when applied to previously unseen data.

Several immune-profiling and subtyping strategies have been proposed in recent years, each capturing different aspects of the tumor immune landscape. Pan-cancer immune state classifiers, such as the six immune subtypes (C1–C6) defined by Thorsson et al. through analysis of the TCGA Pan-Cancer Atlas, cluster tumors using curated immune and stromal gene-set scores rather than raw transcript levels, enabling broad categorization of immune phenotypes across thousands of genes [8]. Similarly, cell-type deconvolution algorithms such as CIBERSORT estimate relative fractions of 22 leukocyte subsets using ν-support vector regression applied to the 547-gene LM22 matrix and are widely used to study immune infiltration and correlate individual cell types with prognosis or therapy response [28]. A third class of approaches consists of cancer-specific immune gene-set clustering studies, where panels ranging from a few dozen to a few hundred immune-related genes are subjected to non-negative matrix factorization or consensus clustering to derive 2–4 immune subtypes optimized for a single tumor type [29–33]. These methods have yielded valuable insights, but most rely either on narrow fixed gene panels, or are limited to one cancer context, or classify broad immune states without providing a unified cross-cancer framework [34]. In contrast, FlahyRecovery leverages a larger curated panel of 1,275 immune-signaling genes and develops a unified three-state subtype classifier applicable across 14 solid tumor types using a machine-learning–based framework. Unlike prior methods optimized for cell fractions or broad immune states, FlahyRecovery produces clinically interpretable EI/MI/SI subtypes that directly correlate with immunotherapy response, treatment resistance, and potential need for combination therapy, thereby providing actionable guidance for precision oncology.

In addition to CIBERSORT and TIMER, several other transcriptome-based deconvolution and gene signature methods have been developed to quantify the immune and stromal composition of the TME from bulk RNA seq data. MCP counter infers the absolute abundance of multiple immune and stromal cell populations using carefully curated transcriptomic markers and has been shown to recover survival associated microenvironment patterns in lung, colorectal, and breast cancers [35]. xCell extends signature based approaches to 64 immune and stromal cell types using a spillover compensation framework and in turn improving robustness across tissues and experimental platforms [36]. EPIC (Estimating the Proportion of Immune and Cancer cells) enumerates cancer and immune cell types in solid tumors by fitting bulk expression profiles to reference signatures derived from RNA seq and microarray data [37]. quanTIseq uses a RNA seq derived signature matrix and deconvolution pipeline to quantify ten major immune cell types and to characterize the “tumor immune contexture” in large clinical cohorts, with applications to pharmacological modulation of the TME and immunotherapy response prediction [38]. Together with comparative evaluations of these tools, these studies underscore the maturity of transcriptome-based strategies for understanding of the TME and highlight that multiple, partly complementary frameworks exist for quantifying immune infiltration from bulk data [39].

Beyond cell fraction estimation, several transcriptomic frameworks have been proposed specifically as biomarkers of immunotherapy response that are closely related to the clinical objectives of FlahyRecovery. Jiang et al. developed TIDE (Tumor Immune Dysfunction and Exclusion), which models two dominant mechanisms of immune escape T cell dysfunction in highly infiltrated tumors and T cell exclusion in poorly infiltrated tumors and showed that TIDE scores predict response to anti PD 1 and anti CTLA 4 therapy more accurately than PD L1 expression or tumor mutational burden in multiple melanoma cohorts [6]. Bagaev et al. performed an integrative pan cancer transcriptomic analysis of more than 10,000 tumors and defined four conserved TME subtypes that combine malignant and microenvironmental signals. These subtypes act as generalized biomarkers of response to PD 1/PD L1 blockade across 20 cancer types [9]. Recent pan cancer studies on common molecular components of the human TME further reinforce that immune activation, stromal remodeling, and immunosuppressive circuits represent recurrent axes of variation that can be leveraged for prognostic and predictive modeling across tumor types [7]. The present work builds on this evolving literature by integrating a broad immune signaling gene panel with a supervised, pan cancer classifier that yields standardized EI/MI/SI subtypes, thereby unifying prior evidence on immune hot/immune cold phenotypes, and TME based immunotherapy biomarkers into a single clinically interpretable framework.

## METHODS

### Data Collection

To train and test the FlahyRecovery subtyping model we utilized an annotated gene expression dataset of 7424 specimens (**Figure 4**). The unannotated gene expression data was obtained from the United States National Institute of Health Cancer Database. Along with the gene expression data, we retrieved Masked Somatic Mutation data as well as gene level Copy Number Alteration data of the same patients. The total gene expression data was a culmination of 14 different datasets across the following cancers: Breast Cancer, Non-Small Cell Lung Cancer, Head and Neck Cancer, Gastric Cancer, Skin Cancer, Colon Cancer, Kidney Cancer, Bladder Cancer, Esophageal Cancer, Pancreatic Cancer, Prostate Cancer, Brain Cancer, Cervical Cancer, and Liver Cancer (**Table 3**). Each expression dataset was annotated (immune-subtyped) individually, and then all 14 cancer datasets were combined along with their respective annotated labels. Furthermore, the tumor immune-cell infiltration estimates of all the patients which we had in our 14 datasets were downloaded from the Tumor Immune Estimation Resource (TIMER) database and comparison was made among the 3 immune subtypes of tumors.

**Table 3:** Summary of TCGA Cancer Cohorts Used in This Study.

| **Cancer Type** | **TCGA project IDs** | **Number of Patients** |
|---|---|---|
| Breast Cancer | TCGA-BRCA | 1230 |
| Head and Neck Cancer | TCGA-HNSC | 522 |
| Gastric Cancer | TCGA-STAD | 412 |
| Melanoma | TCGA-SKCM | 472 |
| Colon Cancer | TCGA-COAD | 482 |
| Kidney Cancer | TCGA-KIRP,<br>TCGA-KIRC | 814 |
| Bladder Cancer | TCGA-BLCA | 412 |
| Esophageal Cancer | TCGA-ESCA | 185 |
| Prostate Cancer | TCGA-PRAD | 502 |
| Pancreatic Cancer | TCGA-PAAD | 179 |
| Glioma | TCGA-GBM | 158 |
| Cervical cancer | TCGA-CESC | 306 |
| Liver Cancer | TCGA-LIHC | 371 |
| Non-Small Cell Lung Cancer | TCGA: LUAD,<br>TCGA: LUSC,<br>TCGA: CPTAC3 | 1379 |

### Data Annotation

Each of the 7,424 TCGA specimens was assigned a ground-truth immune subtype label— Elevated Immunity (EI), Moderate Immunity (MI), or Suppressed Immunity (SI)—using a standardized annotation pipeline. The below procedure described in two parts was performed independently for each of the 14 cancer types.

#### Pathway Activity Quantification Using GSVA

FPKM-normalized TCGA RNA-seq gene expression matrices were used as input. FPKM values were selected because they represent the standardized, publicly released TCGA quantifications, ensuring reproducibility and consistency across datasets. As GSVA relies on relative expression patterns rather than raw count modelling, FPKM-normalized values are suitable for pathway-level enrichment analysis.

Activity scores were computed for 40 curated immune-related gene sets representing key immune signaling and functional pathways, including cytotoxic T-cell signatures, interferon-γ (IFN-γ) signaling, antigen presentation, chemokine signaling, stromal activation, and immune-exclusion pathways. Pathway activities were quantified using Gene Set Variational Analysis (GSVA). It is an unsupervised, non-parametric method that transforms gene-level expression data into pathway-level activity scores for each sample. Instead of focusing on differential expression of individual genes, GSVA evaluates the coordinated expression of predefined gene sets and computes a continuous enrichment score representing the relative activation state of each pathway across samples.

For a given cancer type, the GSVA output was of the form:

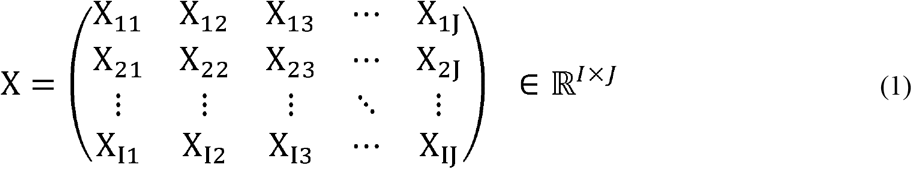

where X*_IJ_*denotes the GSVA enrichment score for sample I and gene set J.

#### Spectral Embedding, Clustering, and Annotation

Once the GSVA was done spectral embedding and spectral clustering were used to identify immune subtypes from the GSVA-derived pathway activity matrix. Spectral embedding is a nonlinear dimensionality-reduction technique that constructs a similarity graph between samples and uses the eigenvectors of the graph Laplacian to project the data into a lower-dimensional space. This approach captures the intrinsic manifold structure of the data and preserves complex, nonlinear relationships that may not be detectable using linear methods. This enables to reduce dimensionality of the data before clustering. The resulting embedded representation is given by:

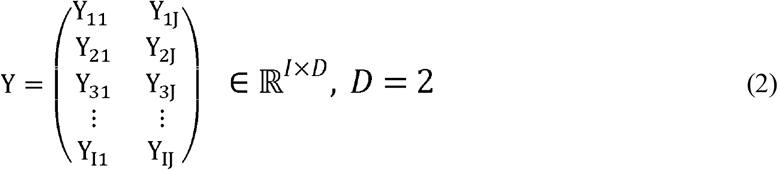

where Y_IK_ represents the K^th^ embedding coordinate of sample I where K = 1, D and D=2 denotes the number of embedding dimensions.

Spectral clustering was then applied to embedded representation, to enhance separability and enable detection of non-spherical or heterogeneous cluster structures. Each resulting cluster was mapped to Elevated (EI), Moderate (MI), or Suppressed Immunity (SI) based on predefined biological criteria: EI clusters showed high cytotoxic, IFN-γ, and APC signatures as samples in the EI cluster had the highest respective GSVA scores; The SI cluster exhibited low immune activation with high stromal/immune-exclusion signals as samples in the SI cluster had the lowest respective GSVA scores. Finally, the MI clusters showed intermediate or mixed scores, as samples in the MI cluster did not show any significant enrichment or suppression based on the GSVA scores. Following is the output from spectral clustering and cluster annotation step:

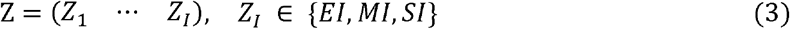

where Z_I_ denotes the immune subtype label assigned to sample I. This three-step process Eq (1) to Eq (3) was done individually for each of the 14 cancer types for annotating the specimens for model training.

The annotation relied solely on pathway-level immune activity scores derived from curated gene sets (e.g., cytotoxicity signatures, interferon signaling, antigen presentation, chemokine signaling, stromal activation) and not on immune cell infiltration data thereby reflecting functional immune pathway activation, not immune cell abundance estimates.

### Validation of Specimen Annotation with Immune cell infiltration data

The annotation of the specimens into the three subtypes – EI, MI and SI were further validated with the immune cell infiltration estimates available on the TIMER database of all the specimens that we included in our 14 datasets [40–42]. TIMER is a resource designed for the systematic analysis of immune cell infiltration across diverse cancer types. The tumor immune subtype-wise distribution plots were generated for the 7 major cell types – CD4+ T cells, Dendritic cells, CD8+ T cells, B cells, Neutrophils, Macrophages, and T-regulatory cells to validate whether the annotations are correct i.e. whether EI specimens have a higher level of immune cell (CD4+ T cells, Dendritic cells, CD8+ T cells, B cells, Neutrophils, and Macrophages) infiltration versus MI and SI specimens, and whether SI specimens have a lower level of immune cell (CD4+ T cells, Dendritic cells, CD8+ T cells, B cells, Neutrophils, and Macrophages) infiltration versus MI and EI specimens. TIMER infiltration values were used only after subtype labels had been assigned, as an external, orthogonal biological check. TIMER employs a constrained regression model integrating multiple non-overlapping reference profiles and is not derived from the pathway scores or gene signatures used for annotation. Thus, TIMER infiltration values serve as an independent validation layer: confirming that samples annotated as Elevated Immunity exhibit higher CD8+, CD4+, dendritic cell, and macrophage infiltration, while Suppressed Immunity samples show the lowest levels.

### Model Training

The combined database was split into training and validation, with the expression data of 6087 specimens used for training and the expression data of 1337 specimens used for testing. The training dataset was provided to the subtyping model which learned to classify specimens into EI, MI, or SI based on the expression of the FlahyRecovery panel genes. To optimize performance, a wide hyperparameter search grid was conducted and model training was done using 5-fold cross-validation. The search grid encompassed the key parameters of the subtyping model including the number of estimators (trees) in the forest, the training criterion and the depth of the tree. Cross-validation is a technique for estimating how well a model generalizes to unseen data by systematically splitting the training dataset into multiple parts (folds). This approach offers a more robust and less biased performance assessment compared to a single train/test split, as it helps prevent overfitting. In each iteration, the model was trained on four folds and validated on the remaining one, and the Compute Area Under the Receiver Operating Characteristic Curve for each validation was recorded, as each iteration contained different hyperparameter values. The model with the highest Compute Area Under the Receiver Operating Characteristic Curve was chosen.

## RESULTS

### Annotation validation through Immune-cell Infiltration data

**Figure 5** shows the immune cell infiltration patterns across combined cancer types. It can be observed that the EI tumor subtype shows the highest immune cell infiltration followed by MI and SI. **Supplementary Figures S1-S16** further show the cancer wise distribution patterns of different immune cells in the 3 tumor immune subtypes (EI, MI, and SI) which further provides a proof of validity of the immune subtype annotations. The Welch’s t-test and the Mann-Whitney U test were performed across specimens amongst the 3 tumor immune subtypes (EI-MI, EI-SI, and MI-SI) for different immune cells and the values are listed in **Supplementary Tables S2-S19**. Further we observed that majority of the cancers (BLCA, CESC, COAD, ESCA, GBM, KIRC, KIRP, LIHC, LUAD, PAAD, and STAD) showed higher T-regulatory cell infiltration in the SI subtype when compared to EI subtype (**Supplementary Figure S17**). T-regulatory cells (Tregs) are key mediators of immunosuppression, facilitating tumor immune evasion and progression through various mechanisms. Tregs inhibit the function of effector T cells (Helper CD4+ T cells and cytotoxic CD8+ T cells) [43,44]. Further, Tregs secrete cytokines such as TGF-β, IL-10, and IL-35, which directly suppress the activity and proliferation of effector T cells and natural killer (NK) cells within the TME [43,44]. These cytokines also inhibit the production of pro-inflammatory cytokines and cytolytic molecules by effector cells, further dampening the anti-tumor response. Tregs can also suppress the maturation and function of dendritic cells (DCs) through modulation of CTLA-4 molecules, thereby reducing ability of DCs to present tumor antigens and activate effector T cells [4,43]. Tregs also express high levels of CD25 (the IL-2 receptor α-chain), allowing them to consume IL-2, a cytokine essential for effector T cell survival and proliferation, thus interfering with the growth of other T cells [43]. Therefore, higher T-regulatory cell infiltration is suggestive of immune tolerance, tumor progression and poor prognosis and hence, makes the SI tumor subtype to respond poorly to immunotherapies and standard treatments [43, 45, 46].

### Model testing

The trained & fine-tuned subtyping model was then tested on 1337 distinct patient tumor specimens, and confusion matrices for each subtype were computed. To optimize FlahyRecovery’s performance, we conducted a grid search over the Random Forest parameter space. The final model selected via five-fold cross-validation was: **RandomForestClassifier(criterion=’log_loss’, max_depth=35, max_features=0.3, min_samples_split=6, n_estimators=953)**.

This configuration balances predictive accuracy with model stability.

To assess biological interpretability, we examined the top **N = 150 most important genes** identified by the trained model. Strikingly, the highest-ranking genes were overwhelmingly enriched for canonical immune pathways, validating the biological relevance of the learned decision boundaries. The top features clustered into coherent immune modules:

1. **Antigen presentation (MHC-I and MHC-II machinery):** *HLA-DRA, HLA-DRB1, HLA-DPB1, HLA-DMB, CD74, TAP1, HLA-B, B2M*, reflecting strong interferon signaling and antigen processing activity.
2. **T-cell receptor signaling and lineage markers:** *CD3D, CD3E, CD3G, CD247, CD2, CD4, CD5, LCK, LAT*, indicating T-cell infiltration and activation status.
3. **Chemokines and chemokine receptors (immune trafficking):** *CCR5, CCR2, CCR7, CXCR6, CXCL11, CCL22, CCL4*, supporting T-cell, dendritic cell, and macrophage recruitment biology.
4. **Innate sensing and inflammatory pathways:** *TLR8, NLRP3, MAVS, IRF1, IRF8, STAT1, TNFAIP3*, representing antiviral and NF-κB-mediated responses.
5. **Complement system components:** *C1QA, C1QB, C1QC, C1R, MASP1, SERPING1*, consistent with macrophage and stromal-associated innate immunity.
6. **Myeloid and antigen-presenting cell markers:** *CD14, CSF1R, CYBB, HCK, LILRB2, CD1C*, indicating monocyte/macrophage infiltration.
7. **Cytokine receptors and immune regulatory genes:** *IL12RB1, IL10RA, IL7R, IL2RB, IL4R, CSF2RB, EBI3*, reflecting differentiation and immunoregulatory signaling.
8. **Immune checkpoint and exhaustion markers:** *PDCD1LG2, TIGIT, ICOS, VSIR, CD86*, capturing immune suppression and T-cell dysfunction axes.

The aforementioned modules show that FlahyRecovery prioritizes biologically meaningful immune circuits, antigen presentation, effector T-cell activity, chemotaxis, innate sensing, and checkpoint signaling, which aligns with the expected molecular architecture of EI, MI, and SI phenotypes. This feature-importance analysis further supports the validity of the model’s learned decision rules and its mechanistic relevance to tumor–immune interactions.

The confusion matrices are shown in **Figure 6**. A confusion matrix compares a classification model’s predictions with the actual outcomes, showing the counts of true positives, false positives, false negatives, and true negatives. It is used for sensitivity and specificity calculations. Importantly the Area Under the Receiver Operating Characteristic Curve (AUC) in a one-vs-rest setting was also computed to assess performance on unseen (test) tumor specimens. A Receiver Operating Characteristic (ROC) curve is a plot of sensitivity (true positive rate) versus 1 – specificity (false positive rate) across all possible decision thresholds for a diagnostic test. The AUC, ranging from 0 to 1, provides a convenient summary of overall diagnostic accuracy, with 1 indicating a perfectly accurate test and 0 indicating a perfectly inaccurate one [47]. This metric is also compliant with the Standards for Reporting of Diagnostic Accuracy Studies [48]. In the one-vs-rest scheme, each class is treated in turn as “positive,” while all other classes are combined as “negative,” enabling a comprehensive evaluation of performance across multiple classes. The AUC value came out to be 0.93. The ROC curves, along with the respective histograms in one-vs-rest approach are plotted in **Figure 7**. They present the final model’s performance in differentiating the EI, MI, and SI subtypes, using both histogram plots (top row) of the predicted probabilities and one-vs-rest ROC curves (bottom row) for each class. Each histogram depicts the distribution of the model’s predicted probabilities for a particular subtype (EI, MI, or SI) on the x-axis, with the y-axis representing the count of specimens. The orange bars (“Class: X”) indicate specimens that truly belong to the relevant subtype, while the blue bars (“Rest”) represent all other classes. Notably, for each subtype, the probabilities for the correct class are centered toward higher values, whereas the probabilities for the non-matching classes cluster toward zero. This clear separation suggests that the model assigns high confidence to correct classifications and low confidence to incorrect ones, indicating strong discriminative capability. The one-vs-rest ROC curves show how effectively the model differentiates each target class from all other classes. The curves plot the true positive rate against the false positive rate at various probability thresholds. All three subtypes display curves that rise sharply and remain well above the diagonal (which represents random classification). The model demonstrates robust performance in classifying the EI, MI, and SI subtypes. The histograms highlight a strong distinction between each class and the combined “Rest” while the ROC curves confirm that this distinction persists over a range of classification thresholds. Together, these results suggest that the subtyping model is highly effective in accurately identifying immune subtypes and support its suitability for clinical applications where precise subtype classification is essential.

### Performance Metrics

The subtype wise Sensitivity, Specificity, and Area Under ROC Curve (AUC) in a one-versus-rest setting of FlahyRecovery is shown in **Table 4**. The subtyping model of FlahyRecovery has an average sensitivity of 79%, a macro average accuracy of 81%, and an average specificity of 89% across all the three subtypes.

**Table 4:** Subtype wise Performance Metrics listing Sensitivity, Specificity, and Area Under ROC Curve (AUC)

| **Immune Subtype** | **Positive Predictive Value** | **Sensitivity** | **Specificity** | **AUC** |
|---|---|---|---|---|
| Elevated Immunity (EI) | 0.83 | 0.78 | 0.94 | 0.96 |
| Moderate Immunity (MI) | 0.72 | 0.82 | 0.78 | 0.87 |
| Suppressed Immunity (SI) | 0.88 | 0.76 | 0.96 | 0.95 |

### Benchmarking Against an Established Reference

To independently validate the subtyping model’s predicted labels and biological relevance of our EI/MI/SI immune subtypes, we quantified immune cell infiltration using CIBERSORT-ABS and compared T-cell compartment signals across the three groups to which our test samples were assigned [49]. Across multiple T-cell subsets, a consistent stepwise pattern was observed, with EI showing the highest infiltration, MI intermediate, and SI the lowest (**Supplementary Figures S18 to S33**). This trend was most evident for CD8+ T cells and resting CD4+ memory T cells, both of which were enriched in EI compared to MI and SI. A similar distribution was observed for T follicular helper cells and regulatory T cells (Tregs), suggesting that the immune-enriched class is characterized by broader T-cell engagement rather than a single isolated lineage

## DISCUSSION

The present study contributes a distinct advancement to the immune-subtyping landscape. First, FlahyRecovery leverages a substantially broader functional gene panel (1,275 genes) than commonly used immune reference sets such as LM22 (547 genes) or the smaller immune-signature lists used in NMF-based clustering studies [28–34]. This allows the classifier to capture multi-dimensional immune activity including effector signaling, checkpoint pathways, stromal suppression, and antigen presentation rather than relying on limited cell-fraction signatures. Second, the model is trained across 14 cancer types, enabling pan-cancer generalizability and standardized EI/MI/SI classification, in contrast to existing cancer-specific immune subtype frameworks. Third, and most importantly, the EI/MI/SI subtypes generated by FlahyRecovery have direct therapeutic implications: EI tumors align with higher immunotherapy responsiveness, SI tumors with immune exclusion and combination therapy benefit, and MI tumors with mixed features. This direct mapping to clinical decision-making differentiates FlahyRecovery from descriptive immune state models and supports its potential utility as an actionable biomarker in precision oncology. It is important to note that FlahyRecovery is not a single molecular feature, but a machine-learning–based immune profiling classifier that generates a composite, multigene immune biomarker readout. Specifically, FlahyRecovery uses tumor gene expression data as a quantitative readout of immune signaling and immune–stromal interactions within the tumor microenvironment (TME). The role of the classifier is to map high-dimensional gene expression patterns onto clinically interpretable immune phenotypes of the TME – Elevated Immunity (EI), Moderate Immunity (MI), and Suppressed Immunity (SI). In this sense, the classifier serves as the *analytical engine*, while the output immune subtype represents the biomarker readout, analogous to how multigene assays (e.g., Oncotype DX, MammaPrint) function, that is as composite biomarkers rather than single molecular entities. Furthermore, the results emphasize the following aspects of clinical utility and application of FlahyRecovery.

### Classifier Model Selection

We evaluated multiple classifier families during model development, including neural network–based classifiers and ensemble tree–based methods. While neural networks achieved comparable performance during exploratory testing, we did not adopt them as the final model due to limitations in interpretability and clinical transparency. In contrast, the Random Forest classifier provided a strong balance between predictive performance, robustness to noise, and biological interpretability.

The feature importance scores analysis presented in the model testing section, enabled direct mapping of the learned decision rules to known immune pathways (e.g., antigen presentation, T-cell signalling, chemokine signalling), which is critical for reliability and biological validation. Given the intended clinical application of FlahyRecovery, we therefore selected a Random Forest–based classifier as the final model architecture

### Informing Treatment Decisions

By clearly delineating tumors into Elevated (hot), Moderate (intermediate), or Suppressed (cold) immune states, the biomarker will assist oncologists in selecting therapies most likely to benefit individual patients. Hot tumors, for instance, respond robustly to immunotherapies like PD-1/PD-L1 inhibitors, while cold tumors might warrant strategies to enhance T-cell infiltration or combination therapies.

### Prognostic Value

Elevated immunity within the TME correlates with increased OS in most cancer types and increased therapeutic responsiveness to immunotherapy. Suppressed immunity correlates with more aggressive disease trajectories for Breast Cancer, Non-Small Cell Lung Cancer, Gastric Cancer, Head & Neck Cancer, Skin Cancer, Colon Cancer, Esophageal Cancer, Bladder Cancer, Liver Cancer, Pancreatic Cancer, and Cervical Cancer. FlahyRecovery’s capacity to stratify patients based on these profiles offers valuable precision medicine insights, and informing clinicians to tailor surveillance and follow-up plans, leading to timely treatment changes for a better outcome.

### Multi-Tumor Applicability

The FlahyRecovery subtyping model’s extensive training set, encompassing a broad spectrum of tumor types (breast, non-small cell lung, head and neck, gastric, skin, colon, kidney, pancreatic, bladder, esophageal, prostate, brain, cervical, and liver cancer), underscores its versatility. This wide applicability ensures that clinicians can use the biomarker in diverse oncology contexts.

### Accuracy and Reproducibility

FlahyRecovery builds on established immune subtyping principles and enhances them with a robust, AI-driven classification system. Its high sensitivity (79%), specificity (89%), and AUC (0.93) demonstrates consistent, reproducible classification across multiple solid tumors. The gene-expression based approach ensures a more holistic immune signature capture compared to tests using a single immunohistochemistry based marker, allowing clinicians to gain deeper insights into the functional state of the TME.

### Comparison with Existing Subtyping Methods

Existing immune subtyping methodologies have undoubtedly advanced our understanding of cancer immunology. However, most rely on limited narrow genomic signatures, or single immunohistochemistry-based markers. Beyond transcriptome based profiling, several complementary technologies are widely used to investigate the tumor microenvironment. Conventional chromogenic immunohistochemistry (IHC) and hematoxylin–eosin–based tumor infiltrating lymphocyte scoring remain routine clinical methods to quantify key markers such as PD L1 and CD8, but they are limited to one or a few markers per section and suffer from observer-to observer variability [50,51]. Multiplex IHC/immunofluorescence (mIHC/IF) enables simultaneous detection of multiple immune and stromal markers on a single formalin fixed, paraffin embedded tissue section, providing quantitative, spatially resolved maps of the tumor immune microenvironment that can be linked to prognosis and immunotherapy response [50,51]. High parameter imaging platforms, including imaging mass cytometry, allow in situ measurement of 30–40 or more proteins at subcellular resolution, thereby revealing spatial arrangements of tumor, immune, and stromal cells and identifying prognostic TME architectures across diverse cancers [52]. More recently, spatial multi omics and spatial proteomics technologies integrate high plex protein detection with spatial transcriptomics or other omics layers to dissect the localization and interaction of cell populations within intact tissue architecture, uncovering microenvironmental niches associated with therapy resistance and clinical outcome. These histology and protein based approaches provide rich spatial context and single cell–level phenotyping, but they are constrained by panel size, cost, and scalability, underscoring the complementary value of scalable, bulk transcriptome–driven classifiers such as FlahyRecovery for routine, pan cancer [53–55] immune subtyping. These allow clinicians to gain deeper insights into the functional state of the T M E. When compared with established immune-profiling methodologies, FlahyRecovery represents a distinct contribution. TCGA’s six immune subtypes offer a valuable pan-cancer reference for broad immune states, but they were derived using metagene-score clustering rather than supervised classification, and they do not provide a streamlined three-state framework optimized for clinical interpretability [8]. Likewise, CIBERSORT and similar deconvolution tools remain the standard for estimating immune-cell proportions, but they do not perform immune subtyping and are constrained to the fixed 547-gene LM22 matrix, which captures cell-fraction patterns rather than integrated pathway-level immune activity [28]. Tumor-specific NMF-based immune subtype studies generate meaningful clusters, but their narrow gene lists and cancer-specific optimization limit generalizability across tumor types [29–34]. In contrast, FlahyRecovery integrates a broader functional immune-signaling panel and a machine-learning classifier trained across 14 cancers, enabling consistent EI/MI/SI subtyping at scale. This positions FlahyRecovery not as a replacement for existing tools, but as a complementary cross-cancer subtyping framework that captures global immune activation and suppression states with high predictive performance and enhanced clinical relevance.

### Evidence and Validation

From retrospective analyses to prospective clinical applications, results from numerous studies have consistently shown that tumors classified as EI have higher immunotherapy response rates and improved patient survival curves, and that tumors classified as SI have higher combination therapy response rates and thus, improved patient survival curves. Clinicians can therefore have confidence in usage of FlahyRecovery.

### Further inclusions to sub-typing model input for enhanced immune subtyping and treatment personalization

Apart from gene expression data, the copy number alteration and somatic mutation data of the patients are also being analyzed for all the mentioned cancers. The process includes identifying the amplified, deleted, or mutated genes, and processing these alterations via an AI model for enhanced immune subtyping of solid tumors for subsequent treatment recommendations. Currently, we have identified ∼100 gene candidates, whose copy number and mutation data can be used to identify the tumor immune-subtype and predict the potential therapeutic outcomes.

## CONCLUSION

Using artificial intelligence to process patient tumor biology works well for immune subtyping and yielding actionable clinical insights. The developed immune profiling biomarker provides robust TME characterization, and equips oncologists with AI driven clarity, ultimately informing personalized treatment decisions. FlahyRecovery’s comprehensive approach to immune subtyping helps translate cutting-edge research into tangible patient benefits, fulfilling the promise of truly personalized cancer care.

## AUTHOR CONTRIBUTION

Jagjit Singh (Conceptualization, Methodology, Data Curation, Formal analysis, Software, Visualization, Validation, Writing – original draft, Writing – review and editing), Pragya Singh (Validation, Data Curation, Formal Analysis, Writing – review and editing), Christopher E Mason (Conceptualization, Writing – review and editing), Sonia (Validation, Data Curation, Formal Analysis, Writing – review and editing).

## Ethics approval

This study is a secondary analysis of publicly available, de-identified data from The Cancer Genome Atlas (TCGA) and CPTAC-3 cohorts listed in Table 3, accessed through the National Cancer Institute (NCI) Genomic Data Commons (GDC). No new patient samples were collected and there was no contact with patients. Ethical approval and informed consent were obtained by the original data-generating studies, and no additional ethics review was required for this analysis.

## Consent to participate

Not applicable. This study used publicly available, de-identified data; informed consent was obtained by the original studies.

## Consent for publication

Not applicable. No individual-level identifiable data is used in this manuscript.

## Data Availability

All data used in this study is available for download at the The Cancer Genome Atlas database.

---

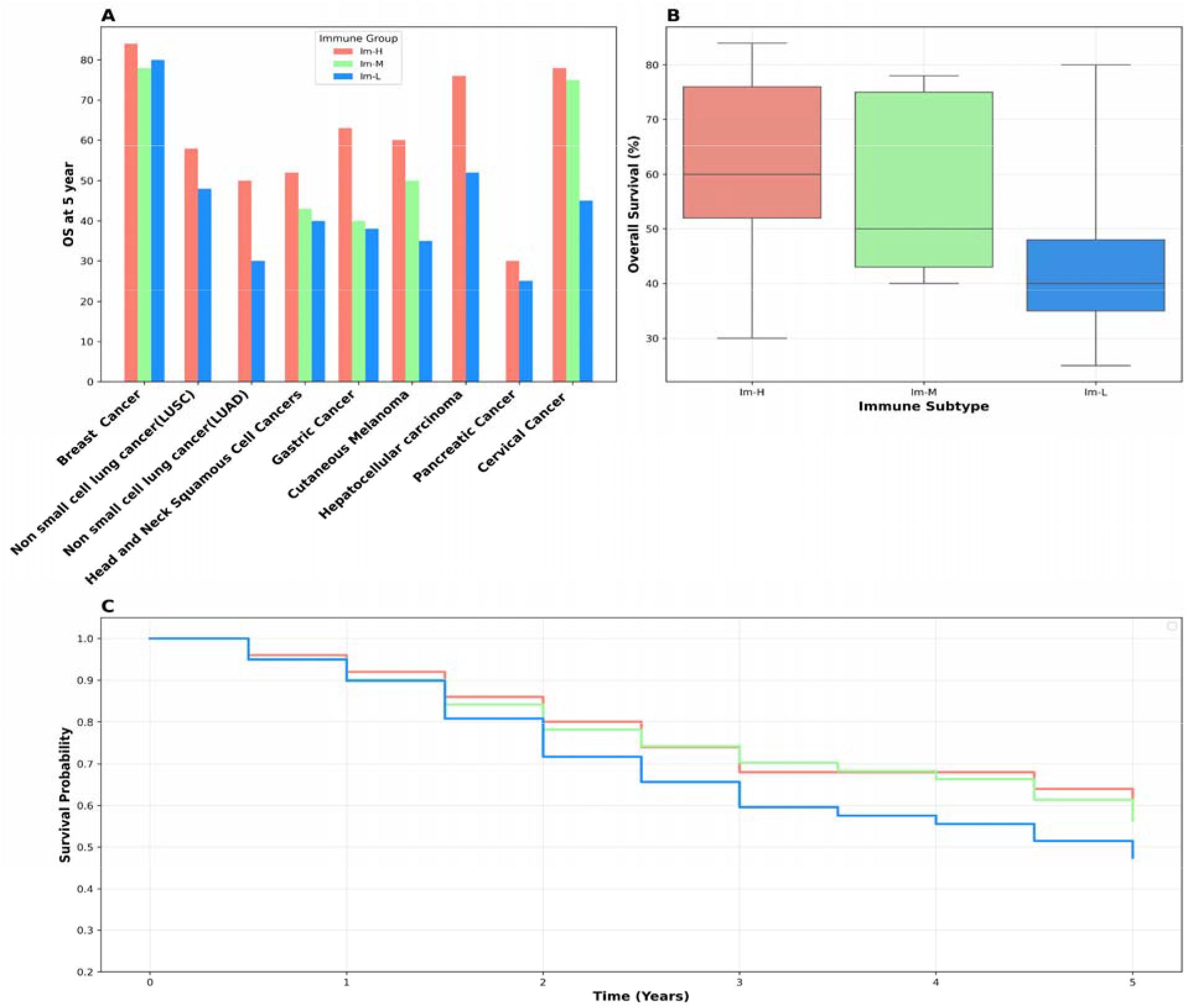

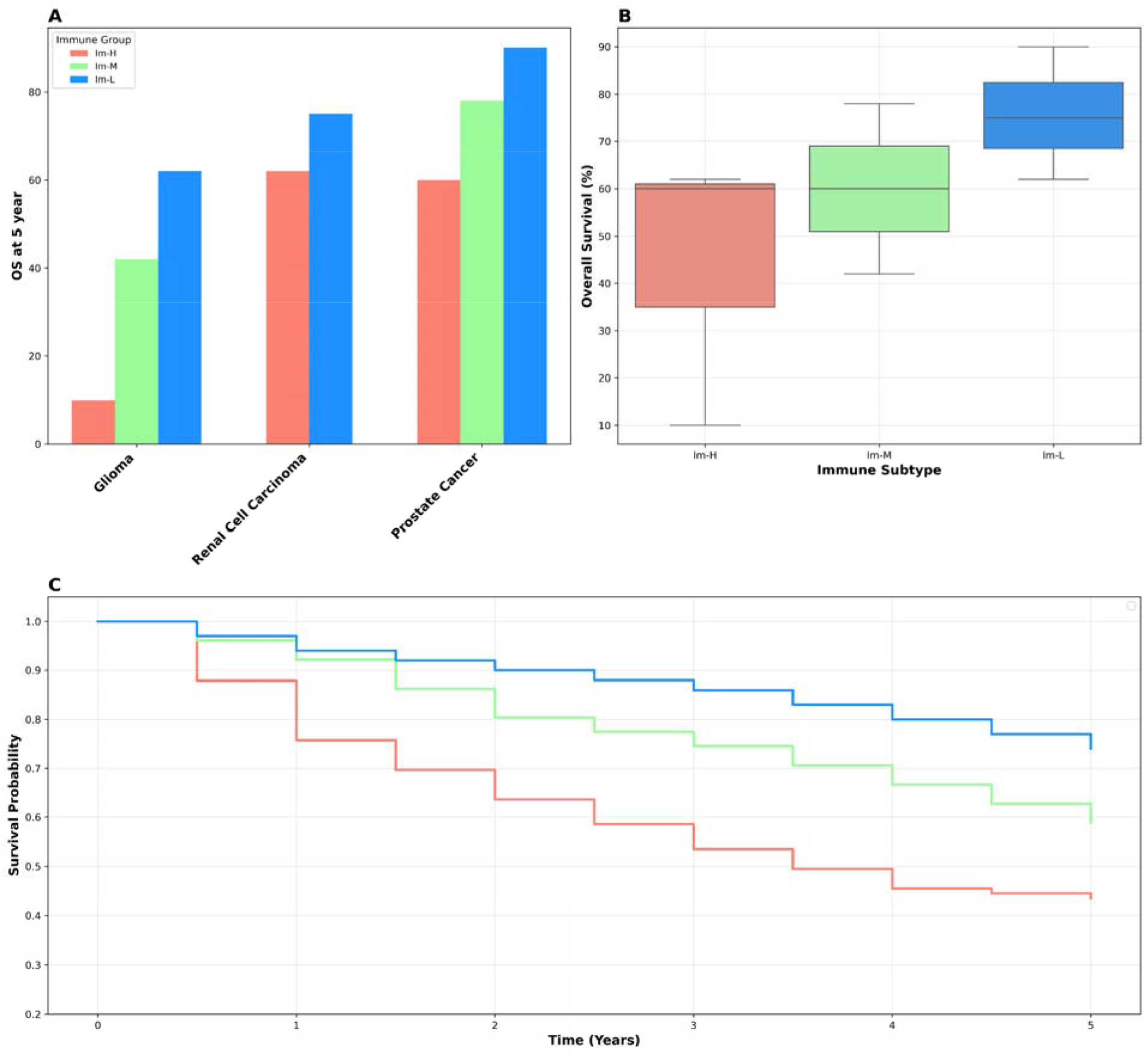

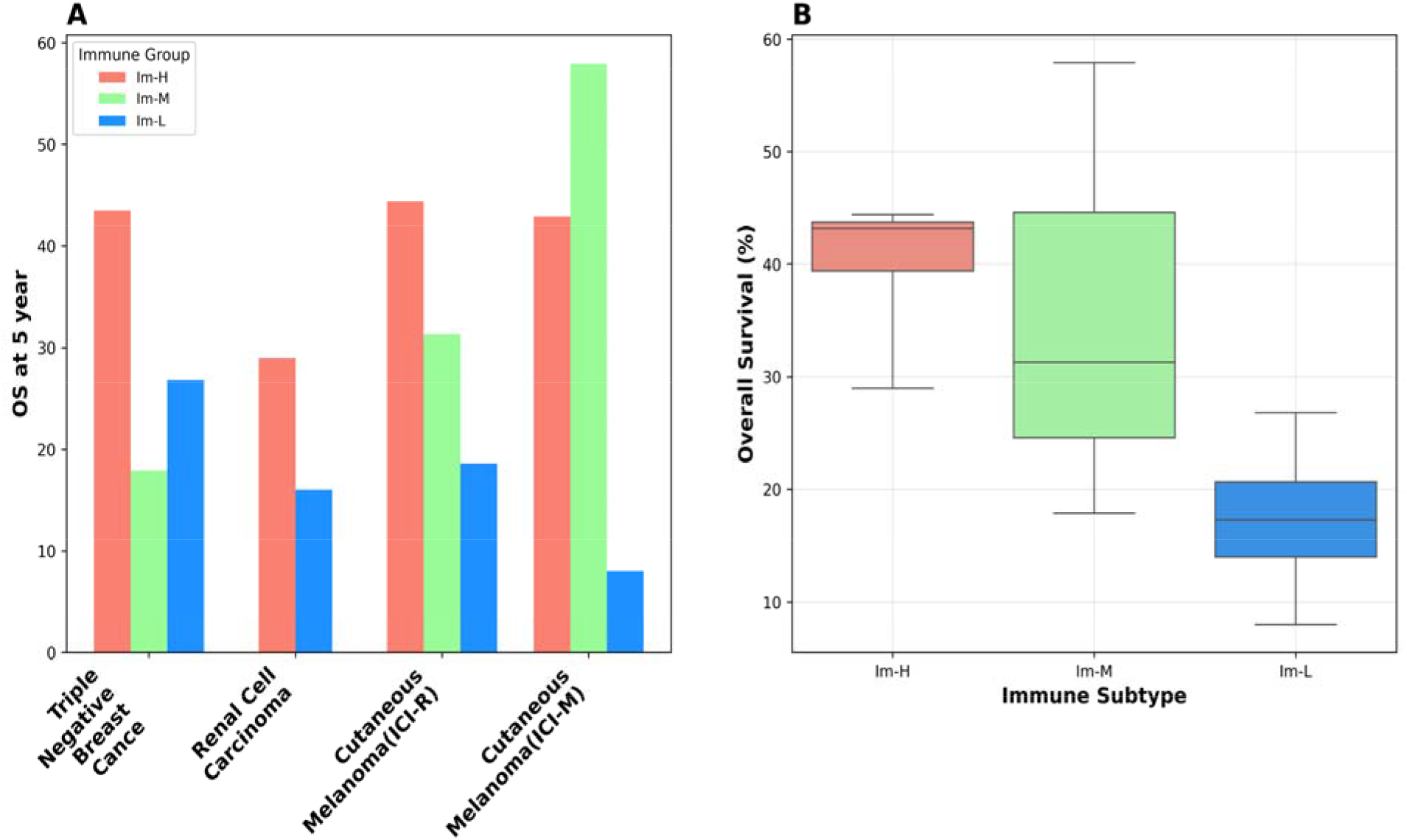

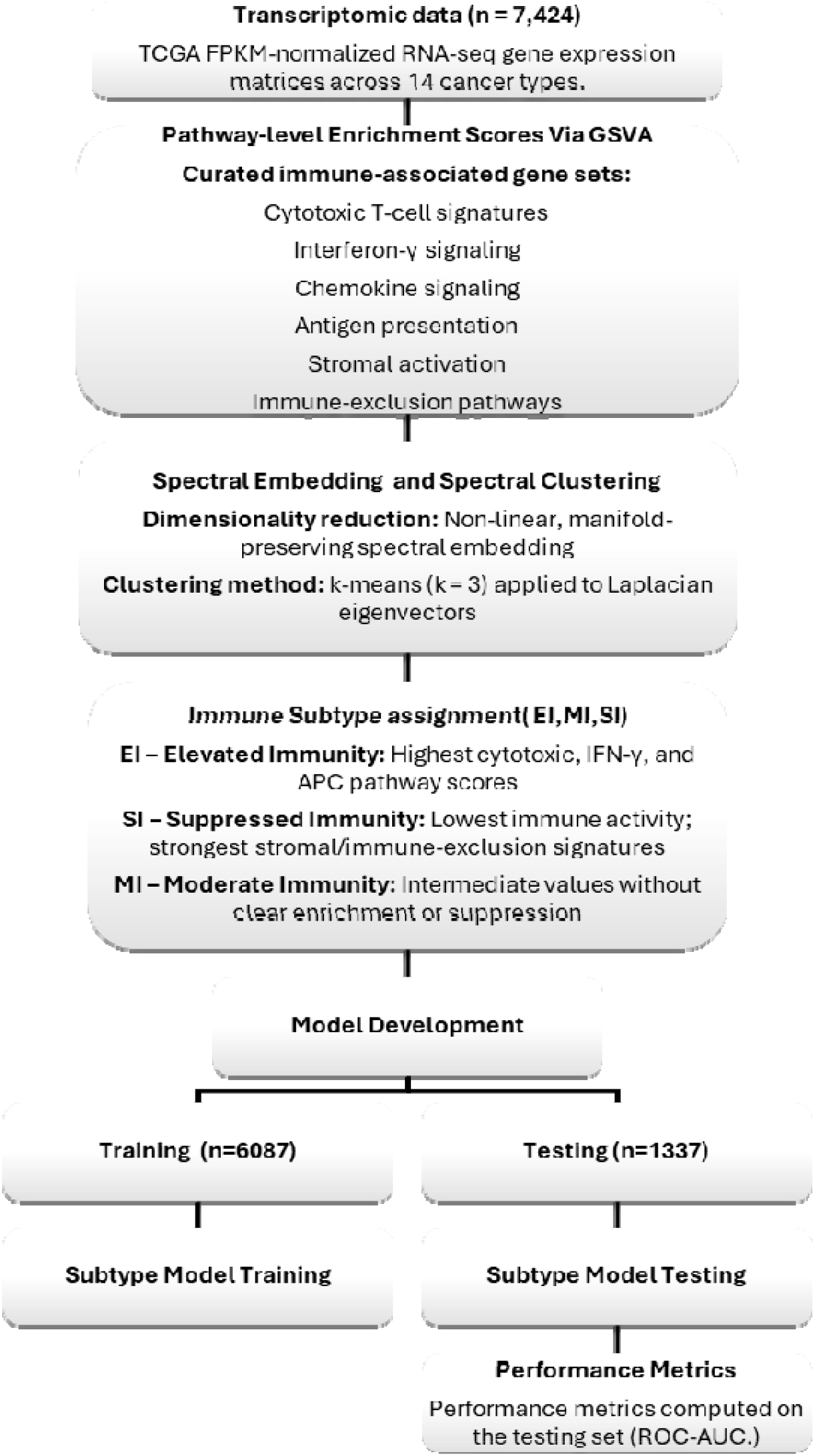

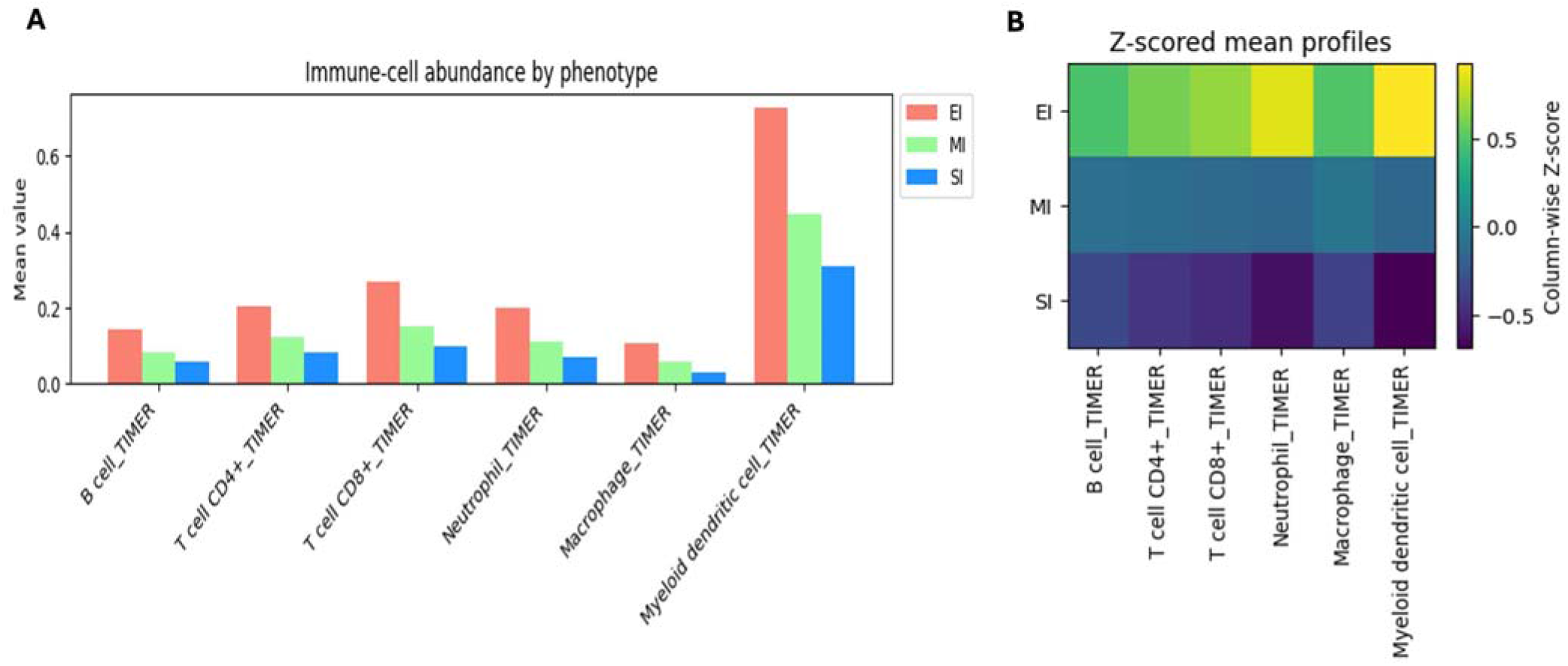

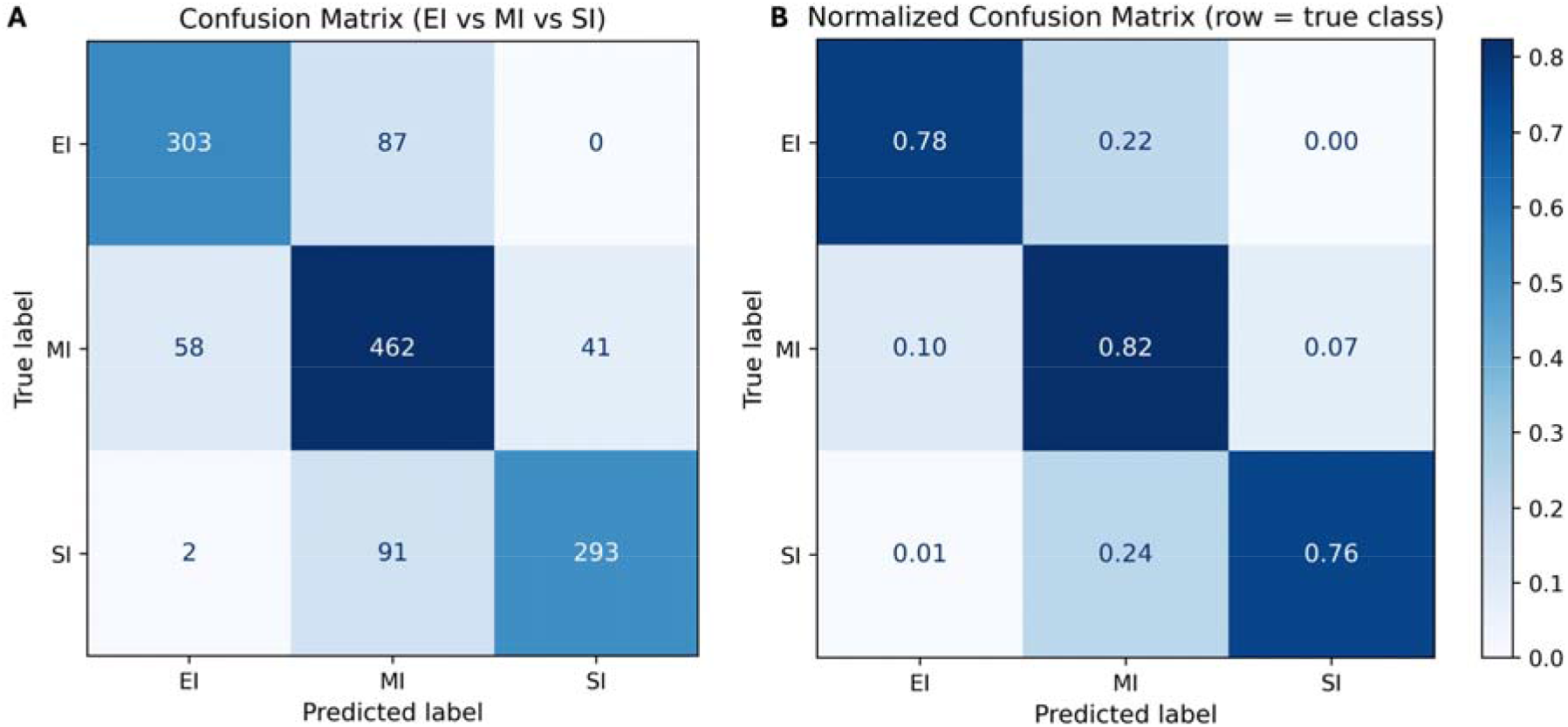

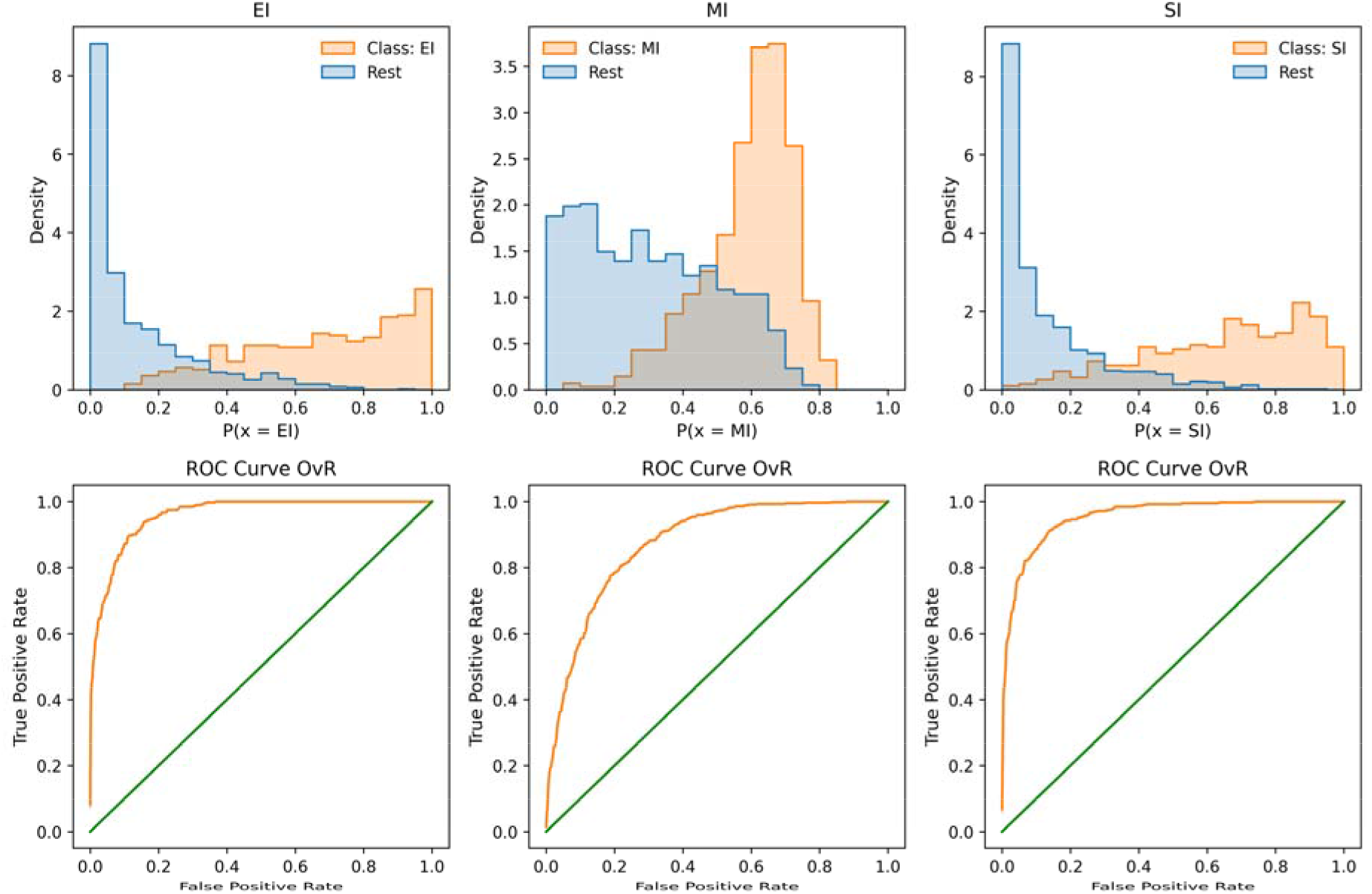

