## Supplementary Information for "FlahyRecovery is an AI & gene expression based immune profiling biomarker for decision support in precision oncology"

**TABLE OF CONTENTS**

| **S.No.** | **Content** | **Page No.** |
| --- | --- | --- |
| 1. | Figure S1: Immune Cell Distribution in EI, MI, SI for BLCA. | 4 |
| 2. | Figure S2: Immune Cell Distribution in EI, MI, SI for BRCA. | 5 |
| 3. | Figure S3: Immune Cell Distribution in EI, MI, SI for CESC. | 6 |
| 4. | Figure S4: Immune Cell Distribution in EI, MI, SI for COAD. | 7 |
| 5. | Figure S5: Immune Cell Distribution in EI, MI, SI for ESCA. | 8 |
| 6. | Figure S6: Immune Cell Distribution in EI, MI, SI for GBM. | 9 |
| 7. | Figure S7: Immune Cell Distribution in EI, MI, SI for HNSC. | 10 |
| 8. | Figure S8: Immune Cell Distribution in EI, MI, SI for KIRC. | 11 |
| 9. | Figure S9: Immune Cell Distribution in EI, MI, SI for KIRP. | 12 |
| 10. | Figure S10: Immune Cell Distribution in EI, MI, SI for LIHC. | 13 |
| 11. | Figure S11: Immune Cell Distribution in EI, MI, SI for LUAD. | 14 |
| 12. | Figure S12: Immune Cell Distribution in EI, MI, SI for LUSC. | 15 |
| 13. | Figure S13: Immune Cell Distribution in EI, MI, SI for PAAD. | 16 |
| 14. | Figure S14: Immune Cell Distribution in EI, MI, SI for PRAD. | 17 |
| 15. | Figure S15: Immune Cell Distribution in EI, MI, SI for SKCM. | 18 |
| 16. | Figure S16: Immune Cell Distribution in EI, MI, SI for STAD. | 19 |
| 17. | Figure S17: T-regulatory Cell Distribution in EI, MI, SI for all the 14 cancers. | 20 |
| 18 | Figure S18: Immune cell infiltration across EI, MI, and SI groups in BLCA. | 21 |
| 19 | Figure S19: Immune cell infiltration across EI, MI, and SI groups in BRCA. | 22 |
| 20 | Figure S20: Immune cell infiltration across EI, MI, and SI groups in CESC. | 23 |
| 21 | Figure S21: Immune cell infiltration across EI, MI, and SI groups in COAD. | 24 |
| 22 | Figure S22: Immune cell infiltration across EI, MI, and SI groups in ESCA. | 25 |
| 23 | Figure S23: Immune cell infiltration across EI, MI, and SI groups in GBM. | 26 |
| 24 | Figure S24: Immune cell infiltration across EI, MI, and SI groups in HNSC. | 27 |
| 25 | Figure S25: Immune cell infiltration across EI, MI, and SI groups in KIRC. | 28 |
| 26 | Figure S26: Immune cell infiltration across EI, MI, and SI groups in KIRP. | 29 |
| 27 | Figure S27: Immune cell infiltration across EI, MI, and SI groups in LIHC. | 30 |
| 28 | Figure S28: Immune cell infiltration across EI, MI, and SI groups in LUAD. | 31 |
| 29 | Figure S29: Immune cell infiltration across EI, MI, and SI groups in LUSC. | 32 |
| 30 | Figure S30: Immune cell infiltration across EI, MI, and SI groups in PAAD. | 33 |
| 31 | Figure S31: Immune cell infiltration across EI, MI, and SI groups in PRAD. | 34 |
| 32 | Figure S32: Immune cell infiltration across EI, MI, and SI groups in SKCM. | 35 |
| 33 | Figure S33: Immune cell infiltration across EI, MI, and SI groups in STAD. | 36 |
| 34. | Supplementary Table S1: Standard Deviation of Bar plots of Pan-cancer Immune Cell Infiltration | 37 |
| 35. | Supplementary Table S2: P-values of Statistical tests performed between tumor immune-subtype (pan-cancer) for different immune cells | 38 |
| 36. | Supplementary Tables S3-S18: P-values of Statistical tests performed between tumor immune-subtype of individual cancer types for different immune cells | 39-54 |
| 37. | Supplementary Table S19: P-values of Statistical tests performed between tumor immune-subtype of different cancer types for T-regulatory cell distribution | 55-56 |

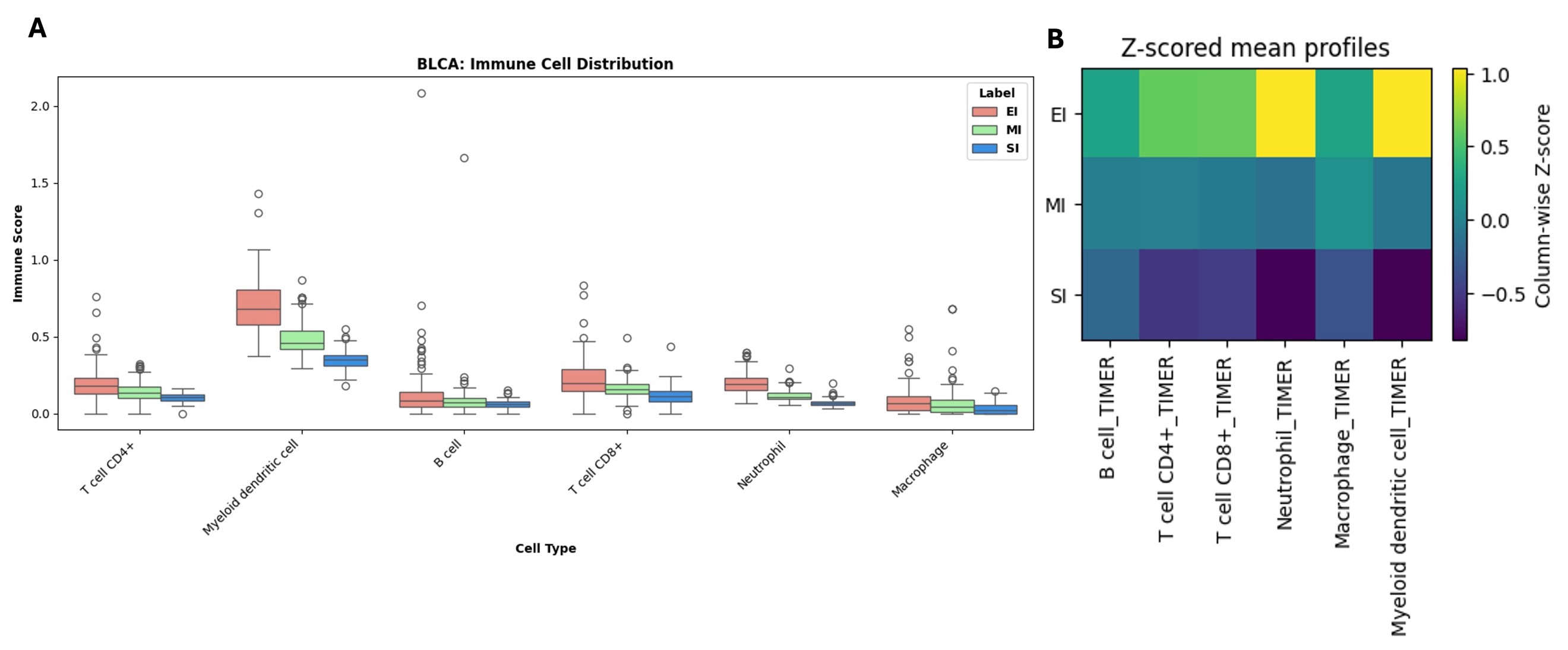

**Figure S1: (A)** Box plot depicting the distribution of each immune cell type by phenotype [EI (red), MI (green), SI (blue)] in BLCA. EI (n = 100), MI (n = 108), SI (n = 113). **(B)** Heatmap showing Z-scored mean profiles of six immune cell types [B cell, CD4+ T cell, CD8+ T cell, Neutrophil, Macrophage, and Myeloid dendritic cell] across three phenotypic groups: EI, MI, and SI in BLCA. Values are column-wise Z-scores, as indicated by the color bar.

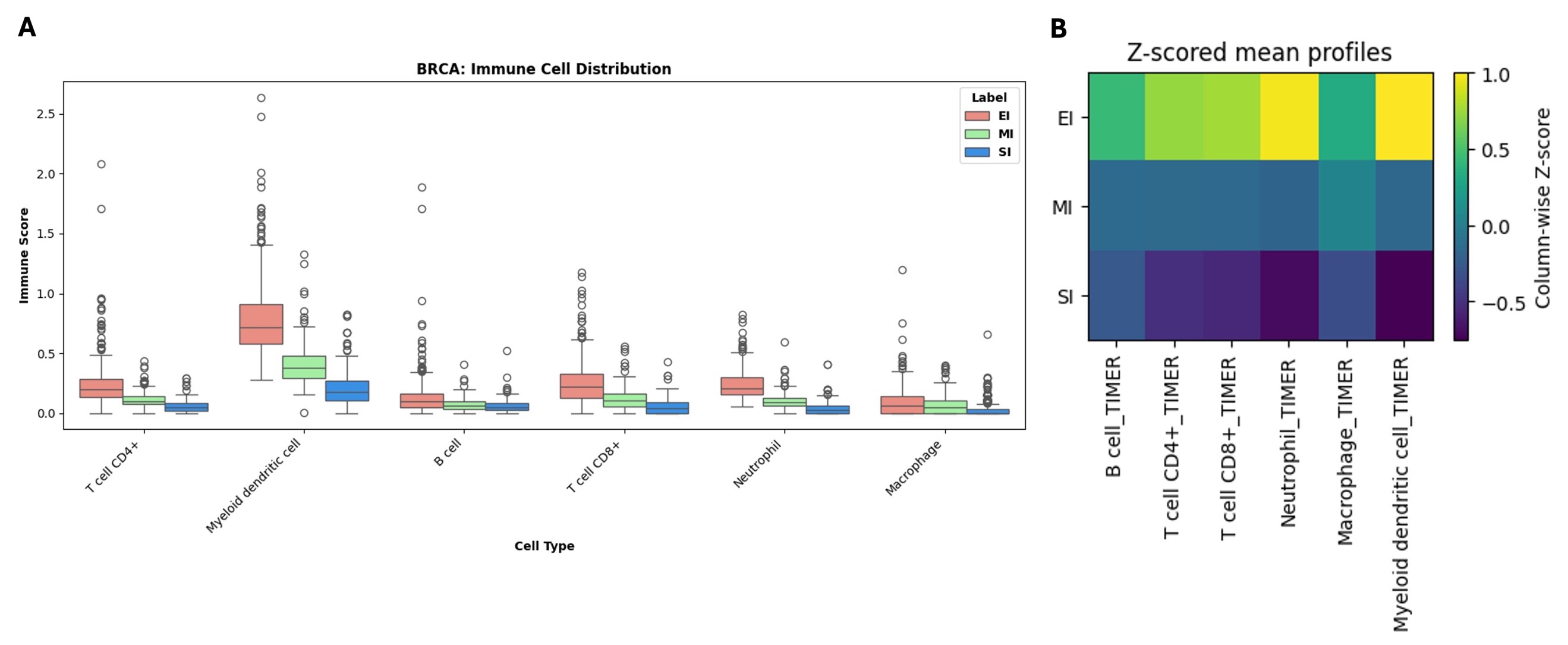

**Figure S2: (A)** Box plot depicting the distribution of each immune cell type by phenotype [EI (red), MI (green), SI (blue)] in BRCA. EI (n = 276), MI (n = 282), SI (n = 308). **(B)** Heatmap showing Z-scored mean profiles of six immune cell types [B cell, CD4+ T cell, CD8+ T cell, Neutrophil, Macrophage, and Myeloid dendritic cell] across three phenotypic groups: EI, MI, and SI in BRCA. Values are column-wise Z-scores, as indicated by the color bar.

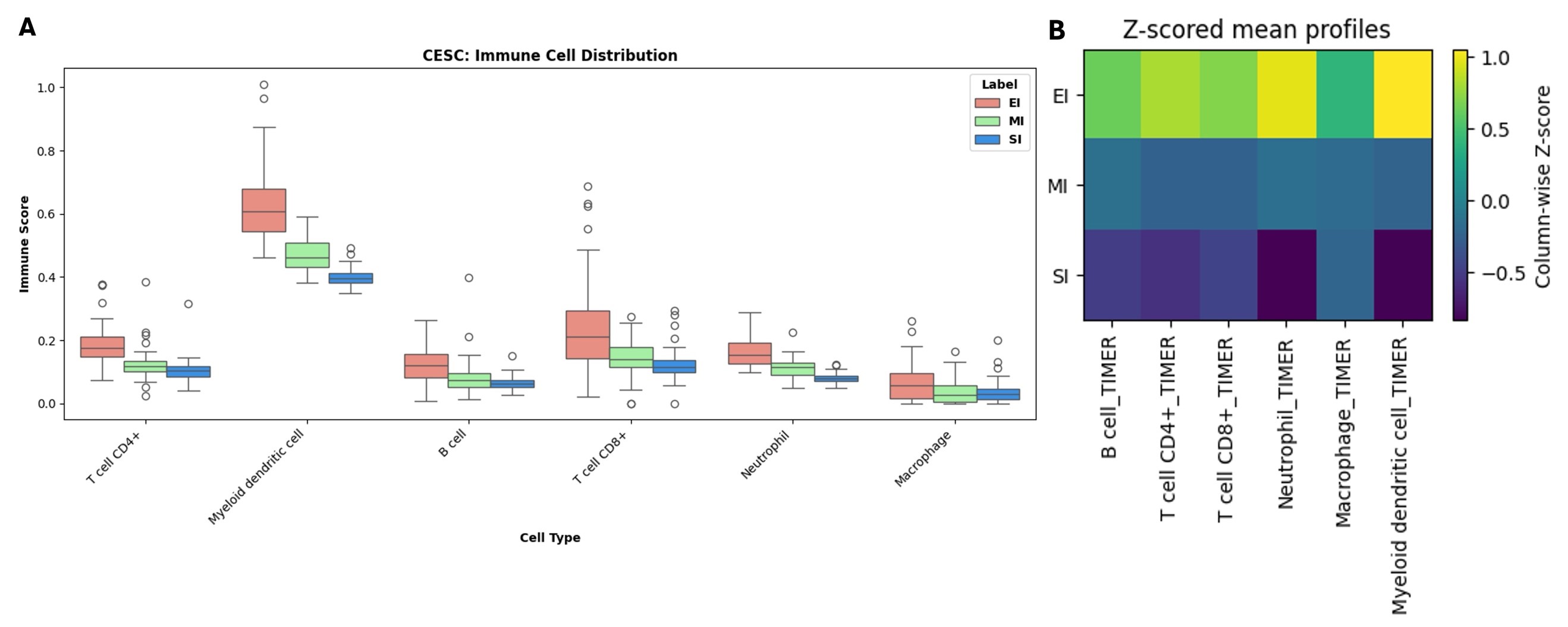

**Figure S3: (A)** Box plot depicting the distribution of each immune cell type by phenotype [EI (red), MI (green), SI (blue)] in CESC. EI (n = 62), MI (n = 63), SI (n = 62). **(B)** Heatmap showing Z-scored mean profiles of six immune cell types [B cell, CD4+ T cell, CD8+ T cell, Neutrophil, Macrophage, and Myeloid dendritic cell] across three phenotypic groups: EI, MI, and SI in CESC. Values are column-wise Z-scores, as indicated by the color bar.

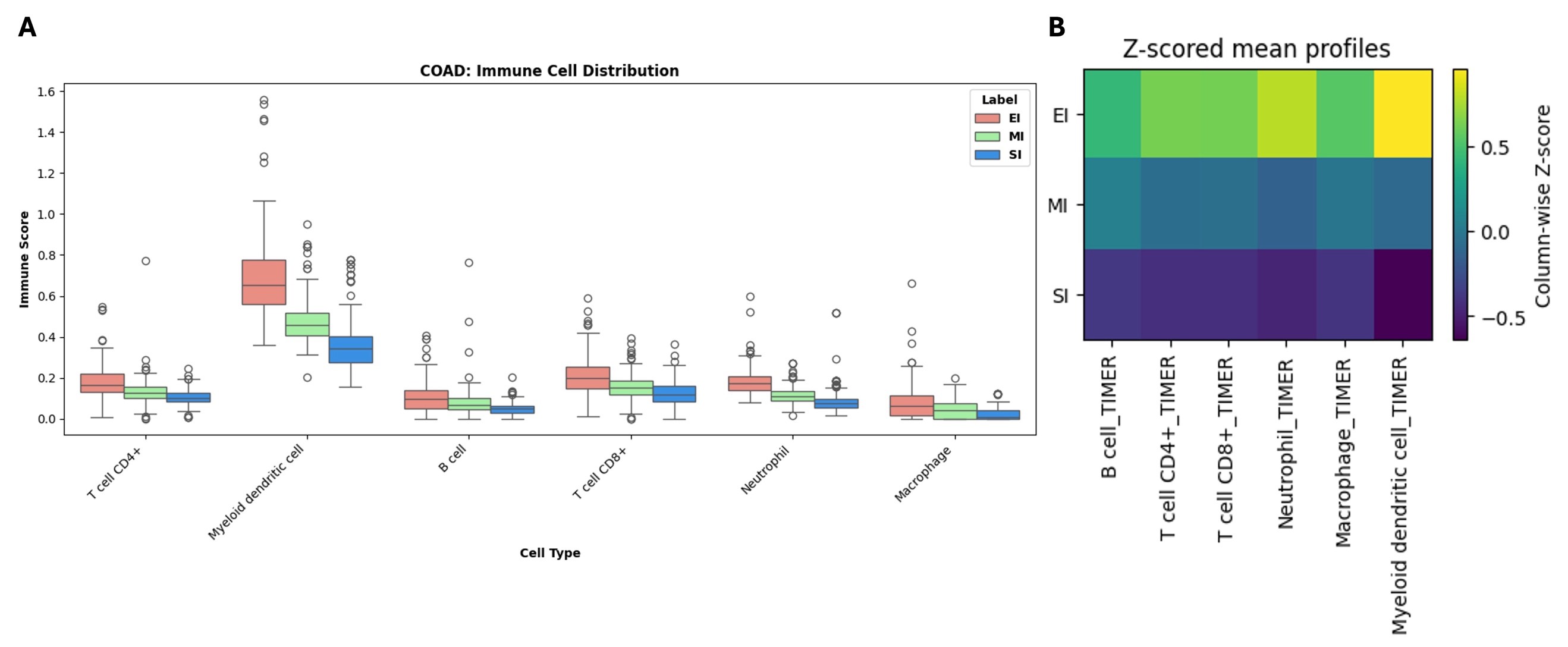

**Figure S4: (A)** Box plot depicting the distribution of each immune cell type by phenotype [EI (red), MI (green), SI (blue)] in COAD. EI (n = 116), MI (n = 142), SI (n = 154). **(B)** Heatmap showing Z-scored mean profiles of six immune cell types [B cell, CD4+ T cell, CD8+ T cell, Neutrophil, Macrophage, and Myeloid dendritic cell] across three phenotypic groups: EI, MI, and SI in COAD. Values are column-wise Z-scores, as indicated by the color bar.

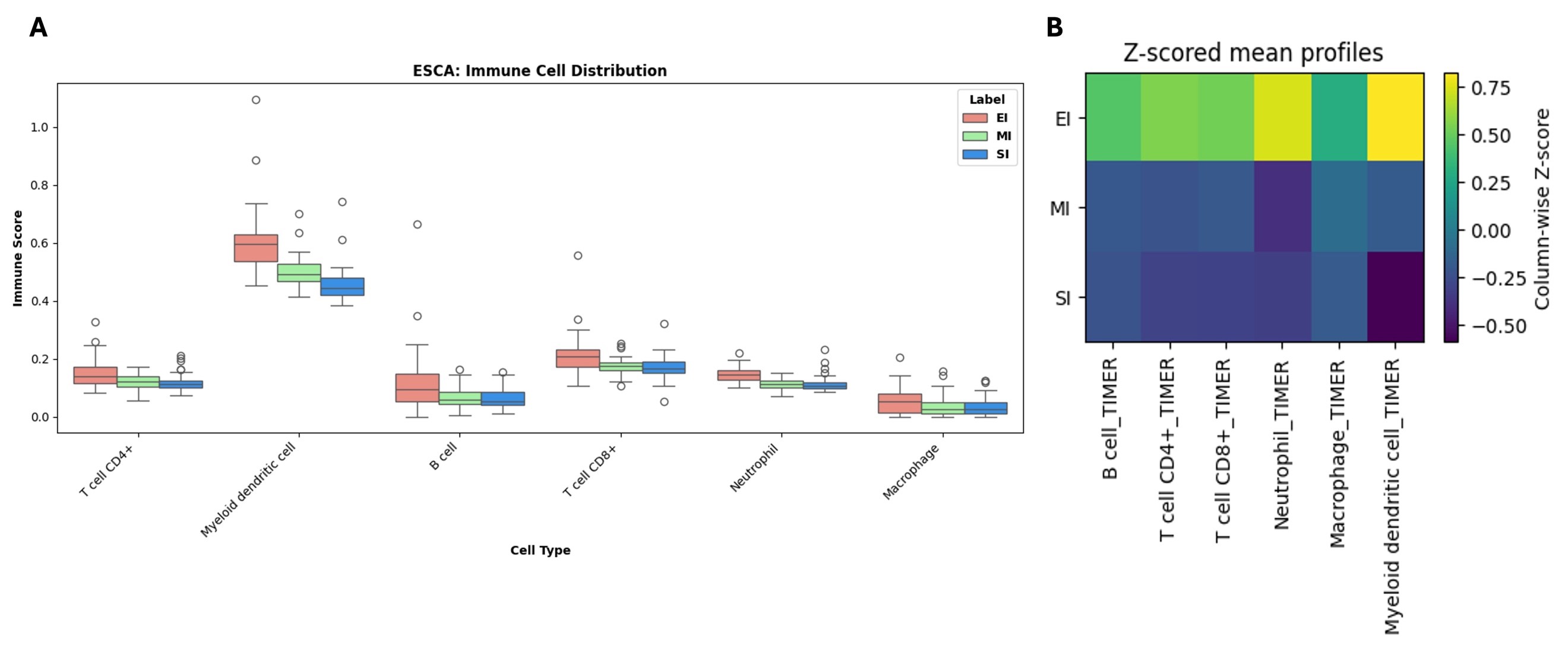

**Figure S5: (A)** Box plot depicting the distribution of each immune cell type by phenotype [EI (red), MI (green), SI (blue)] in ESCA. EI (n = 39), MI (n = 38), SI (n = 43). **(B)** Heatmap showing Z-scored mean profiles of six immune cell types [B cell, CD4+ T cell, CD8+ T cell, Neutrophil, Macrophage, and Myeloid dendritic cell] across three phenotypic groups: EI, MI, and SI in ESCA. Values are column-wise Z-scores, as indicated by the color bar.

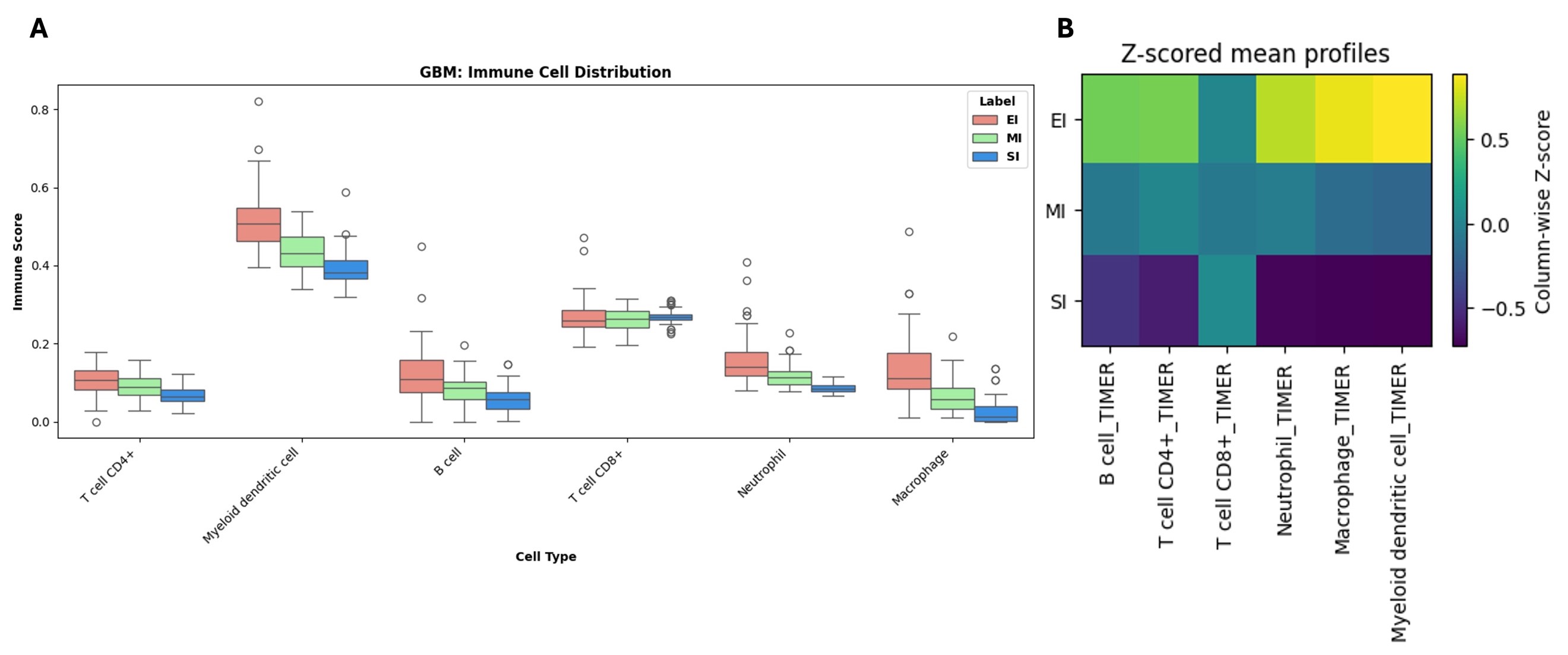

**Figure S6: (A)** Box plot depicting the distribution of each immune cell type by phenotype [EI (red), MI (green), SI (blue)] in GBM. EI (n = 66), MI (n = 63), SI (n = 64). **(B)** Heatmap showing Z-scored mean profiles of six immune cell types [B cell, CD4+ T cell, CD8+ T cell, Neutrophil, Macrophage, and Myeloid dendritic cell] across three phenotypic groups: EI, MI, and SI in GBM. Values are column-wise Z-scores, as indicated by the color bar.

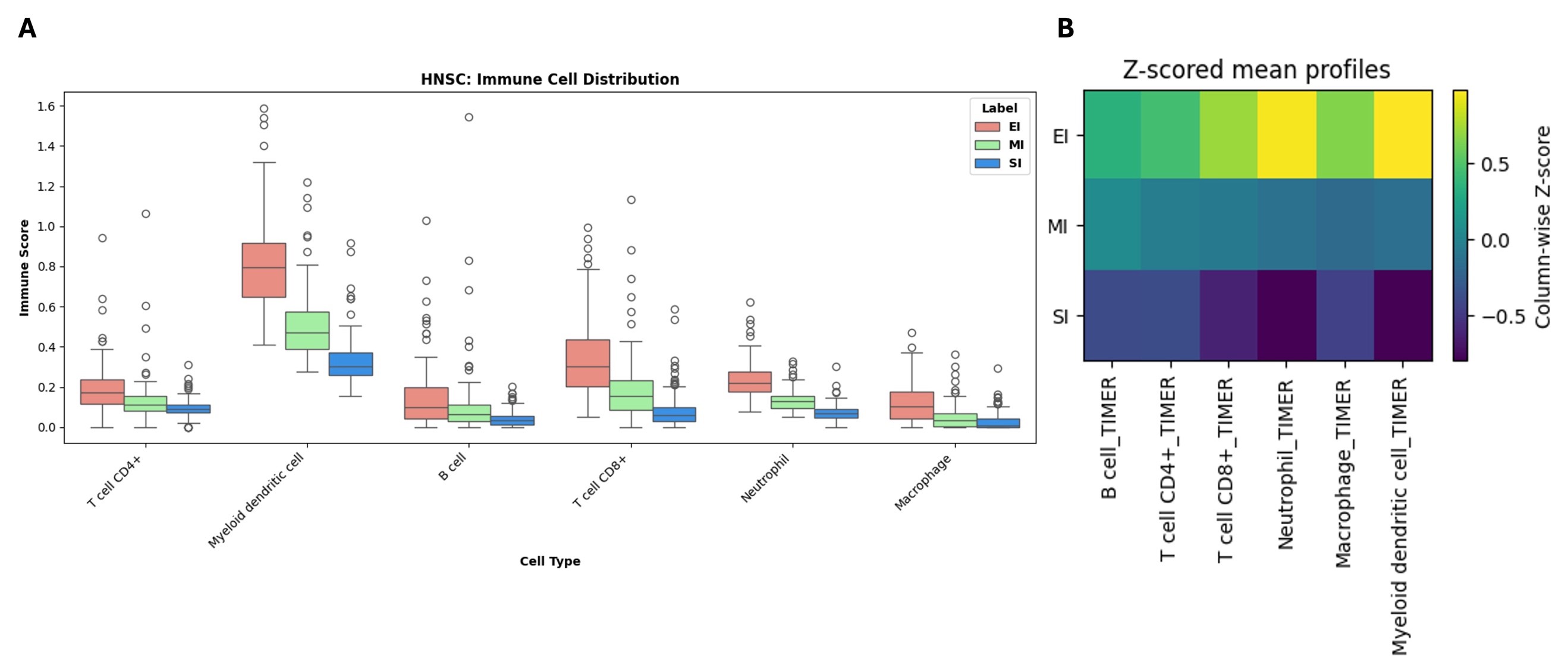

**Figure S7: (A)** Box plot depicting the distribution of each immune cell type by phenotype [EI (red), MI (green), SI (blue)] in HNSC. EI (n = 111), MI (n = 106), SI (n = 117). **(B)** Heatmap showing Z-scored mean profiles of six immune cell types [B cell, CD4+ T cell, CD8+ T cell, Neutrophil, Macrophage, and Myeloid dendritic cell] across three phenotypic groups: EI, MI, and SI in HNSC. Values are column-wise Z-scores, as indicated by the color bar.

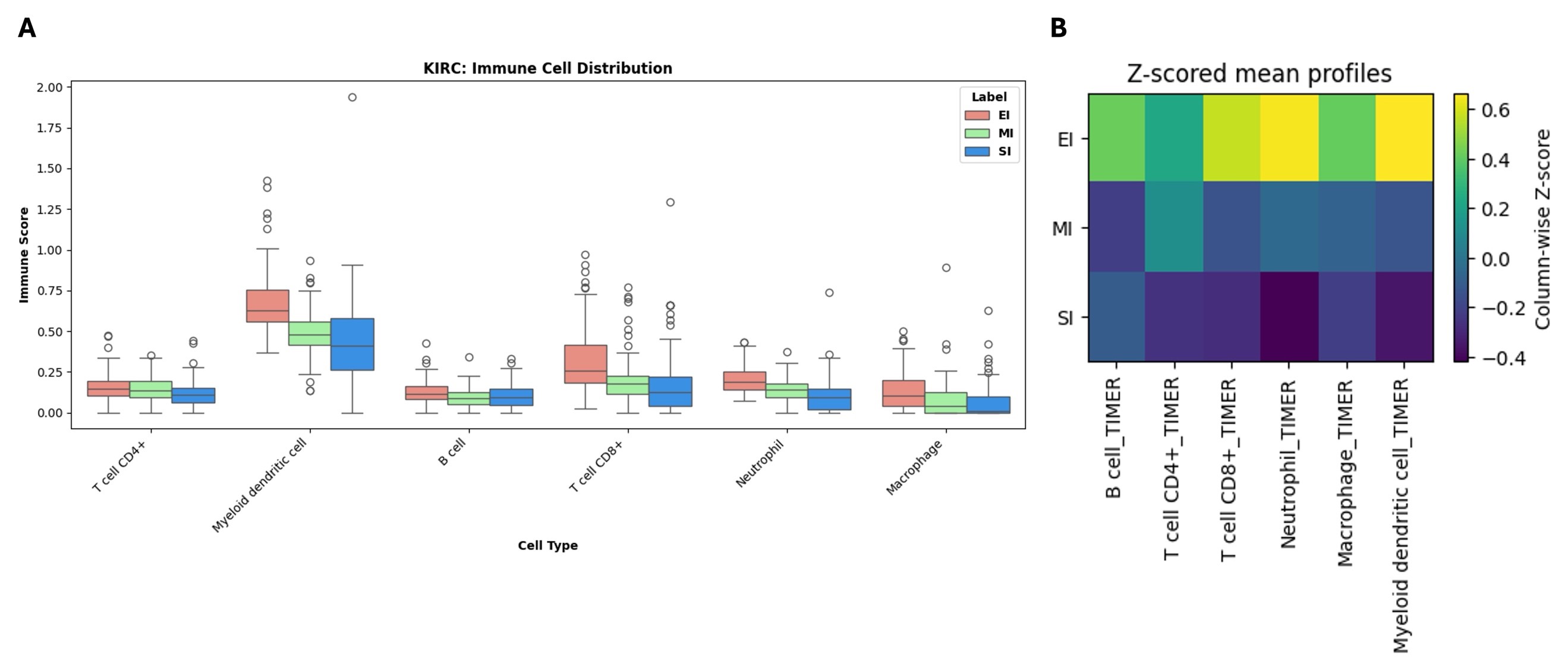

**Figure S8: (A)** Box plot depicting the distribution of each immune cell type by phenotype [EI (red), MI (green), SI (blue)] in KIRC. EI (n = 88), MI (n = 107), SI (n = 124). **(B)** Heatmap showing Z-scored mean profiles of six immune cell types [B cell, CD4+ T cell, CD8+ T cell, Neutrophil, Macrophage, and Myeloid dendritic cell] across three phenotypic groups: EI, MI, and SI in KIRC. Values are column-wise Z-scores, as indicated by the color bar.

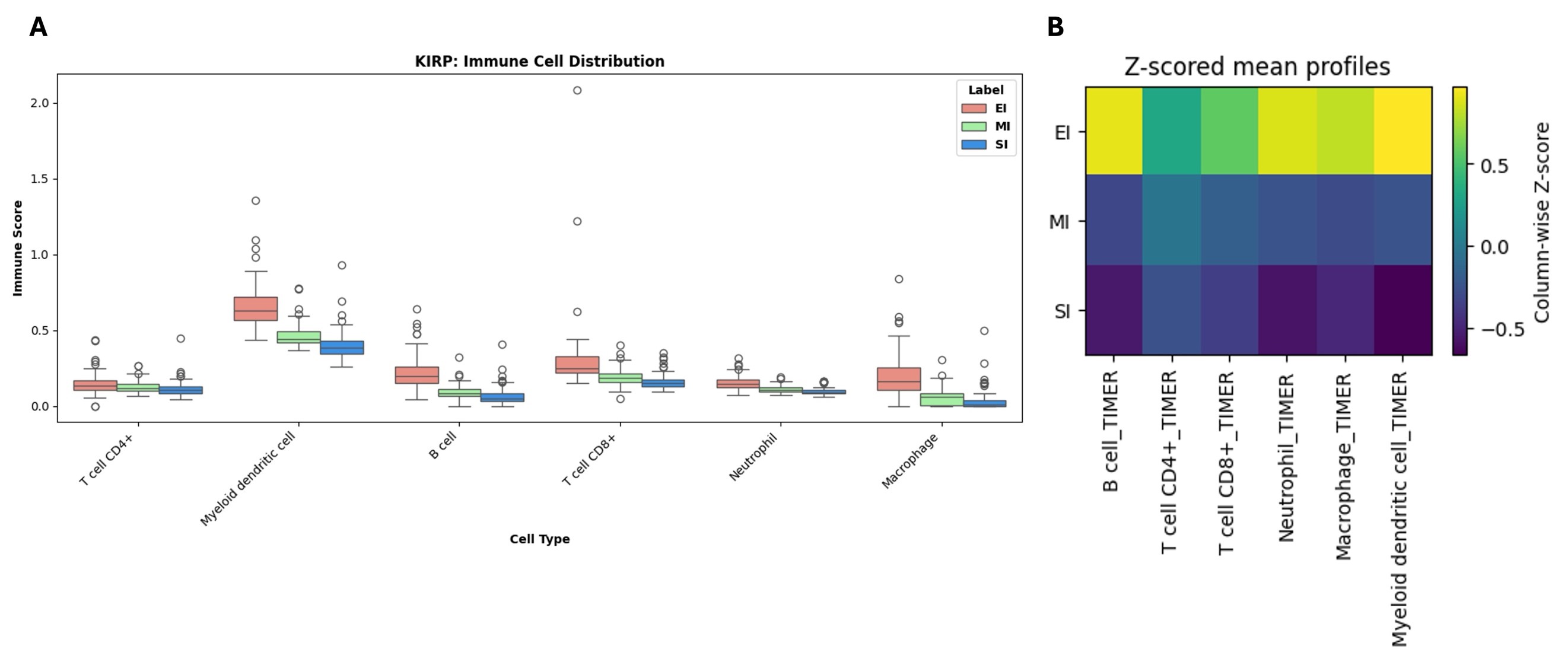

**Figure S9: (A)** Box plot depicting the distribution of each immune cell type by phenotype [EI (red), MI (green), SI (blue)] in KIRP. EI (n = 68), MI (n = 69), SI (n = 74). **(B)** Heatmap showing Z-scored mean profiles of six immune cell types [B cell, CD4+ T cell, CD8+ T cell, Neutrophil, Macrophage, and Myeloid dendritic cell] across three phenotypic groups: EI, MI, and SI in KIRP. Values are column-wise Z-scores, as indicated by the color bar.

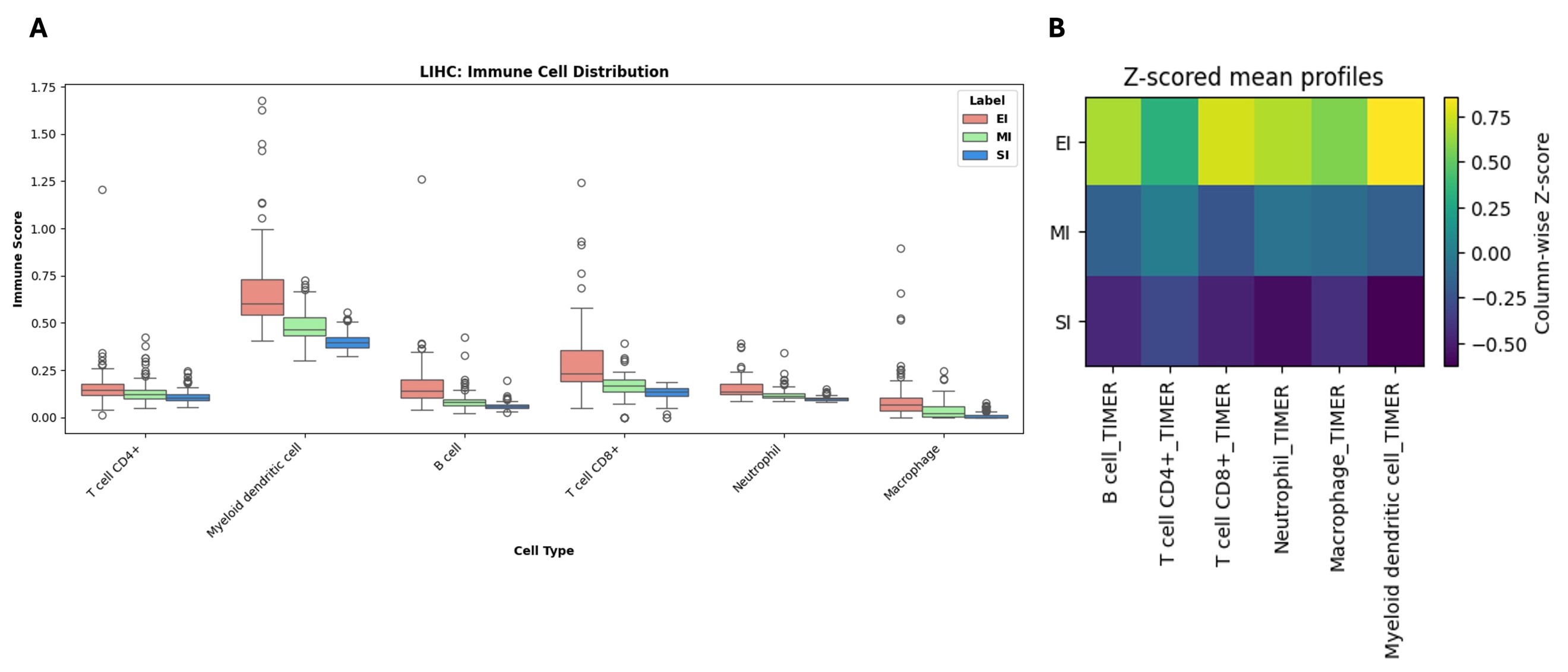

**Figure S10: (A)** Box plot depicting the distribution of each immune cell type by phenotype [EI (red), MI (green), SI (blue)] in LIHC. EI (n = 90), MI (n = 95), SI (n = 97). **(B)** Heatmap showing Z-scored mean profiles of six immune cell types [B cell, CD4+ T cell, CD8+ T cell, Neutrophil, Macrophage, and Myeloid dendritic cell] across three phenotypic groups: EI, MI, and SI in LIHC. Values are column-wise Z-scores, as indicated by the color bar.

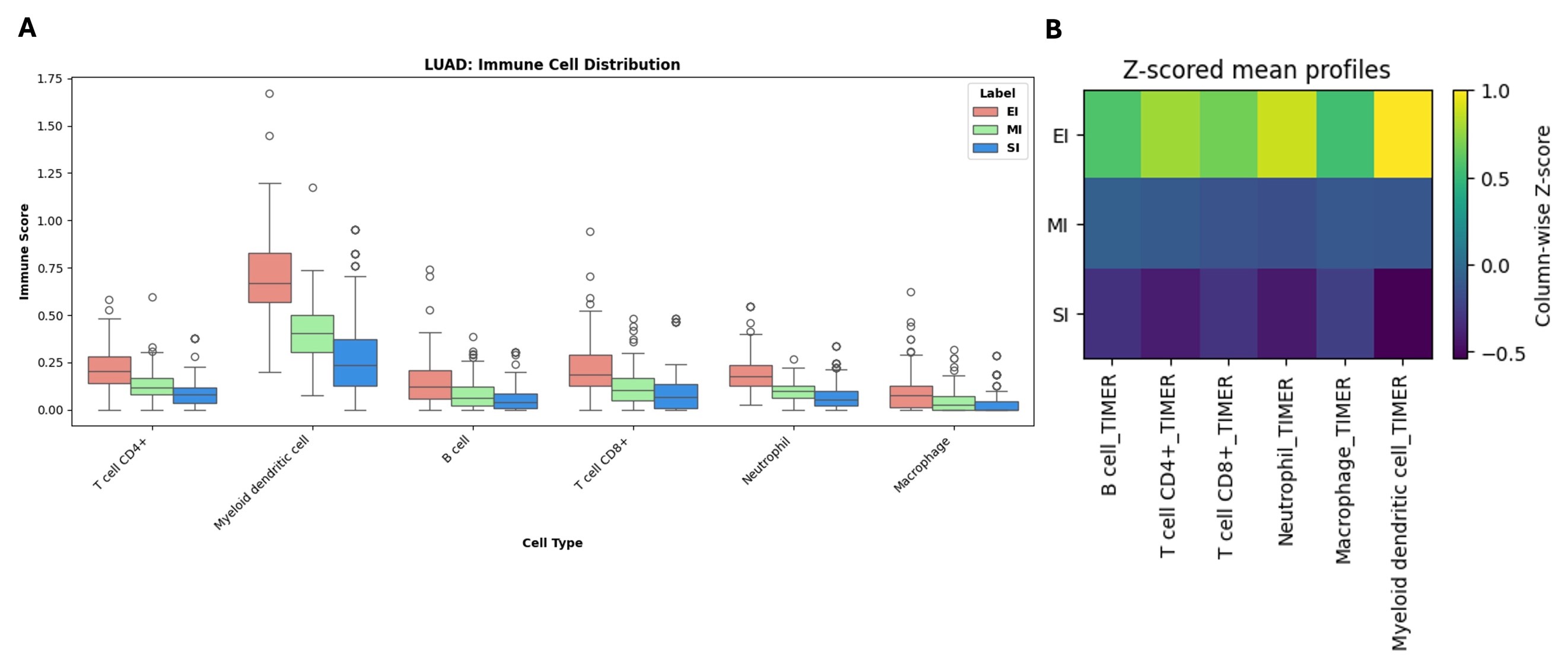

**Figure S11: (A)** Box plot depicting the distribution of each immune cell type by phenotype [EI (red), MI (green), SI (blue)] in LUAD. EI (n = 107), MI (n = 138), SI (n = 169). **(B)** Heatmap showing Z-scored mean profiles of six immune cell types [B cell, CD4+ T cell, CD8+ T cell, Neutrophil, Macrophage, and Myeloid dendritic cell] across three phenotypic groups: EI, MI, and SI in LUAD. Values are column-wise Z-scores, as indicated by the color bar.

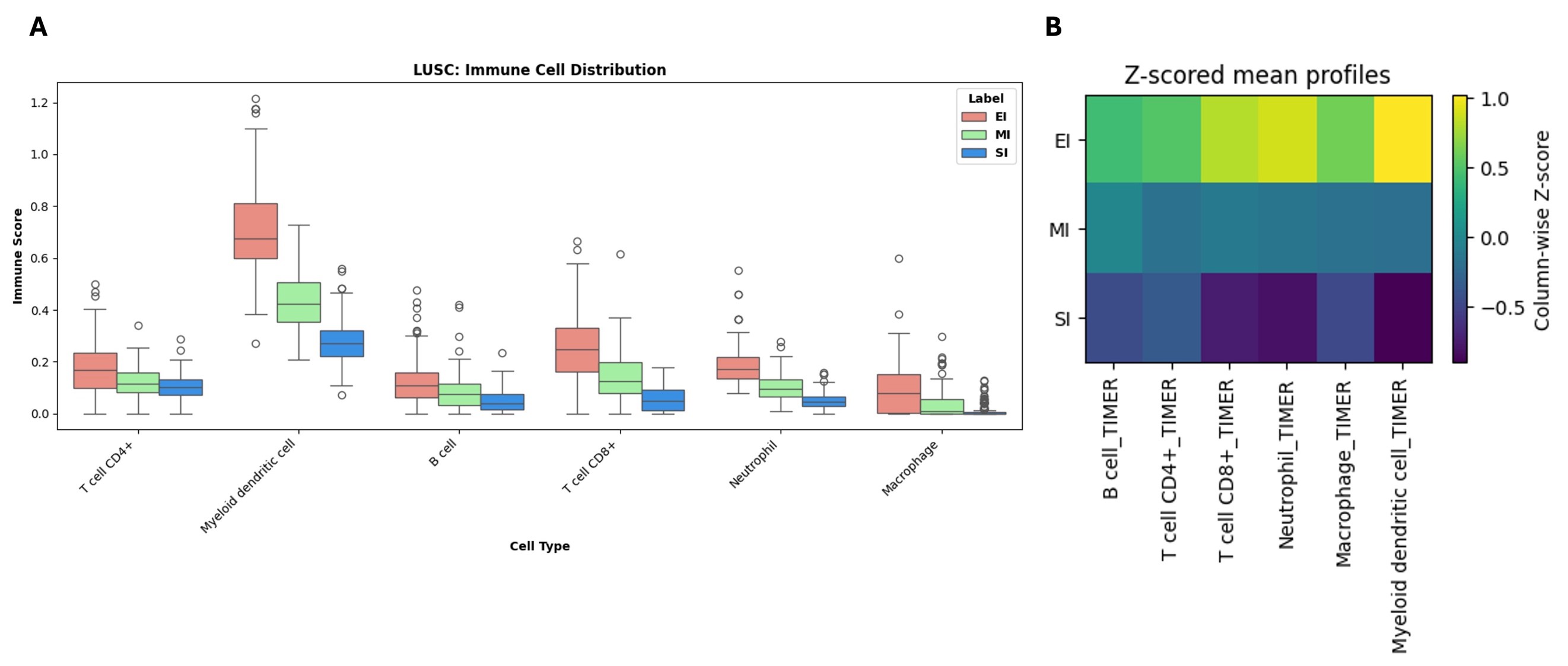

**Figure S12: (A)** Box plot depicting the distribution of each immune cell type by phenotype [EI (red), MI (green), SI (blue)] in LUSC. EI (n = 132), MI (n = 127), SI (n = 121). **(B)** Heatmap showing Z-scored mean profiles of six immune cell types [B cell, CD4+ T cell, CD8+ T cell, Neutrophil, Macrophage, and Myeloid dendritic cell] across three phenotypic groups: EI, MI, and SI in LUSC. Values are column-wise Z-scores, as indicated by the color bar.

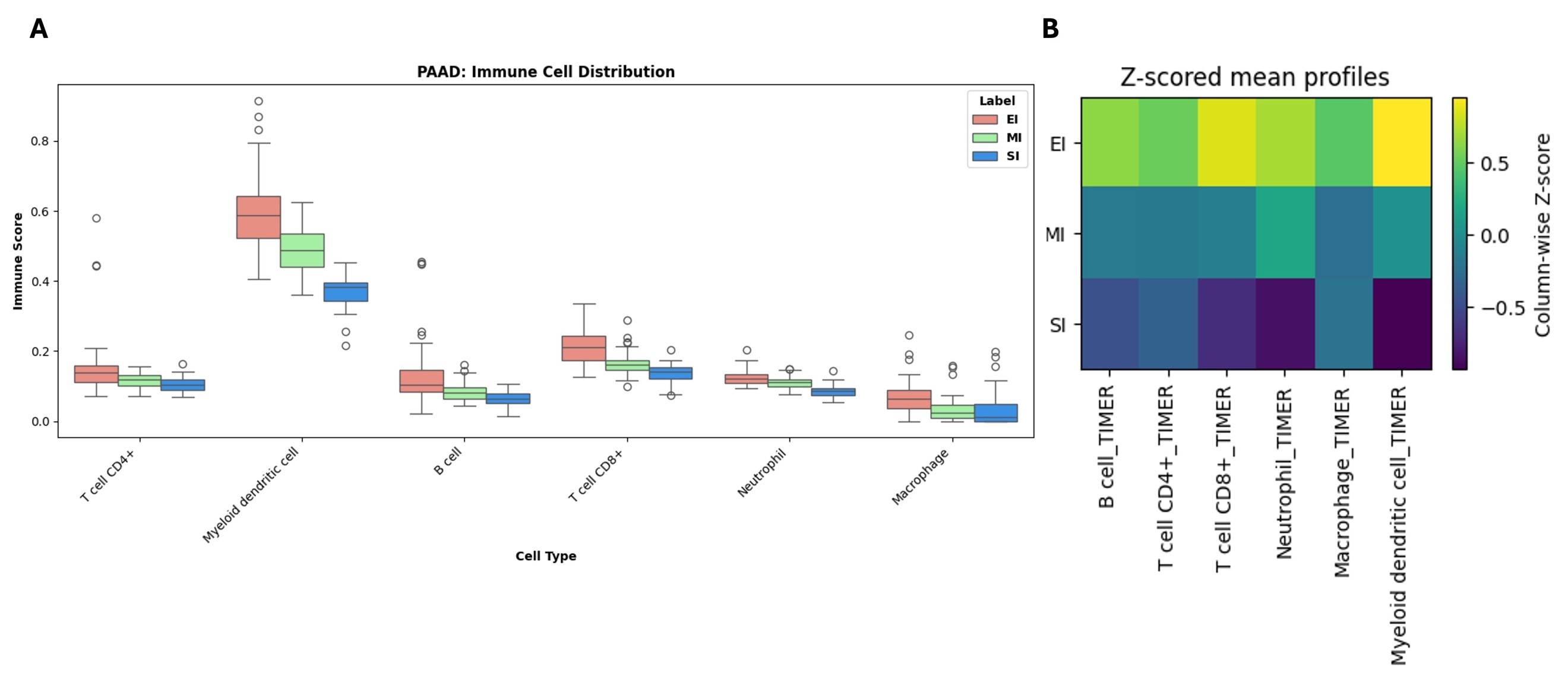

**Figure S13: (A)** Box plot depicting the distribution of each immune cell type by phenotype [EI (red), MI (green), SI (blue)] in PAAD. EI (n = 38), MI (n = 38), SI (n = 40). **(B)** Heatmap showing Z-scored mean profiles of six immune cell types [B cell, CD4+ T cell, CD8+ T cell, Neutrophil, Macrophage, and Myeloid dendritic cell] across three phenotypic groups: EI, MI, and SI in PAAD. Values are column-wise Z-scores, as indicated by the color bar.

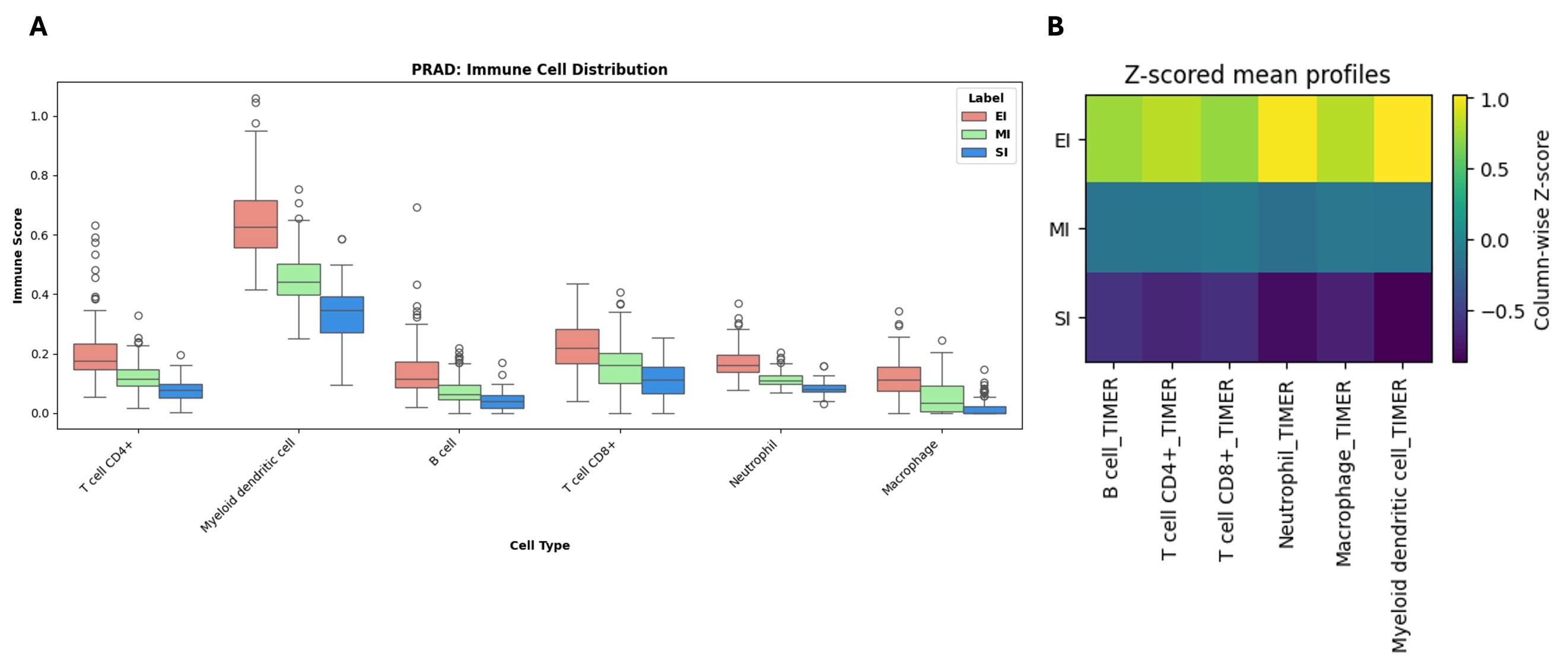

**Figure S14: (A)** Box plot depicting the distribution of each immune cell type by phenotype [EI (red), MI (green), SI (blue)] in PRAD. EI (n = 121), MI (n = 123), SI (n = 125). **(B)** Heatmap showing Z-scored mean profiles of six immune cell types [B cell, CD4+ T cell, CD8+ T cell, Neutrophil, Macrophage, and Myeloid dendritic cell] across three phenotypic groups: EI, MI, and SI in PRAD. Values are column-wise Z-scores, as indicated by the color bar.

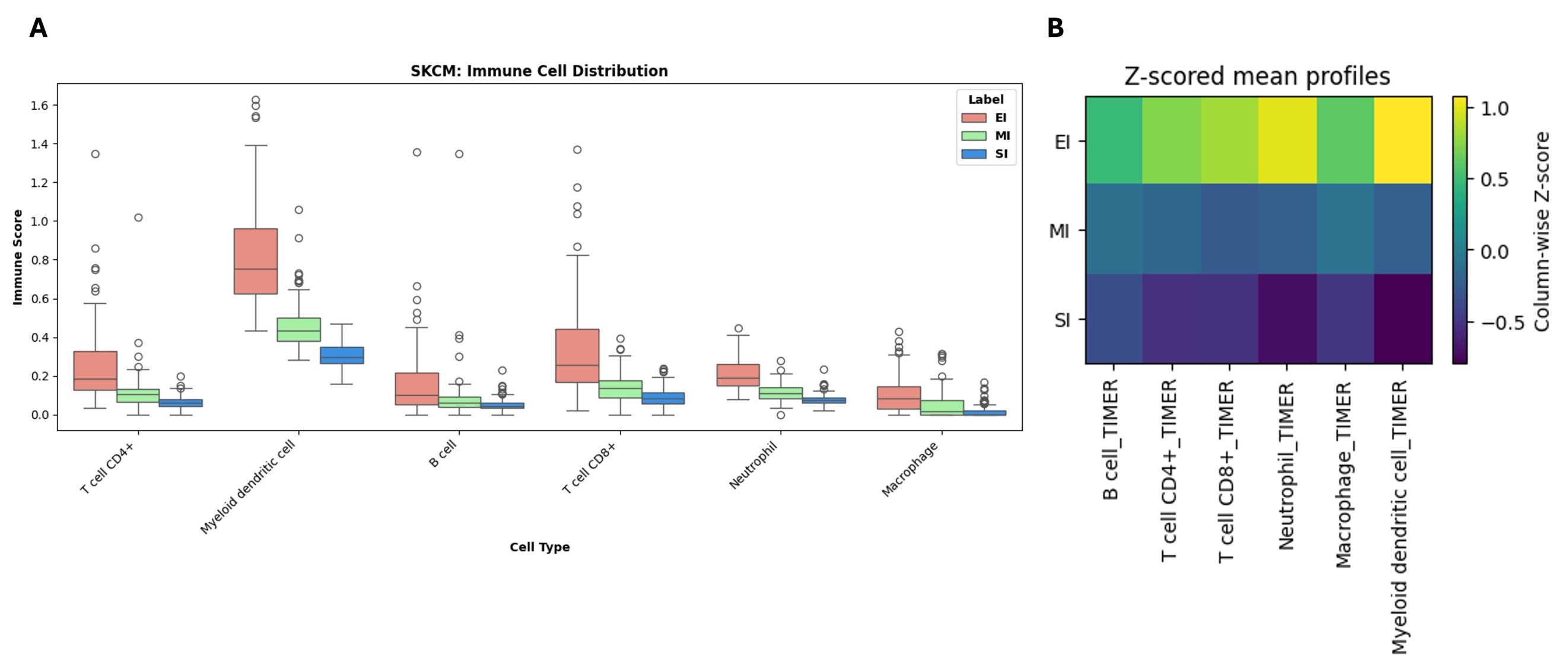

**Figure S15: (A)** Box plot depicting the distribution of each immune cell type by phenotype [EI (red), MI (green), SI (blue)] in SKCM. EI (n = 116), MI (n = 120), SI (n = 121). **(B)** Heatmap showing Z-scored mean profiles of six immune cell types [B cell, CD4+ T cell, CD8+ T cell, Neutrophil, Macrophage, and Myeloid dendritic cell] across three phenotypic groups: EI, MI, and SI in SKCM. Values are column-wise Z-scores, as indicated by the color bar.

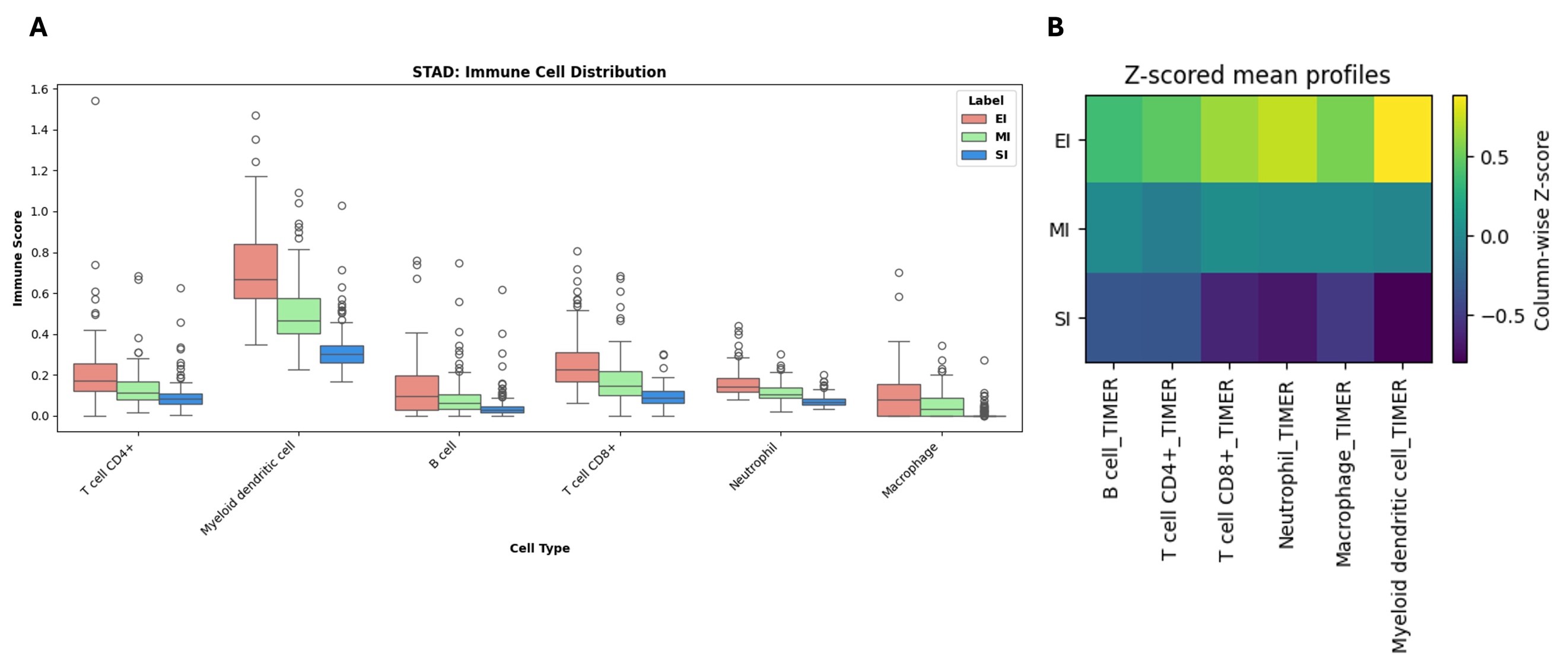

**Figure S16: (A)** Box plot depicting the distribution of each immune cell type by phenotype [EI (red), MI (green), SI (blue)] in STAD. EI (n = 105), MI (n = 113), SI (n = 112). **(B)** Heatmap showing Z-scored mean profiles of six immune cell types [B cell, CD4+ T cell, CD8+ T cell, Neutrophil, Macrophage, and Myeloid dendritic cell] across three phenotypic groups: EI, MI, and SI in STAD. Values are column-wise Z-scores, as indicated by the color bar.

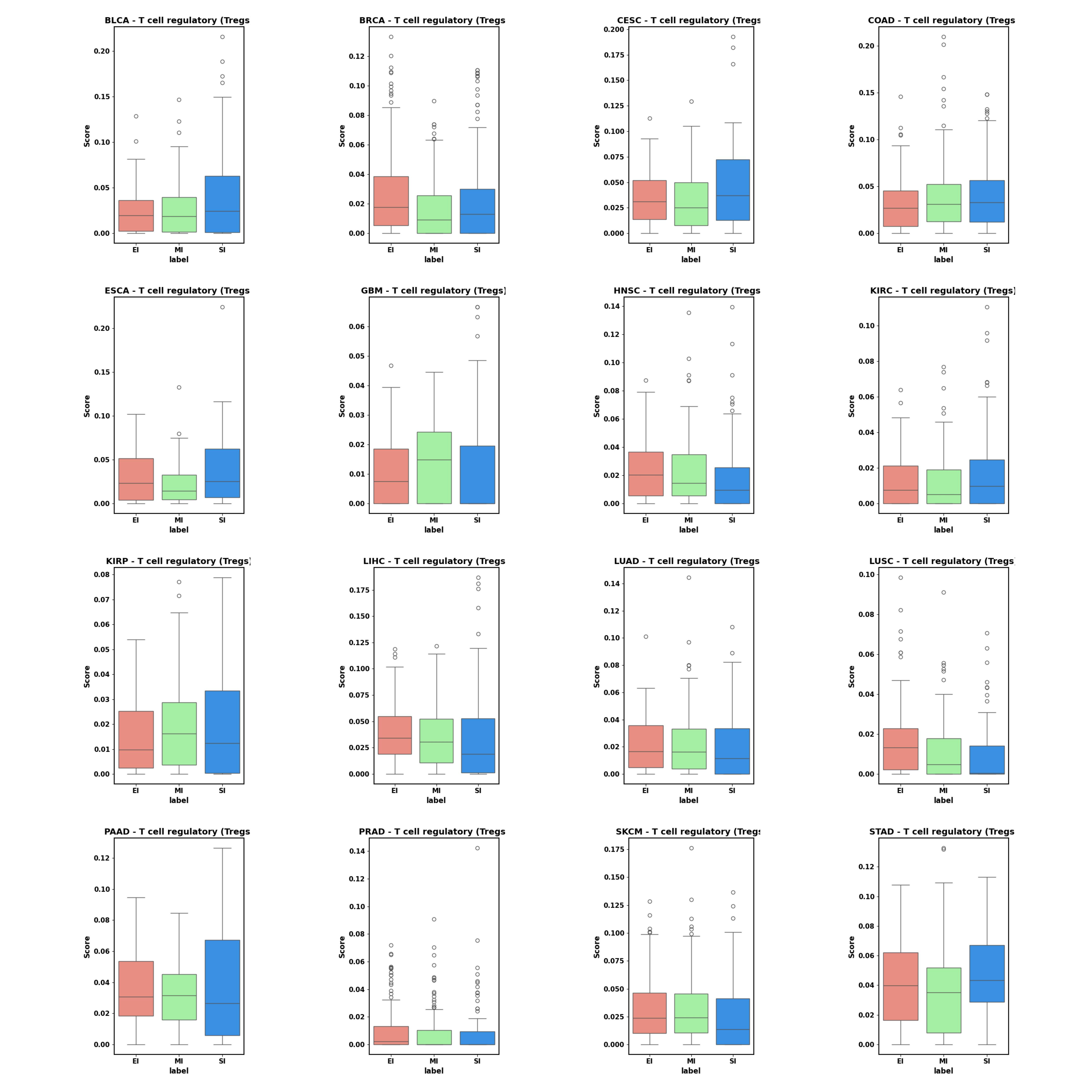

**Figure S17:** Box plot depicting the distribution of T regulatory cells by phenotype [EI (red), MI (green), SI (blue)] in different cancers. The sample number of each label in each cancer is: BLCA (EI:100,MI:108,SI:113), BRCA (EI:276,MI:282,SI:308), CESC (EI:62,MI:63,SI:62), COAD (EI:116,MI:142,SI:154), ESCA (EI:39,MI:38,SI:43), GBM (EI:66,MI:63,SI:64), HNSC (EI:111,MI:106,SI:117), KIRC (EI:88,MI:107,SI:124), KIRP (EI:68,MI:69,SI:74), LIHC (EI:90,MI:95,SI:97), LUAD (EI:107,MI:138,SI:169), LUSC (EI:132,MI:127,SI:121), PAAD (EI:38,MI:38,SI:40), PRAD (EI:121,MI:123,SI:125), SKCM (EI:116,MI:120,SI:121), STAD(EI:105,MI:113,SI:112).

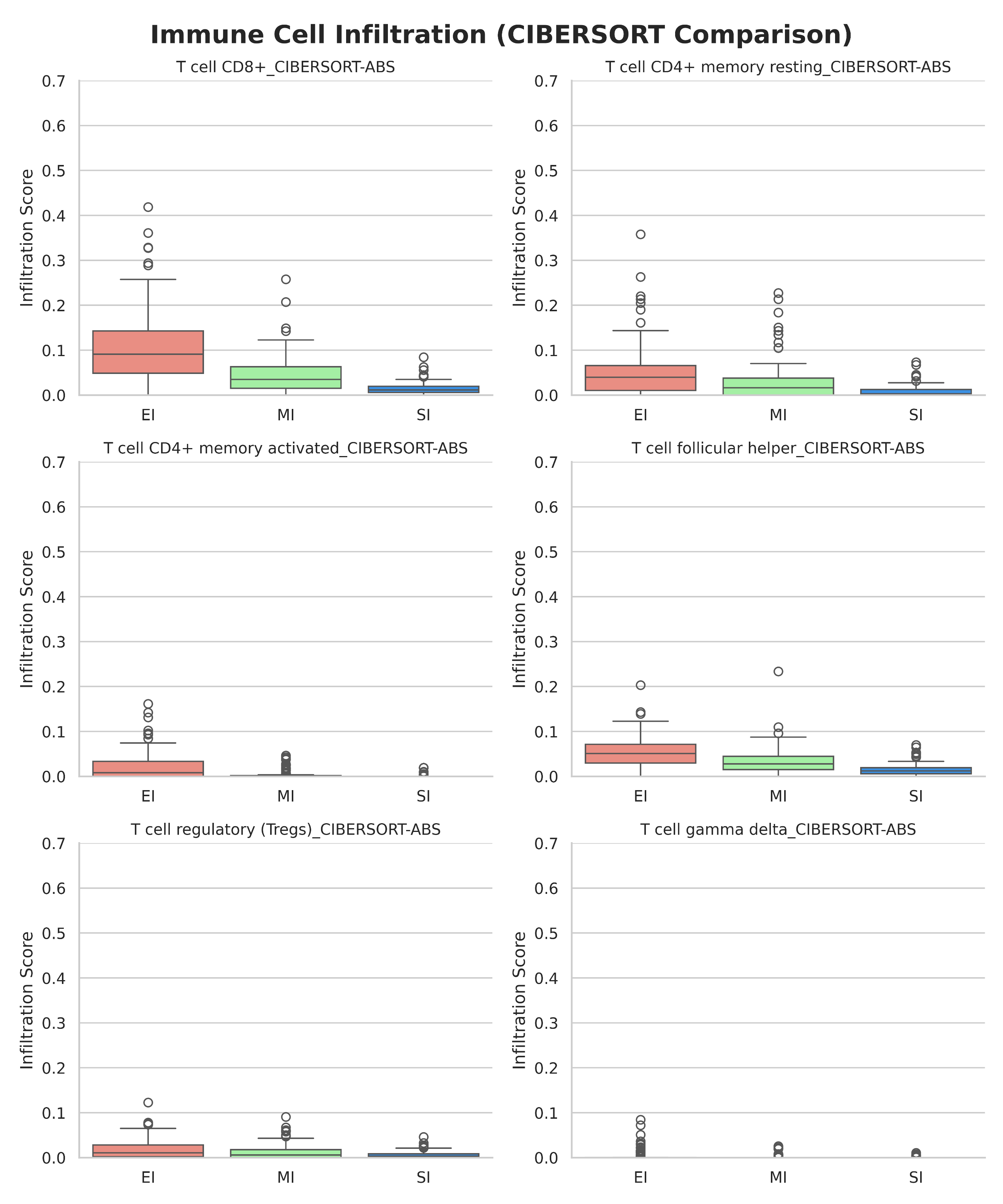

**Figure S18:** Box plot depicting the Immune cell infiltration profiles across [EI (red), MI (green), SI (blue)] groups in BLCA using the CIBERSORT method.

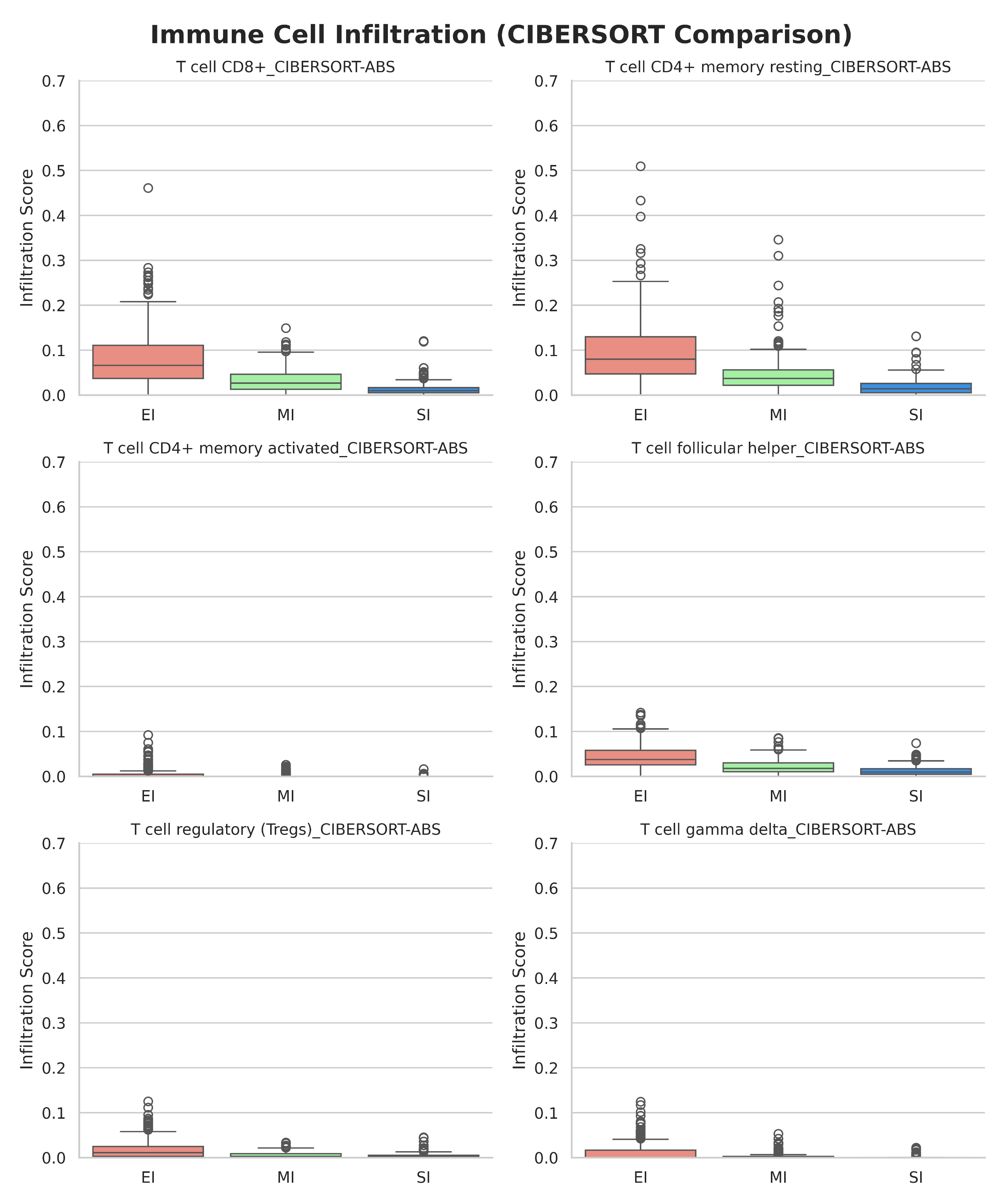

**Figure S19:** Box plot depicting the Immune cell infiltration profiles across [EI (red), MI (green), SI (blue)] groups in BRCA using the CIBERSORT method.

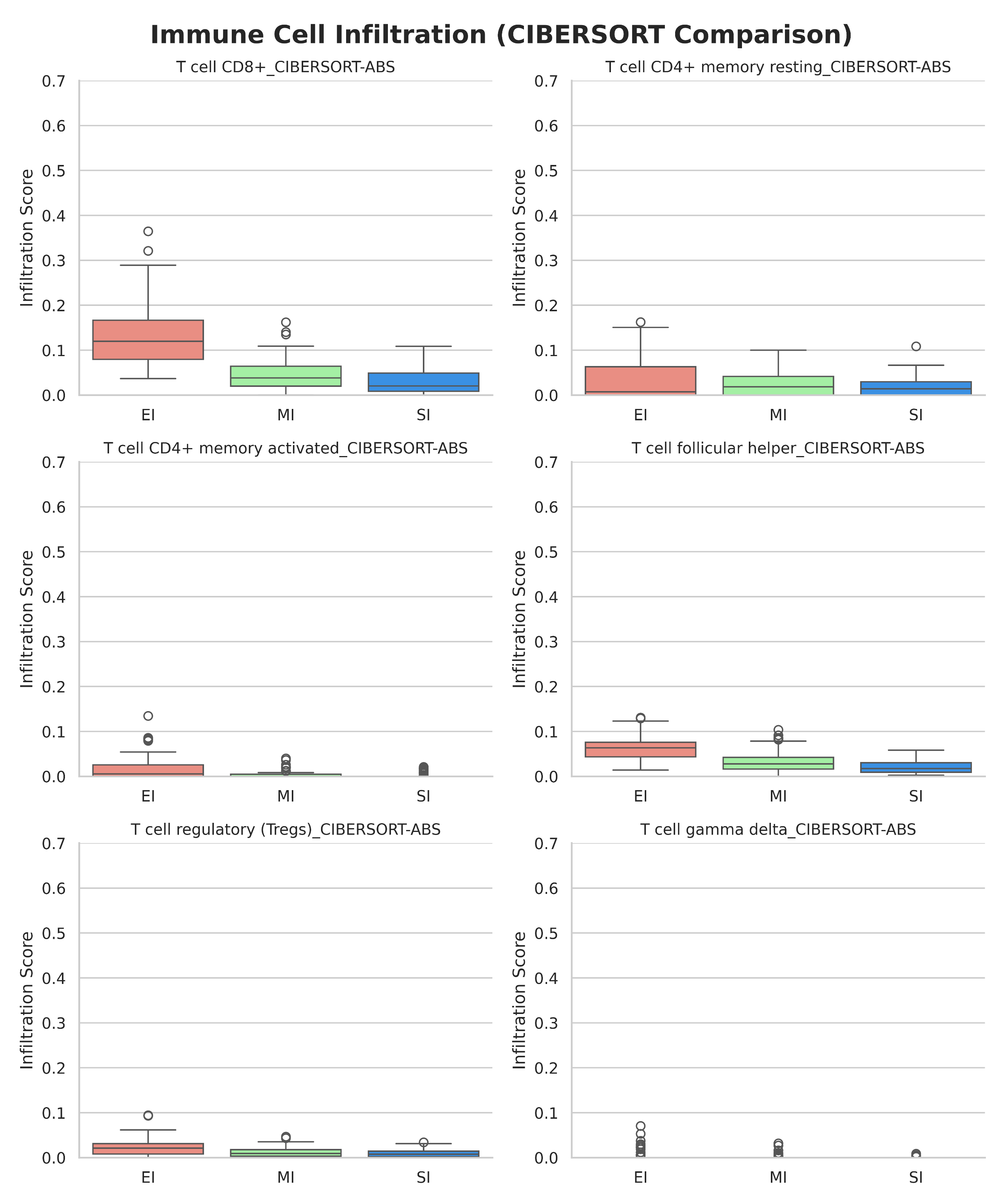

**Figure S20:** Box plot depicting the Immune cell infiltration profiles across [EI (red), MI (green), SI (blue)] groups in CESC using the CIBERSORT method.

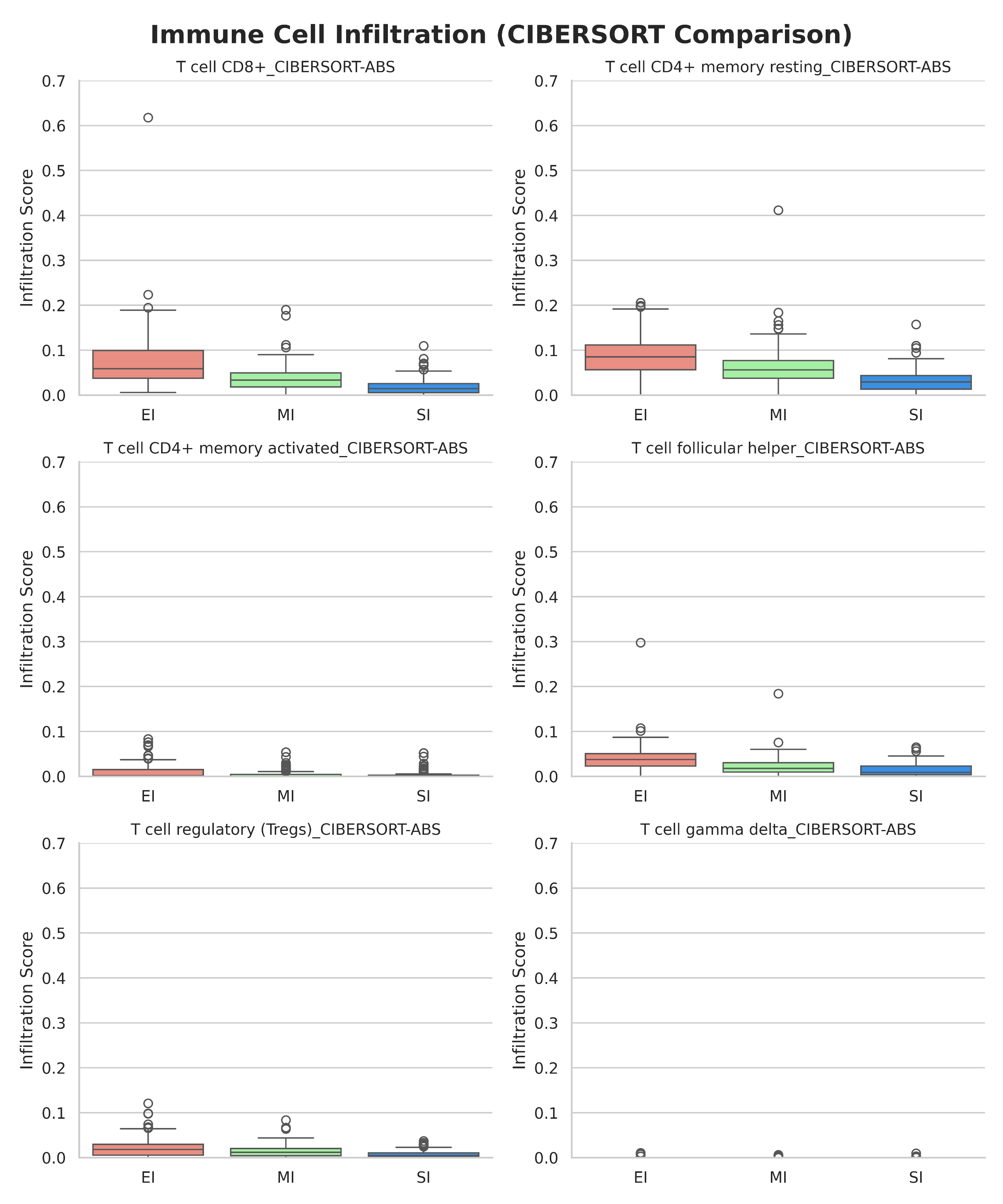

**Figure S21:** Box plot depicting the Immune cell infiltration profiles across [EI (red), MI (green), SI (blue)] groups in COAD using the CIBERSORT method.

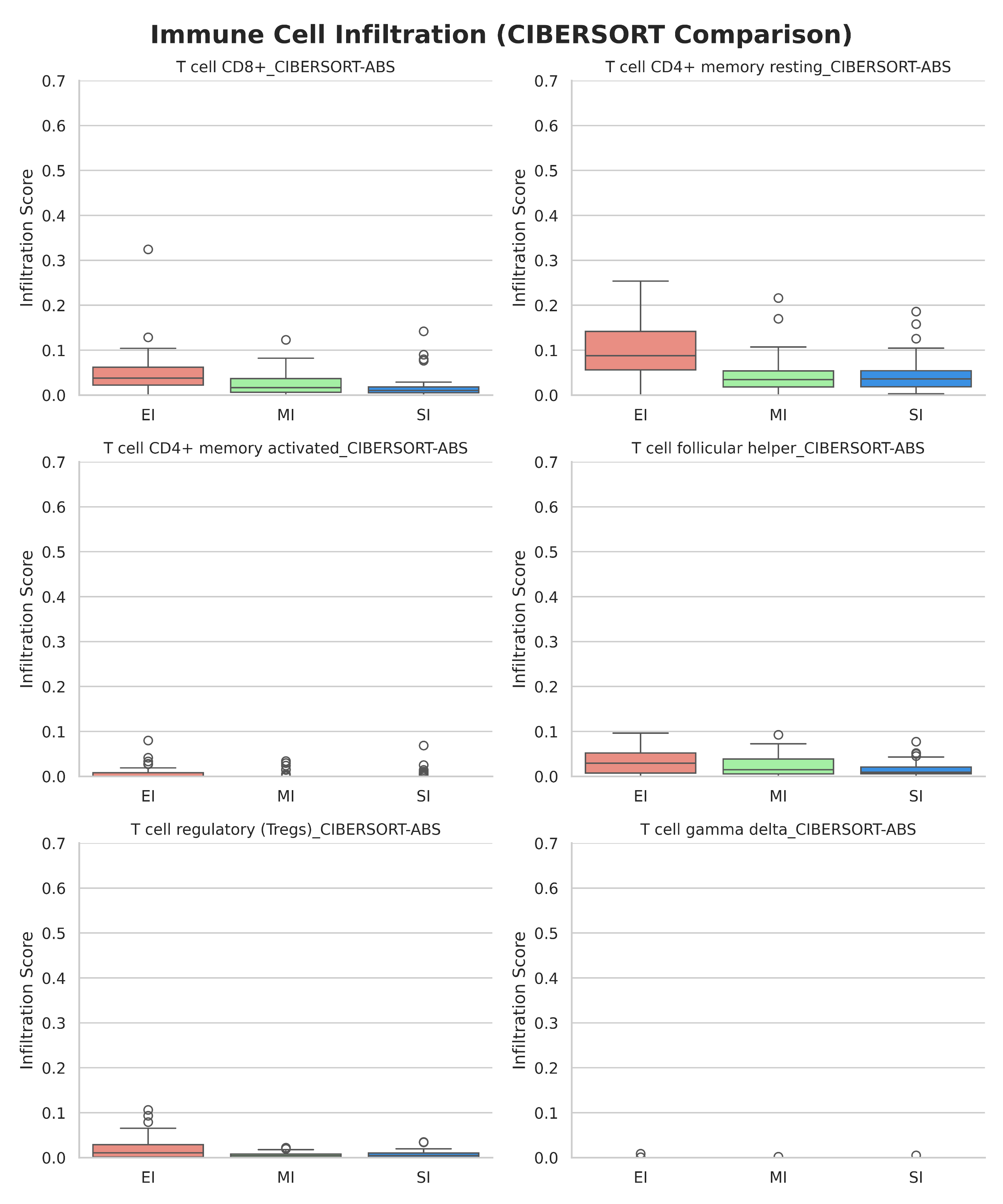

**Figure S22:** Box plot depicting the Immune cell infiltration profiles across [EI (red), MI (green), SI (blue)] groups in ESCA using the CIBERSORT method.

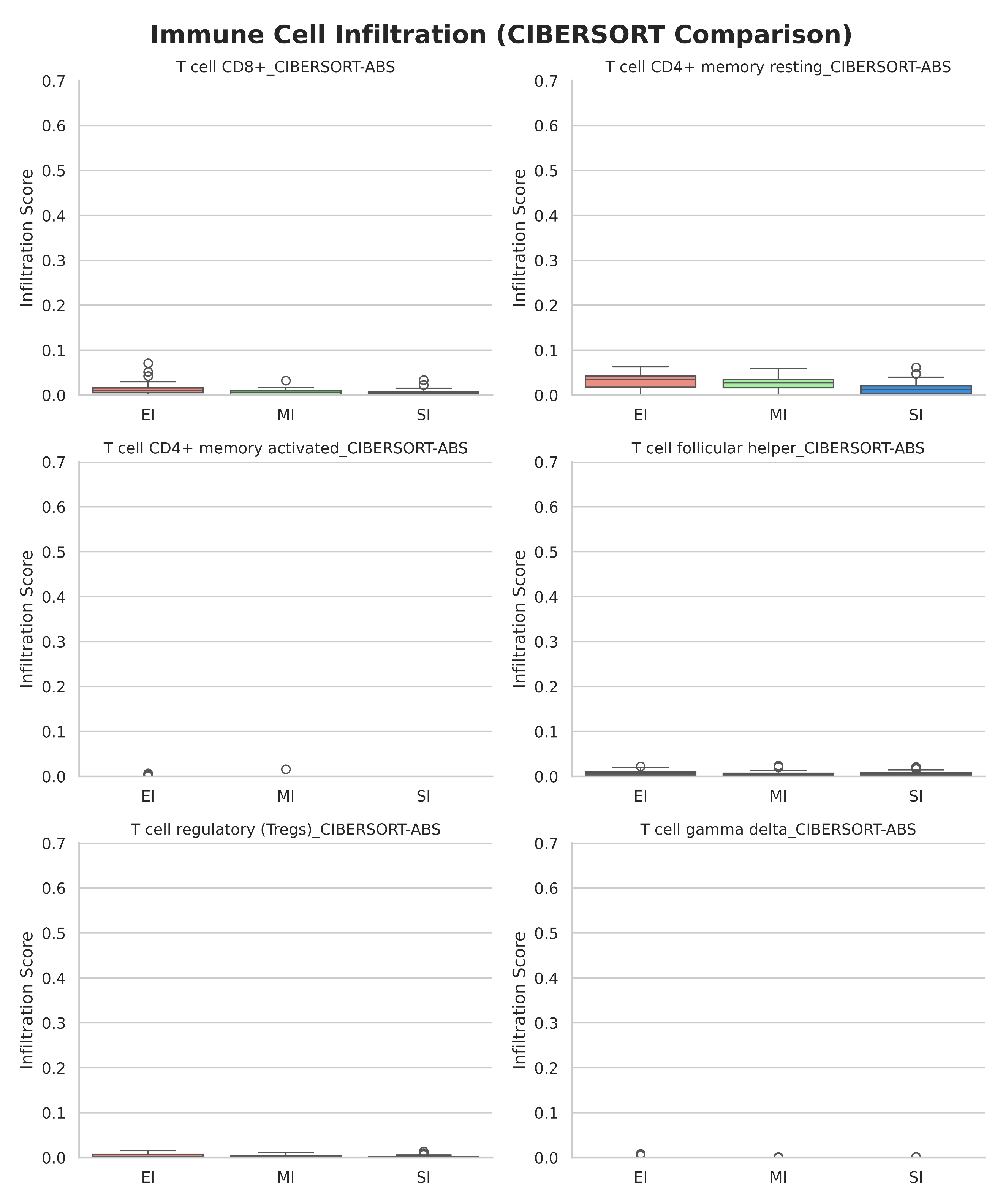

**Figure S23:** Box plot depicting the Immune cell infiltration profiles across [EI (red), MI (green), SI (blue)] groups in GBM using the CIBERSORT method.

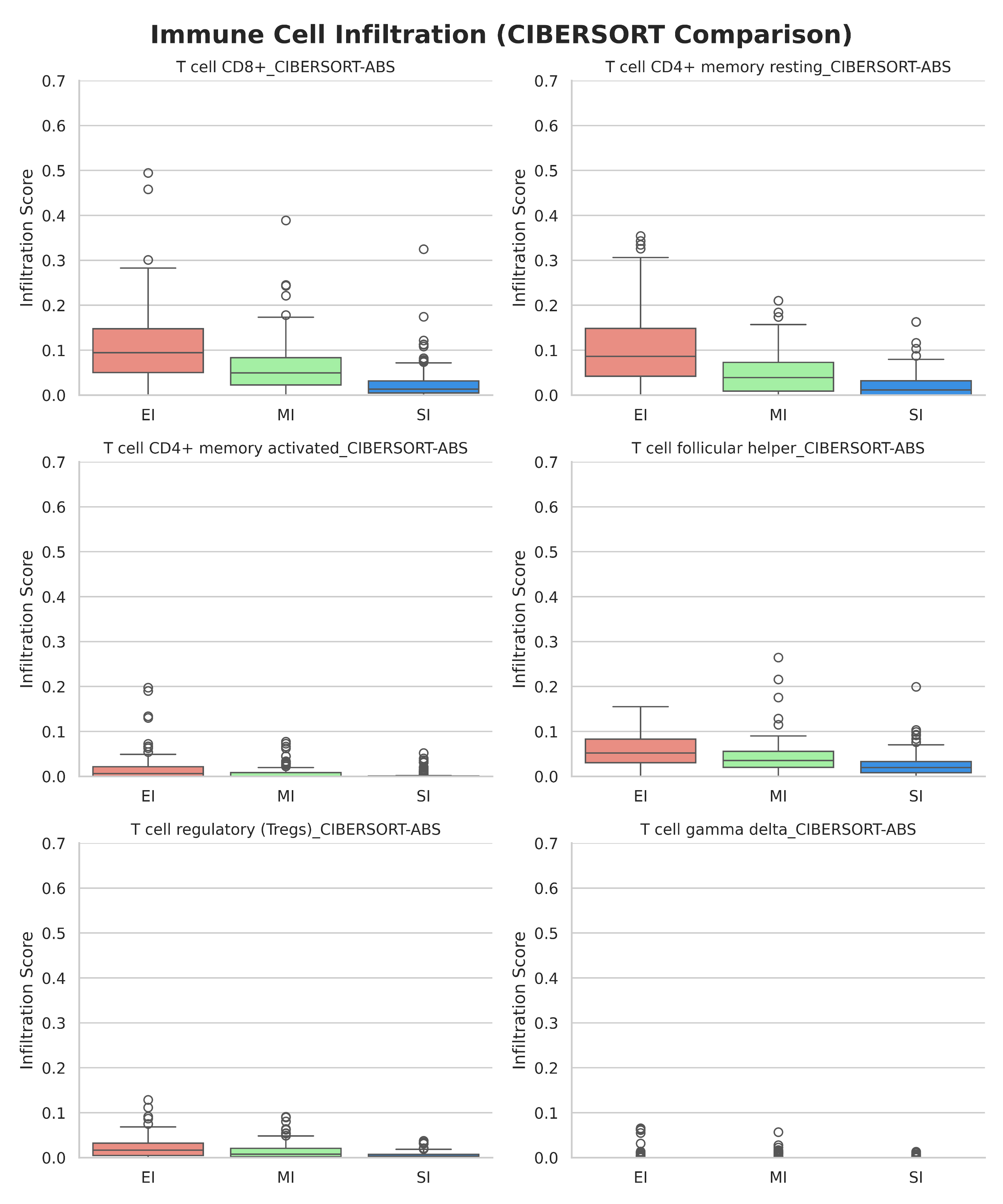

**Figure S24:** Box plot depicting the Immune cell infiltration profiles across [EI (red), MI (green), SI (blue)] groups in HNSC using the CIBERSORT method.

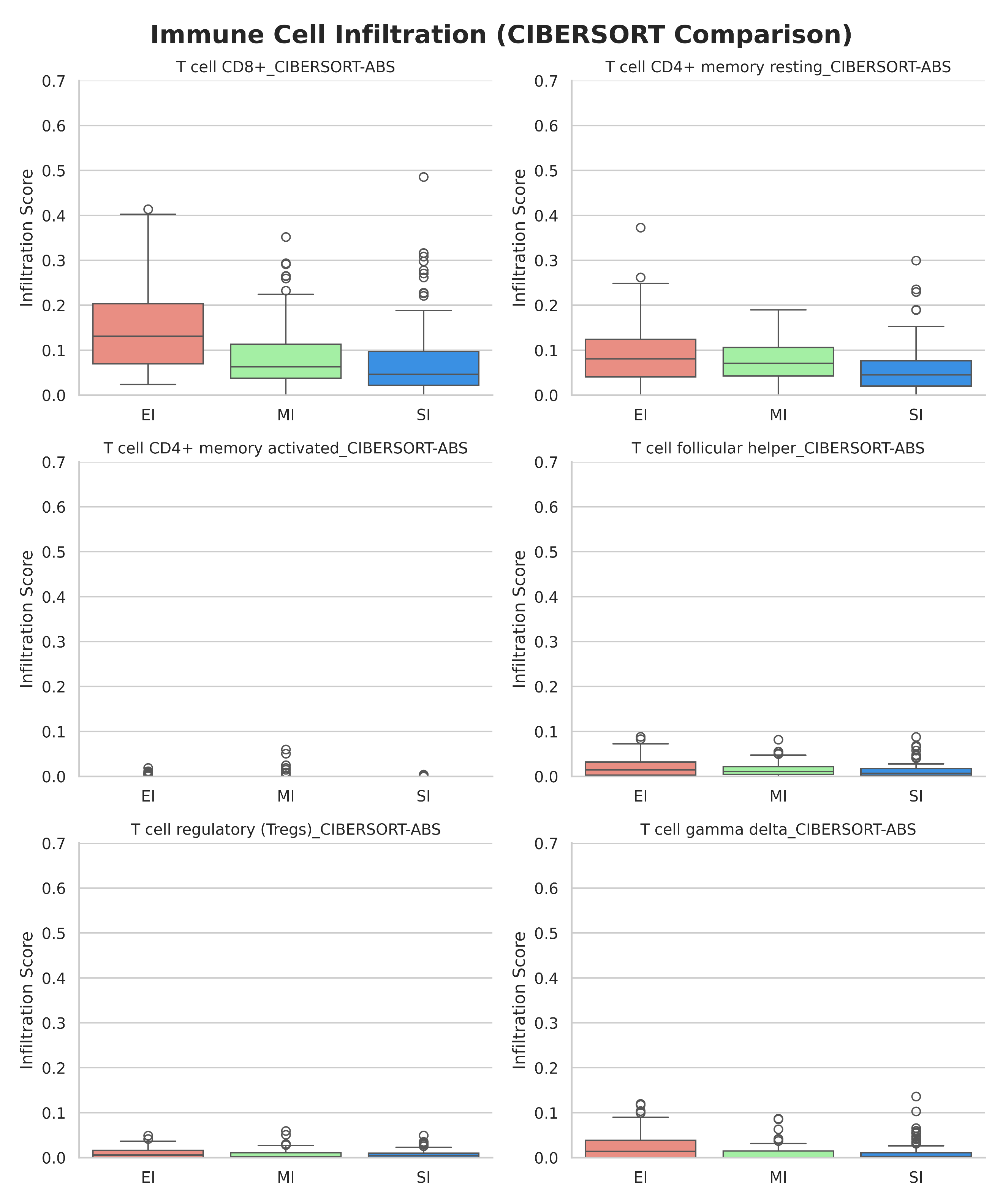

**Figure S25:** Box plot depicting the Immune cell infiltration profiles across [EI (red), MI (green), SI (blue)] groups in KIRC using the CIBERSORT method.

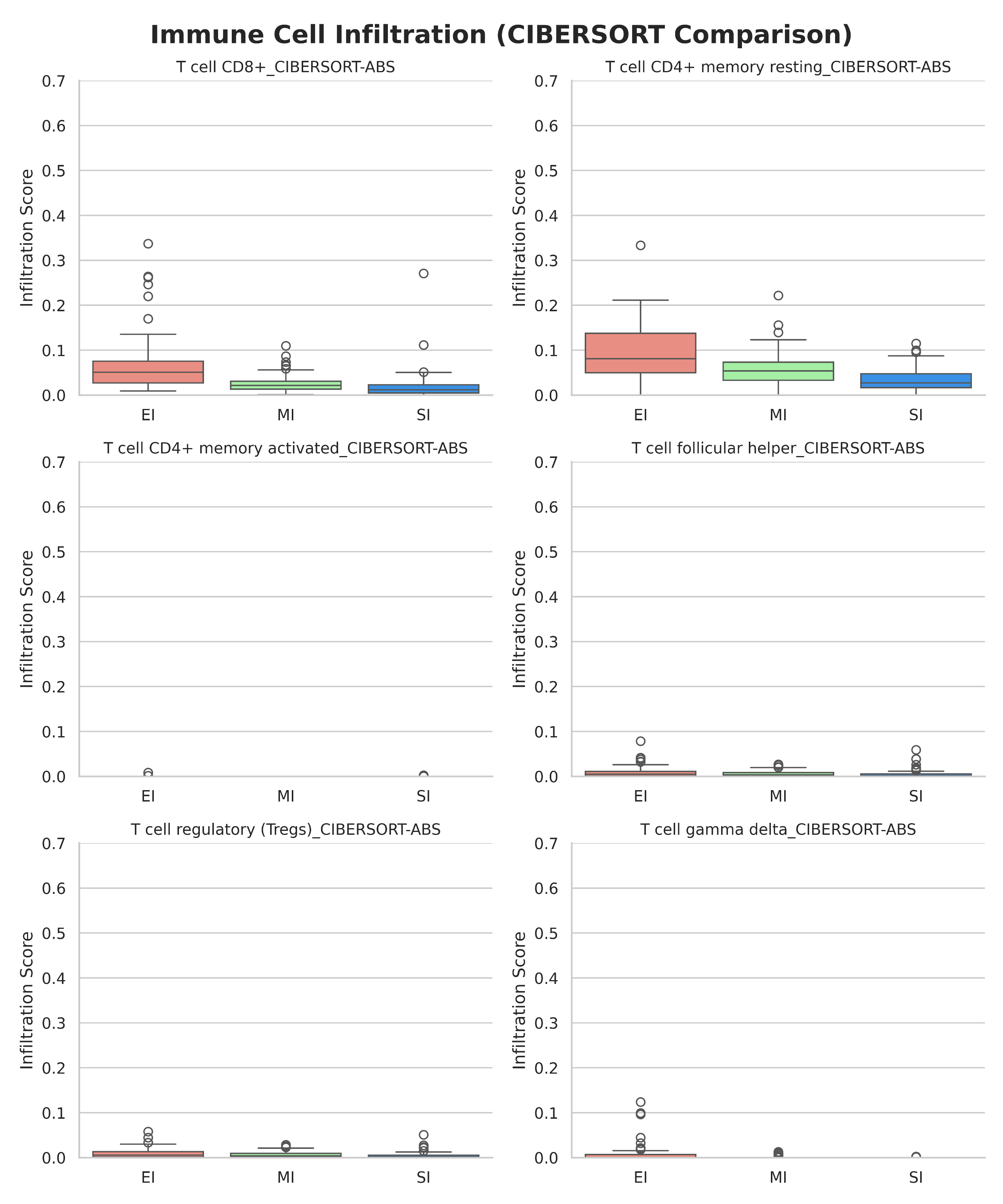

**Figure S26:** Box plot depicting the Immune cell infiltration profiles across [EI (red), MI (green), SI (blue)] groups in KIRP using the CIBERSORT method.

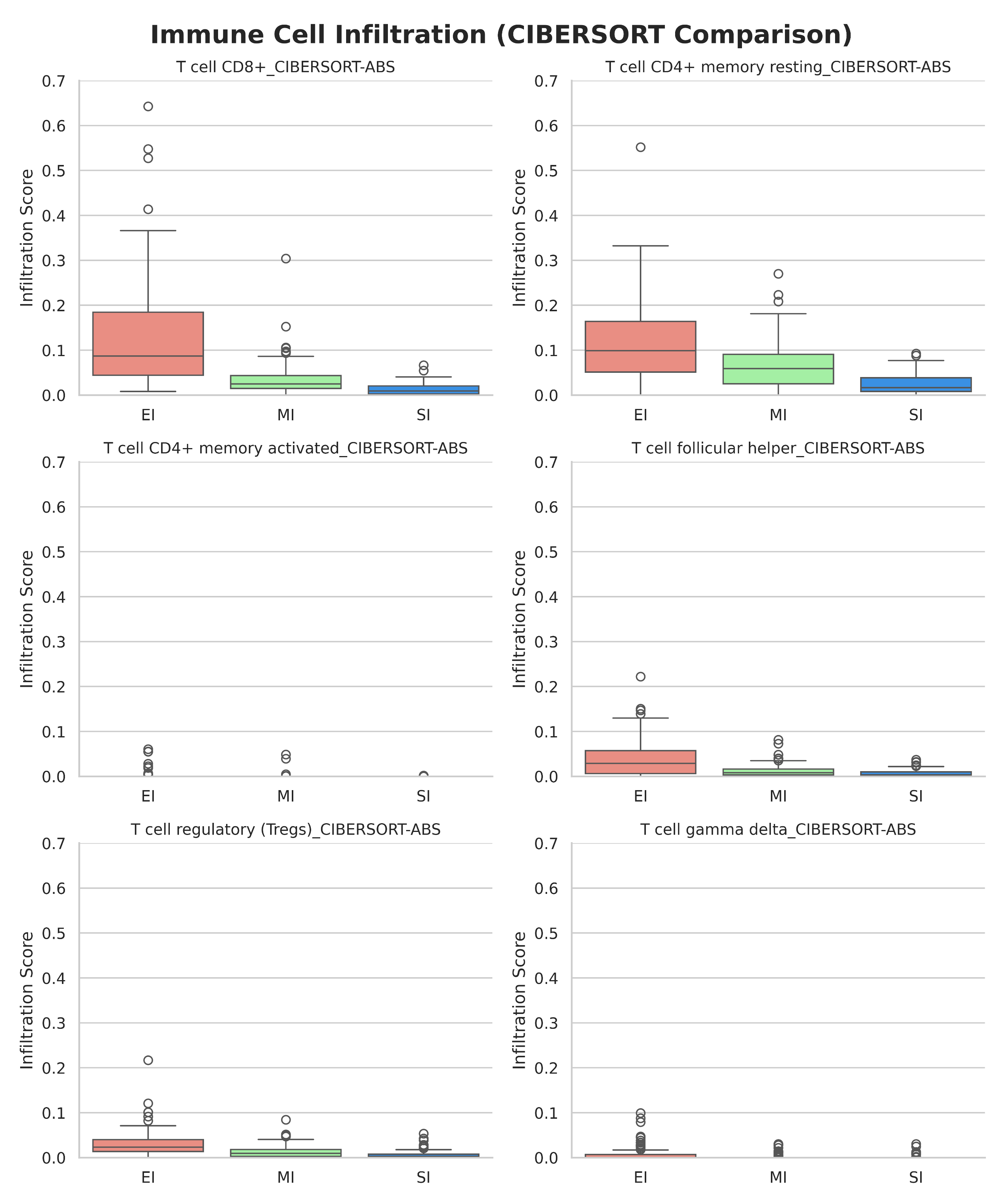

**Figure S27:** Box plot depicting the Immune cell infiltration profiles across [EI (red), MI (green), SI (blue)] groups in LIHC using the CIBERSORT method.

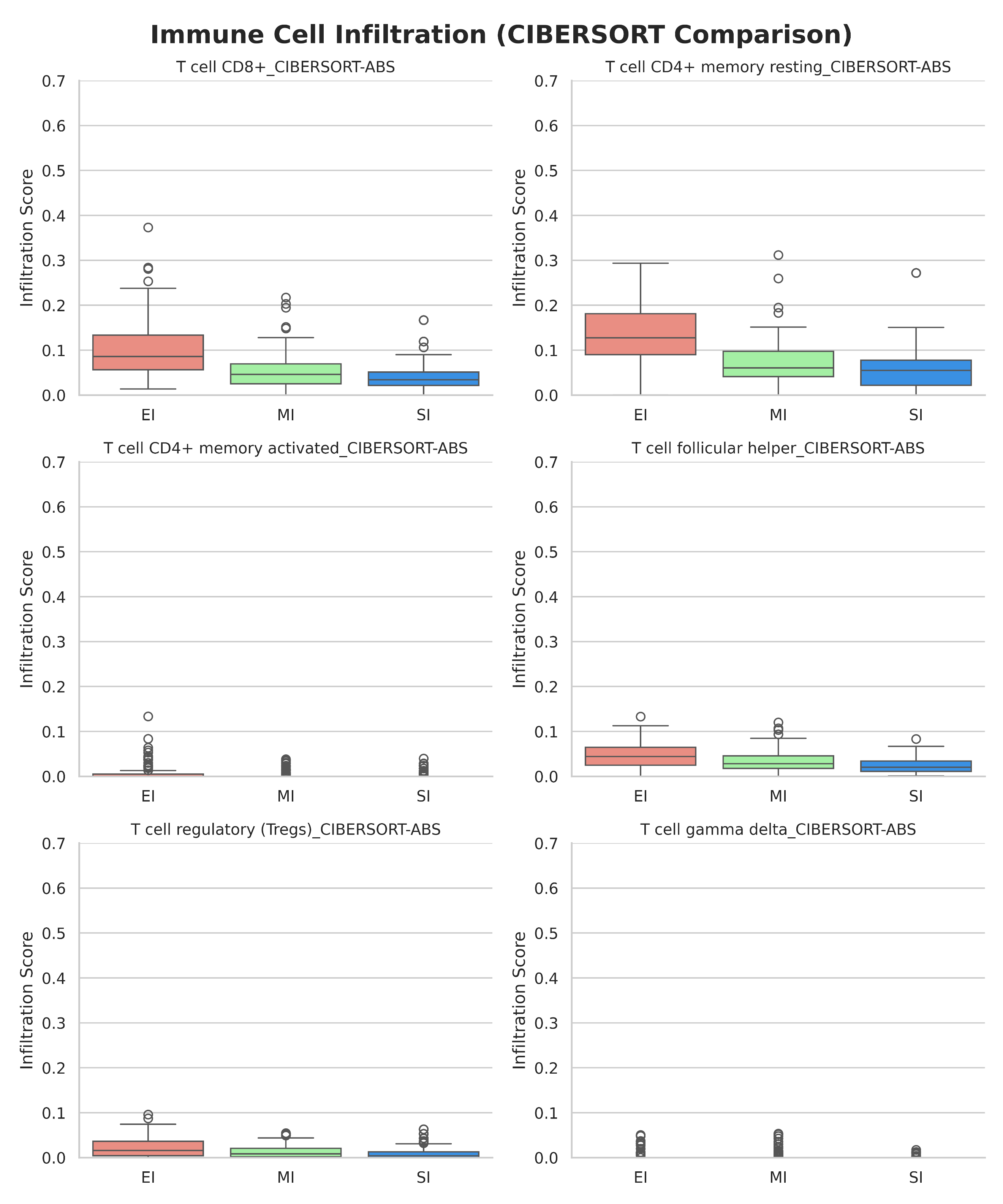

**Figure S28:** Box plot depicting the Immune cell infiltration profiles across [EI (red), MI (green), SI (blue)] groups in LUAD using the CIBERSORT method.

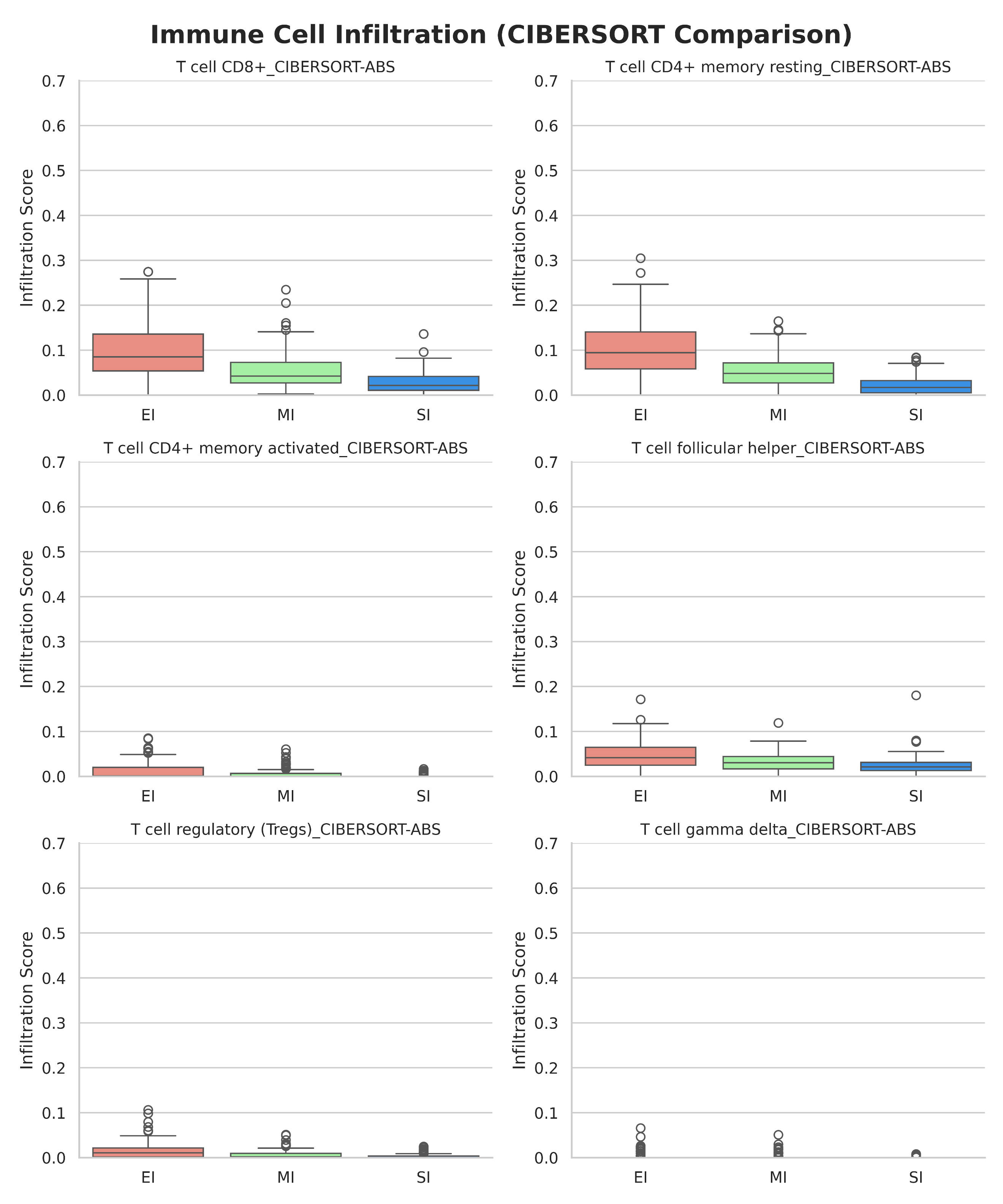

**Figure S29:** Box plot depicting the Immune cell infiltration profiles across [EI (red), MI (green), SI (blue)] groups in LUSC using the CIBERSORT method.

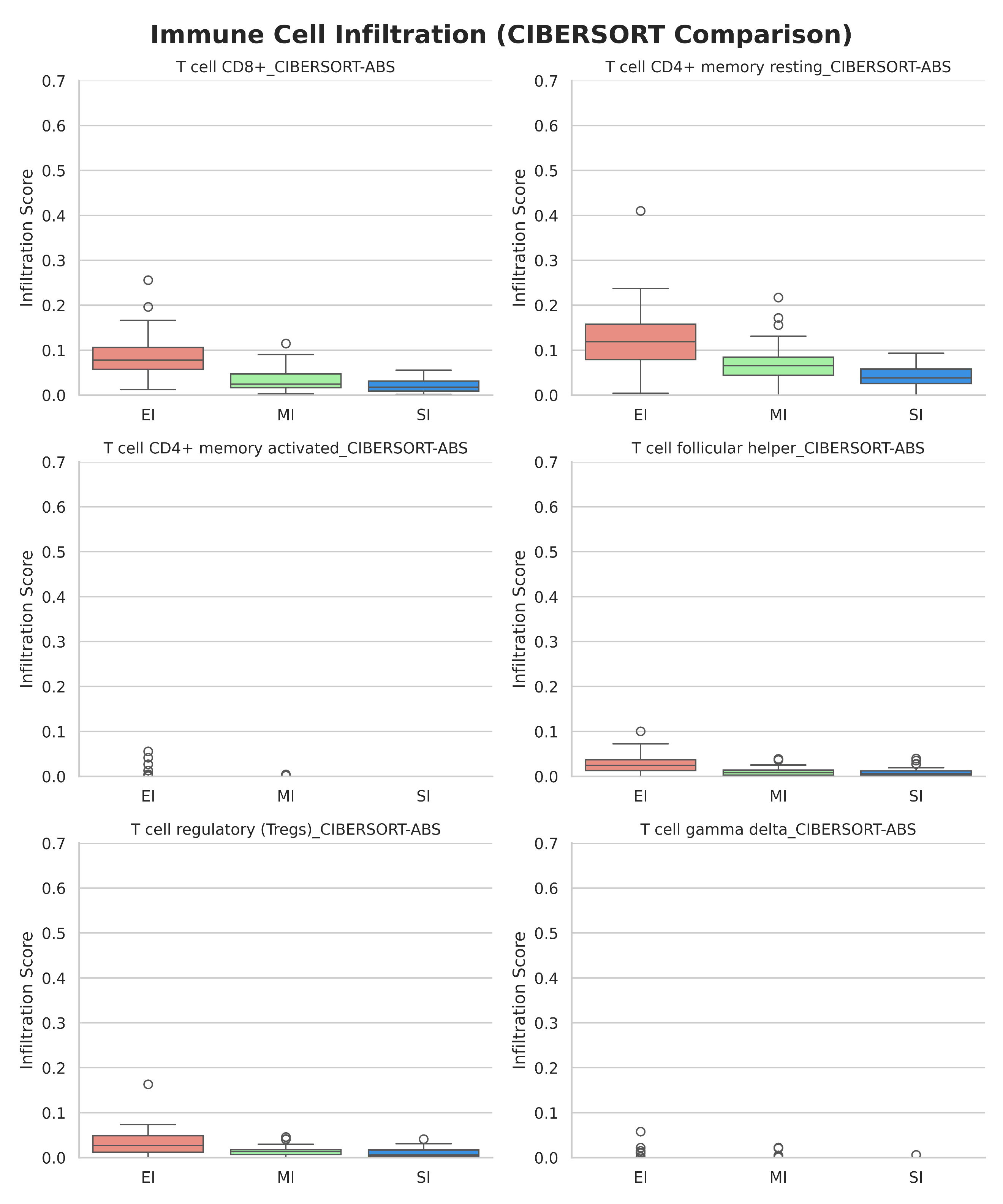

**Figure S30:** Box plot depicting the Immune cell infiltration profiles across [EI (red), MI (green), SI (blue)] groups in PAAD using the CIBERSORT method.

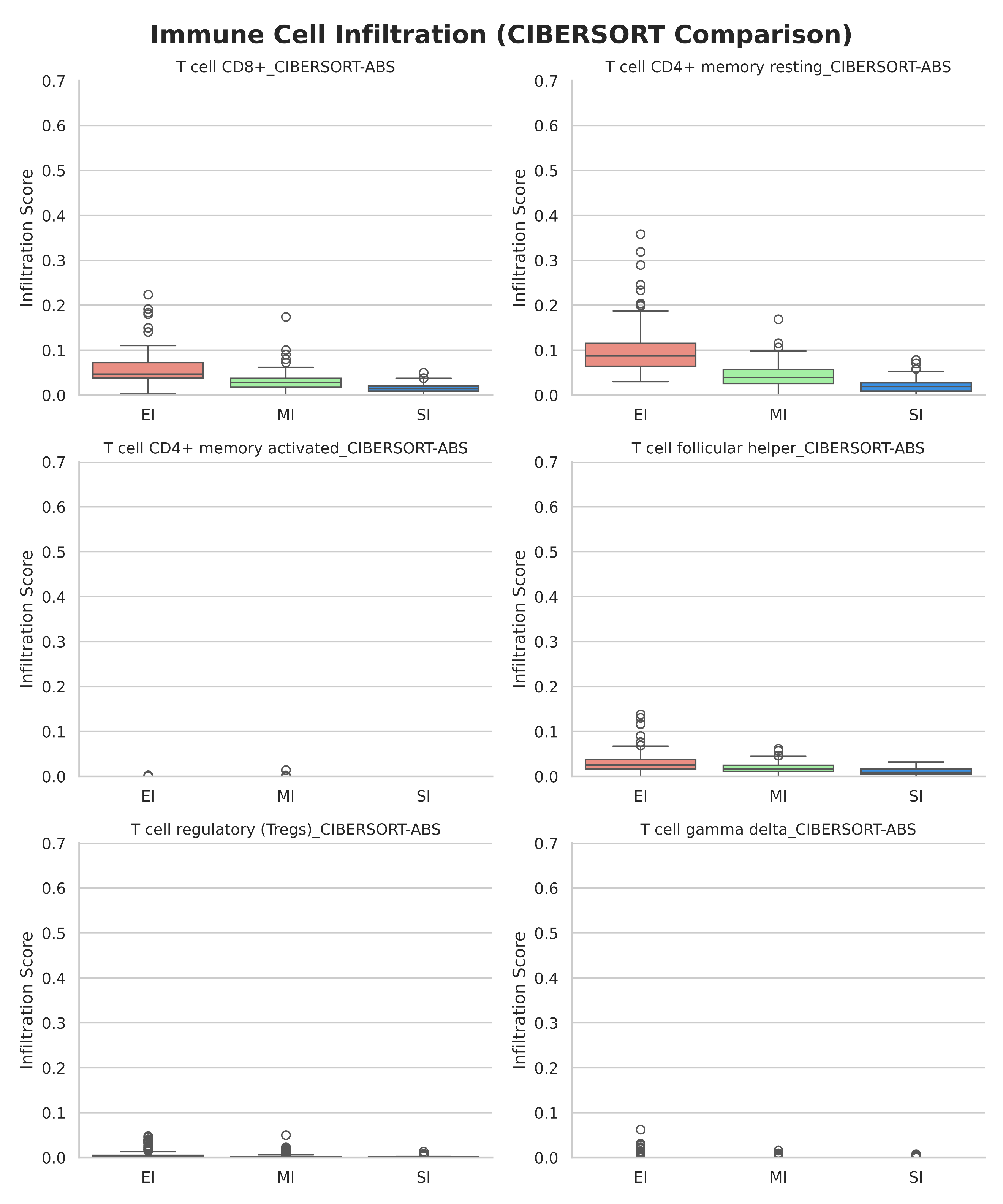

**Figure S31:** Box plot depicting the Immune cell infiltration profiles across [EI (red), MI (green), SI (blue)] groups in PRAD using the CIBERSORT method.

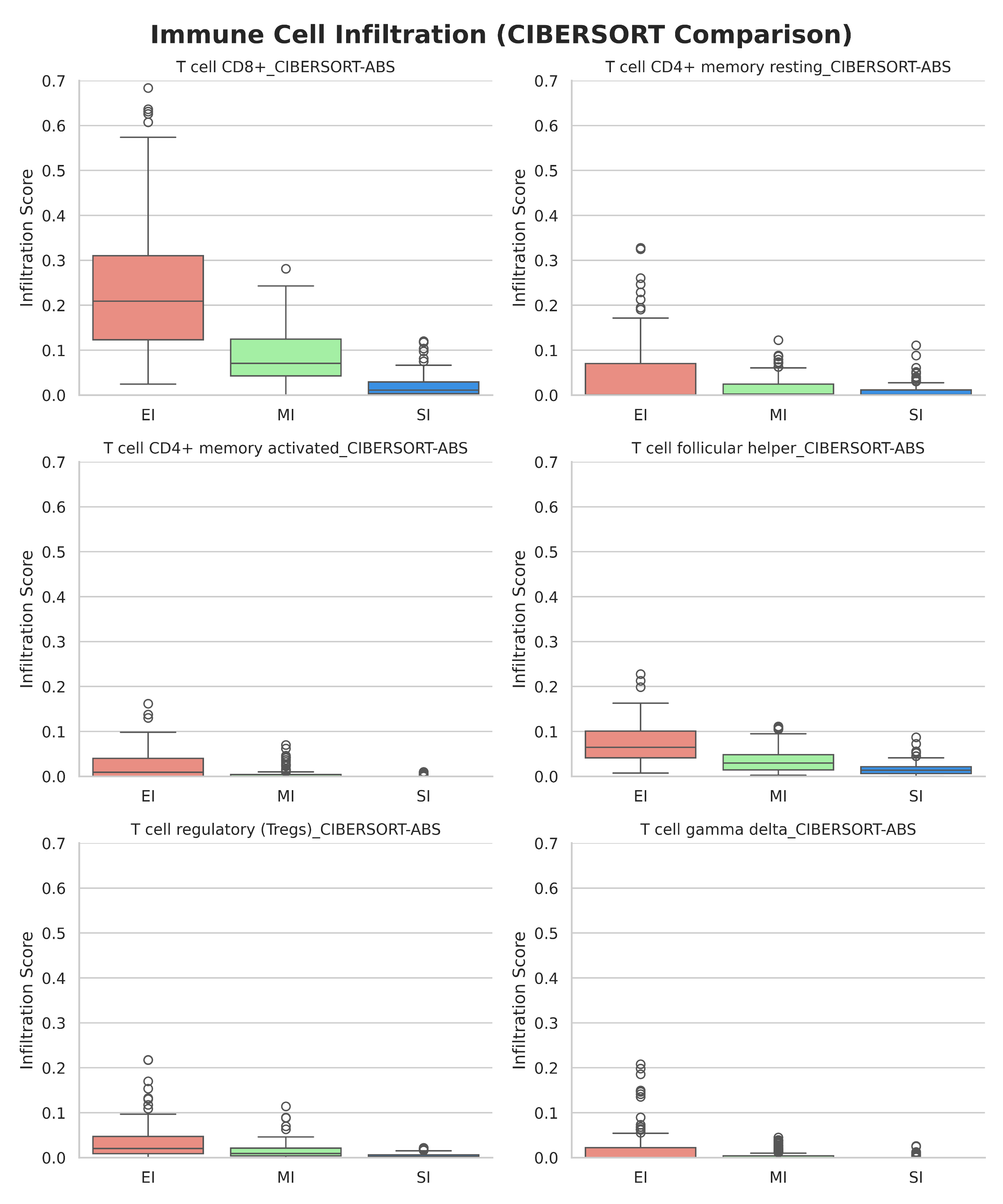

**Figure S32:** Box plot depicting the Immune cell infiltration profiles across [EI (red), MI (green), SI (blue)] groups in SKCM using the CIBERSORT method.

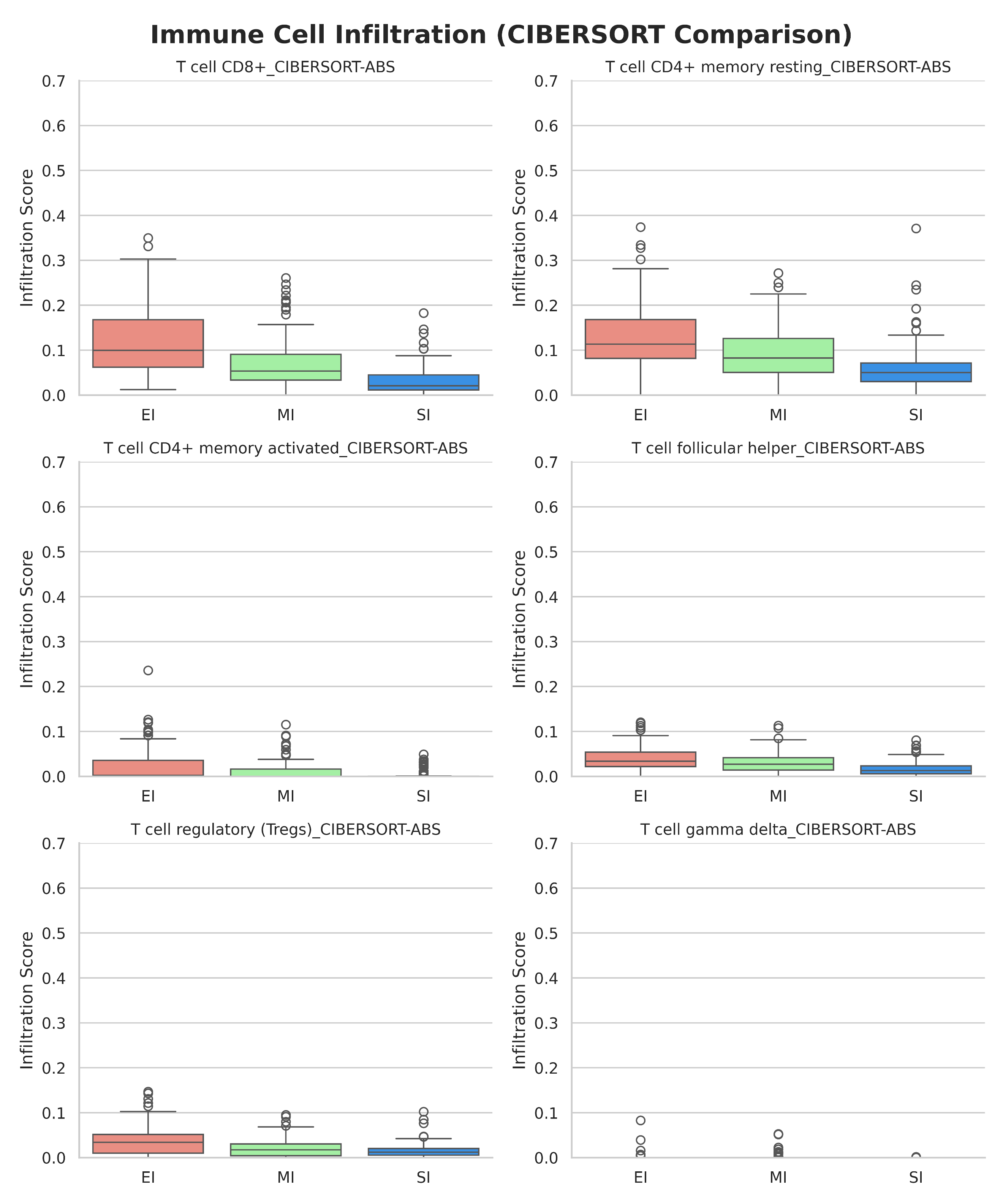

**Figure S33:** Box plot depicting the Immune cell infiltration profiles across [EI (red), MI (green), SI (blue)] groups in STAD using the CIBERSORT method.

**Supplementary Table S1: Standard Deviation of Bar plots of Pan-cancer Immune Cell Infiltration**

| **Label** | **B cell** | **T cell CD4+** | **T cell CD8+** | **Neutrophil** | **Macrophage** | **Myeloid dendritic cell** |
| --- | --- | --- | --- | --- | --- | --- |
| EI | 0.146209 | 0.147198 | 0.176784 | 0.088923 | 0.111524 | 0.239878 |
| MI | 0.091834 | 0.074632 | 0.099726 | 0.048583 | 0.073522 | 0.13118 |
| SI | 0.049853 | 0.055349 | 0.092175 | 0.054315 | 0.053809 | 0.153708 |

**Supplementary Table S2: P-values of Statistical tests performed between tumor immune-subtype (pan-cancer) for different immune cells**

| **Comparison** | **Feature** | ∆**-Mean** | **t-test p^*^** | **MW p** ^#^ |
| --- | --- | --- | --- | --- |
| EI vs MI | B cell | 0.057375 | 1.700106e-40 | 4.484589e-60 |
| EI vs MI | T cell CD4+ | 0.072010 | 2.792299e-66 | 4.132741e-100 |
| EI vs MI | T cell CD8+ | 0.111115 | 1.145282e-100 | 3.477579e-126 |
| EI vs MI | Neutrophil | 0.079276 | 5.054417e-187 | 1.991769e-235 |
| EI vs MI | Macrophage | 0.050406 | 2.288584e-51 | 6.175172e-57 |
| EI vs MI | Myeloid dendritic cell | 0.254324 | 2.702517e-248 | 0.000000e+00 |
| EI vs SI | B cell | 0.083825 | 1.178991e-96 | 1.557139e-150 |
| EI vs SI | T cell CD4+ | 0.107196 | 3.809351e-144 | 1.339798e-246 |
| EI vs SI | T cell CD8+ | 0.159581 | 7.677489e-195 | 2.067175e-273 |
| EI vs SI | Neutrophil | 0.118693 | 0.000000e+00 | 0.000000e+00 |
| EI vs SI | Macrophage | 0.078002 | 1.256799e-128 | 3.169204e-180 |
| EI vs SI | Myeloid dendritic cell | 0.385459 | 0.000000e+00 | 0.000000e+00 |
| MI vs SI | B cell | 0.026450 | 8.986280e-26 | 3.403545e-44 |
| MI vs SI | T cell CD4+ | 0.035185 | 4.819101e-55 | 4.513072e-76 |
| MI vs SI | T cell CD8+ | 0.048466 | 9.590915e-50 | 5.839289e-70 |
| MI vs SI | Neutrophil | 0.039417 | 2.116086e-108 | 9.448916e-161 |
| MI vs SI | Macrophage | 0.027596 | 2.689146e-36 | 1.517454e-58 |
| MI vs SI | Myeloid dendritic cell | 0.131135 | 5.208450e-151 | 9.751229e-194 |

^*^ Welch’s t-test, ^#^ Mann-Whitney U test

**Supplementary Table S3: P-values of Statistical tests performed between tumor immune-subtype (BLCA) for different immune cells**

| **Comparison** | **Feature** | ∆**-Mean** | **t-test p^*^** | **MW p** ^#^ |
| --- | --- | --- | --- | --- |
| EI vs MI | B cell | 0.046041753 | 0.101204274 | 0.112858451 |
| EI vs MI | T cell CD4+ | 0.050847507 | 0.000107074 | 1.611626e-05 |
| EI vs MI | T cell CD8+ | 0.068254 | 1.451553e-05 | 3.544748e-05 |
| EI vs MI | Neutrophil | 0.082708254 | 8.931472e-20 | 1.742256e-20 |
| EI vs MI | Macrophage | 0.013416904 | 0.388823978 | 0.089206051 |
| EI vs MI | Myeloid dendritic cell | 0.216397974 | 6.056821e-19 | 7.493922e-20 |
| EI vs SI | B cell | 0.074496797 | 0.001984647 | 0.001069467 |
| EI vs SI | T cell CD4+ | 0.092807309 | 1.034148e-12 | 4.827699e-19 |
| EI vs SI | T cell CD8+ | 0.111343231 | 1.204437e-11 | 3.293027e-14 |
| EI vs SI | Neutrophil | 0.131926898 | 1.668106e-36 | 2.372839e-34 |
| EI vs SI | Macrophage | 0.058357263 | 2.818647e-07 | 3.825021e-07 |
| EI vs SI | Myeloid dendritic cell | 0.354715046 | 9.153396e-36 | 5.668306e-35 |
| MI vs SI | B cell | 0.028455044 | 0.070351545 | 0.02296439 |
| MI vs SI | T cell CD4+ | 0.041959803 | 2.737075e-09 | 8.829503e-09 |
| MI vs SI | T cell CD8+ | 0.04308923 | 2.762744e-07 | 4.652718e-09 |
| MI vs SI | Neutrophil | 0.049218645 | 5.107095e-21 | 1.198982e-22 |
| MI vs SI | Macrophage | 0.044940359 | 0.000342428 | 0.000535962 |
| MI vs SI | Myeloid dendritic cell | 0.138317073 | 6.320823e-25 | 9.792553e-26 |

^*^ Welch’s t-test, ^#^ Mann-Whitney U test

**Supplementary Table S4: P-values of Statistical tests performed between tumor immune-subtype (BRCA) for different immune cells**

| **Comparison** | **Feature** | ∆**-Mean** | **t-test p^*^** | **MW p** ^#^ |
| --- | --- | --- | --- | --- |
| EI vs MI | B cell | 0.071960501 | 9.391461e-09 | 1.828967e-08 |
| EI vs MI | T cell CD4+ | 0.137131991 | 4.616865e-20 | 3.608382e-33 |
| EI vs MI | T cell CD8+ | 0.143266979 | 1.373770e-23 | 3.081733e-26 |
| EI vs MI | Neutrophil | 0.137846435 | 1.981895e-46 | 1.858115e-57 |
| EI vs MI | Macrophage | 0.028484907 | 0.002140022 | 0.042503112 |
| EI vs MI | Myeloid dendritic cell | 0.394159906 | 3.824356e-50 | 1.308975e-63 |
| EI vs SI | B cell | 0.087135141 | 4.581927e-12 | 5.192525e-17 |
| EI vs SI | T cell CD4+ | 0.195291923 | 2.007426e-35 | 2.502708e-70 |
| EI vs SI | T cell CD8+ | 0.205746583 | 1.616034e-43 | 3.591987e-59 |
| EI vs SI | Neutrophil | 0.199946217 | 2.807386e-79 | 1.573736e-88 |
| EI vs SI | Macrophage | 0.065804711 | 2.929200e-13 | 1.076450e-18 |
| EI vs SI | Myeloid dendritic cell | 0.594516871 | 2.017673e-87 | 1.865049e-89 |
| MI vs SI | B cell | 0.01517464 | 5.216428e-04 | 2.202010e-04 |
| MI vs SI | T cell CD4+ | 0.058159933 | 1.852238e-31 | 2.749187e-35 |
| MI vs SI | T cell CD8+ | 0.062479604 | 2.987130e-20 | 4.691945e-22 |
| MI vs SI | Neutrophil | 0.062099782 | 5.284068e-34 | 2.152178e-44 |
| MI vs SI | Macrophage | 0.037319804 | 4.917623e-09 | 1.713409e-15 |
| MI vs SI | Myeloid dendritic cell | 0.200356965 | 8.264765e-48 | 2.699519e-52 |

^*^ Welch’s t-test, ^#^ Mann-Whitney U test

**Supplementary Table S5: P-values of Statistical tests performed between tumor immune-subtype (CESC) for different immune cells**

| **Comparison** | **Feature** | ∆**-Mean** | **t-test p^*^** | **MW p** ^#^ |
| --- | --- | --- | --- | --- |
| EI vs MI | B cell | 0.039502276 | 9.247134e-05 | 2.622348e-06 |
| EI vs MI | T cell CD4+ | 0.064752921 | 2.171167e-09 | 9.639506e-12 |
| EI vs MI | T cell CD8+ | 0.0994863 | 1.695993e-06 | 3.669823e-06 |
| EI vs MI | Neutrophil | 0.050999131 | 3.074838e-12 | 7.841623e-12 |
| EI vs MI | Macrophage | 0.027125909 | 0.003101559 | 0.012348812 |
| EI vs MI | Myeloid dendritic cell | 0.161108896 | 9.677926e-15 | 6.699253e-16 |
| EI vs SI | B cell | 0.057570486 | 5.974716e-11 | 6.840973e-11 |
| EI vs SI | T cell CD4+ | 0.084398667 | 9.283696e-15 | 2.244804e-16 |
| EI vs SI | T cell CD8+ | 0.121043098 | 1.215092e-08 | 4.425928e-11 |
| EI vs SI | Neutrophil | 0.082285097 | 4.320555e-23 | 3.467524e-21 |
| EI vs SI | Macrophage | 0.029251425 | 0.001280689 | 0.00654234 |
| EI vs SI | Myeloid dendritic cell | 0.235065377 | 1.709037e-22 | 1.043355e-21 |
| MI vs SI | B cell | 0.01806821 | 0.013527378 | 0.025457817 |
| MI vs SI | T cell CD4+ | 0.019645746 | 0.010231372 | 0.000765006 |
| MI vs SI | T cell CD8+ | 0.021556798 | 0.01931237 | 0.002184251 |
| MI vs SI | Neutrophil | 0.031285966 | 3.190952e-12 | 1.741711e-12 |
| MI vs SI | Macrophage | 0.002125516 | 0.746390972 | 0.879916298 |
| MI vs SI | Myeloid dendritic cell | 0.07395648 | 2.353416e-16 | 2.945881e-15 |

^*^ Welch’s t-test, ^#^ Mann-Whitney U test

**Supplementary Table S6: P-values of Statistical tests performed between tumor immune-subtype (COAD) for different immune cells**

| **Comparison** | **Feature** | ∆**-Mean** | **t-test p^*^** | **MW p** ^#^ |
| --- | --- | --- | --- | --- |
| EI vs MI | B cell | 0.026710808 | 0.012112509 | 0.005545017 |
| EI vs MI | T cell CD4+ | 0.051559857 | 9.066975e-07 | 8.383968e-10 |
| EI vs MI | T cell CD8+ | 0.057515823 | 3.338909e-07 | 1.993277e-07 |
| EI vs MI | Neutrophil | 0.070848796 | 6.457969e-16 | 8.029570e-21 |
| EI vs MI | Macrophage | 0.037569676 | 0.000229899 | 0.0012345 |
| EI vs MI | Myeloid dendritic cell | 0.221267263 | 2.520196e-17 | 7.401620e-23 |
| EI vs SI | B cell | 0.058484787 | 1.041503e-10 | 3.151182e-11 |
| EI vs SI | T cell CD4+ | 0.078730122 | 1.296022e-15 | 6.928578e-21 |
| EI vs SI | T cell CD8+ | 0.089168087 | 2.020833e-14 | 1.819898e-16 |
| EI vs SI | Neutrophil | 0.096542836 | 1.167680e-21 | 1.017101e-31 |
| EI vs SI | Macrophage | 0.062842317 | 6.688347e-10 | 1.155333e-12 |
| EI vs SI | Myeloid dendritic cell | 0.336181968 | 5.687719e-31 | 3.385990e-34 |
| MI vs SI | B cell | 0.031773979 | 2.614544e-05 | 6.178587e-07 |
| MI vs SI | T cell CD4+ | 0.027170265 | 8.610711e-05 | 2.132002e-06 |
| MI vs SI | T cell CD8+ | 0.031652264 | 4.998659e-05 | 4.187433e-06 |
| MI vs SI | Neutrophil | 0.02569404 | 0.000249037 | 4.927944e-14 |
| MI vs SI | Macrophage | 0.025272641 | 3.567193e-07 | 1.590579e-06 |
| MI vs SI | Myeloid dendritic cell | 0.114914705 | 6.347039e-14 | 7.138516e-20 |

^*^ Welch’s t-test, ^#^ Mann-Whitney U test

**Supplementary Table S7: P-values of Statistical tests performed between tumor immune-subtype (ESCA) for different immune cells**

| **Comparison** | **Feature** | ∆**-Mean** | **t-test p^*^** | **MW p** ^#^ |
| --- | --- | --- | --- | --- |
| EI vs MI | B cell | 0.048317129 | 0.01629445 | 0.024663976 |
| EI vs MI | T cell CD4+ | 0.030693417 | 0.001282301 | 0.003627297 |
| EI vs MI | T cell CD8+ | 0.038240739 | 0.003758749 | 0.001671018 |
| EI vs MI | Neutrophil | 0.030927985 | 1.858695e-08 | 6.476831e-08 |
| EI vs MI | Macrophage | 0.015845628 | 0.127178558 | 0.138393284 |
| EI vs MI | Myeloid dendritic cell | 0.104198316 | 8.939444e-06 | 6.799787e-07 |
| EI vs SI | B cell | 0.050348653 | 0.011848892 | 0.004902206 |
| EI vs SI | T cell CD4+ | 0.033618114 | 0.000628451 | 0.000433111 |
| EI vs SI | T cell CD8+ | 0.0440872 | 0.001283932 | 4.060681e-05 |
| EI vs SI | Neutrophil | 0.028926415 | 2.648828e-06 | 1.335415e-07 |
| EI vs SI | Macrophage | 0.019707044 | 0.039167716 | 0.093452276 |
| EI vs SI | Myeloid dendritic cell | 0.14663275 | 6.166452e-09 | 5.219288e-11 |
| MI vs SI | B cell | 0.002031524 | 0.802520088 | 0.787380747 |
| MI vs SI | T cell CD4+ | 0.002924698 | 0.63651811 | 0.220371883 |
| MI vs SI | T cell CD8+ | 0.005846461 | 0.444989574 | 0.220371883 |
| MI vs SI | Neutrophil | -0.00200157 | 0.695594671 | 0.55420102 |
| MI vs SI | Macrophage | 0.003861415 | 0.646523671 | 0.977329271 |
| MI vs SI | Myeloid dendritic cell | 0.042434434 | 0.002272077 | 9.848961e-05 |

^*^ Welch’s t-test, ^#^ Mann-Whitney U test

**Supplementary Table S8: P-values of Statistical tests performed between tumor immune-subtype (GBM) for different immune cells**

| **Comparison** | **Feature** | ∆**-Mean** | **t-test p^*^** | **MW p** ^#^ |
| --- | --- | --- | --- | --- |
| EI vs MI | B cell | 0.035588981 | 0.001001639 | 0.000663117 |
| EI vs MI | T cell CD4+ | 0.018141434 | 0.001937296 | 0.001366019 |
| EI vs MI | T cell CD8+ | 0.003090224 | 0.665880757 | 0.900630136 |
| EI vs MI | Neutrophil | 0.038665145 | 1.600614e-05 | 7.678781e-06 |
| EI vs MI | Macrophage | 0.072223049 | 2.953010e-09 | 4.906541e-10 |
| EI vs MI | Myeloid dendritic cell | 0.084510627 | 7.186984e-12 | 1.579795e-11 |
| EI vs SI | B cell | 0.059489331 | 1.185513e-07 | 1.851944e-07 |
| EI vs SI | T cell CD4+ | 0.039105575 | 2.542056e-11 | 6.701931e-10 |
| EI vs SI | T cell CD8+ | -0.00136 | 0.836209953 | 0.113859493 |
| EI vs SI | Neutrophil | 0.07108127 | 1.868323e-13 | 1.095855e-19 |
| EI vs SI | Macrophage | 0.112686168 | 1.831066e-17 | 4.680812e-19 |
| EI vs SI | Myeloid dendritic cell | 0.126382931 | 1.246962e-20 | 3.917180e-19 |
| MI vs SI | B cell | 0.02390035 | 8.708539e-05 | 3.193796e-05 |
| MI vs SI | T cell CD4+ | 0.020964141 | 1.685192e-05 | 3.402259e-05 |
| MI vs SI | T cell CD8+ | -0.00445 | 0.283339007 | 0.28983509 |
| MI vs SI | Neutrophil | 0.032416125 | 1.648642e-12 | 9.479299e-14 |
| MI vs SI | Macrophage | 0.040463118 | 1.211306e-08 | 3.659524e-10 |
| MI vs SI | Myeloid dendritic cell | 0.041872305 | 7.336993e-07 | 2.636298e-07 |

^*^ Welch’s t-test, ^#^ Mann-Whitney U test

**Supplementary Table S9: P-values of Statistical tests performed between tumor immune-subtype (HNSC) for different immune cells**

| **Comparison** | **Feature** | ∆**-Mean** | **t-test p^*^** | **MW p** ^#^ |
| --- | --- | --- | --- | --- |
| EI vs MI | B cell | 0.042491685 | 0.079586448 | 0.006687079 |
| EI vs MI | T cell CD4+ | 0.057250312 | 0.001280546 | 5.069980e-07 |
| EI vs MI | T cell CD8+ | 0.153010206 | 2.073381e-08 | 3.598532e-11 |
| EI vs MI | Neutrophil | 0.102263075 | 4.826460e-20 | 1.290243e-21 |
| EI vs MI | Macrophage | 0.068278078 | 6.985781e-09 | 3.648003e-09 |
| EI vs MI | Myeloid dendritic cell | 0.30950626 | 2.091157e-22 | 4.329442e-22 |
| EI vs SI | B cell | 0.109789171 | 5.869615e-10 | 5.965865e-11 |
| EI vs SI | T cell CD4+ | 0.094689151 | 8.718678e-11 | 3.414647e-15 |
| EI vs SI | T cell CD8+ | 0.265886986 | 3.545808e-24 | 6.064069e-29 |
| EI vs SI | Neutrophil | 0.16575938 | 1.159602e-39 | 7.076020e-36 |
| EI vs SI | Macrophage | 0.090773449 | 1.415451e-15 | 4.775974e-17 |
| EI vs SI | Myeloid dendritic cell | 0.48623223 | 1.201078e-44 | 1.987898e-35 |
| MI vs SI | B cell | 0.067297485 | 0.000434327 | 4.362066e-06 |
| MI vs SI | T cell CD4+ | 0.037438839 | 0.003853244 | 0.000712749 |
| MI vs SI | T cell CD8+ | 0.11287678 | 2.108497e-08 | 2.053723e-13 |
| MI vs SI | Neutrophil | 0.063496305 | 1.001328e-18 | 5.939428e-21 |
| MI vs SI | Macrophage | 0.022495371 | 0.004014987 | 0.000886793 |
| MI vs SI | Myeloid dendritic cell | 0.17672597 | 5.345643e-15 | 2.041788e-20 |

^*^ Welch’s t-test, ^#^ Mann-Whitney U test

**Supplementary Table S10: P-values of Statistical tests performed between tumor immune-subtype (KIRC) for different immune cells**

| **Comparison** | **Feature** | | ∆**-Mean** | **t-test p^*^** | **MW p** ^#^ |
| --- | --- | --- | --- | --- | --- |
| EI vs MI | B cell | | 0.04401754 | 7.558261e-06 | 1.105315e-05 |
| EI vs MI | | T cell CD4+ | 0.009807208 | 0.436797726 | 0.549009348 |
| EI vs MI | | T cell CD8+ | 0.139238451 | 4.736551e-07 | 1.633901e-08 |
| EI vs MI | | Neutrophil | 0.067431771 | 9.893763e-09 | 2.899373e-08 |
| EI vs MI | | Macrophage | 0.056104343 | 0.001356162 | 8.081813e-05 |
| EI vs MI | | Myeloid dendritic cell | 0.182406706 | 1.335300e-11 | 2.682988e-13 |
| EI vs SI | | B cell | 0.036261272 | 0.000340259 | 0.000226597 |
| EI vs SI | | T cell CD4+ | 0.041644923 | 0.001040077 | 0.000343536 |
| EI vs SI | | T cell CD8+ | 0.165540484 | 1.698569e-08 | 2.869894e-12 |
| EI vs SI | | Neutrophil | 0.103402625 | 1.379102e-13 | 6.360879e-15 |
| EI vs SI | | Macrophage | 0.072229311 | 1.012553e-05 | 3.182388e-08 |
| EI vs SI | | Myeloid dendritic cell | 0.232293298 | 3.509756e-12 | 4.943053e-13 |
| MI vs SI | | B cell | -0.0077562 | 0.364277139 | 0.635403417 |
| MI vs SI | | T cell CD4+ | 0.031837715 | 0.001840146 | 0.000374984 |
| MI vs SI | | T cell CD8+ | 0.026302033 | 0.239836081 | 0.007413032 |
| MI vs SI | | Neutrophil | 0.035970853 | 0.001652521 | 1.400102e-05 |
| MI vs SI | | Macrophage | 0.016124968 | 0.25925596 | 0.046259412 |
| MI vs SI | | Myeloid dendritic cell | 0.049886592 | 0.064614396 | 0.011920982 |

^*^ Welch’s t-test, ^#^ Mann-Whitney U test

**Supplementary Table S11: P-values of Statistical tests performed between tumor immune-subtype (KIRP) for different immune cells**

| **Comparison** | **Feature** | ∆**-Mean** | **t-test p^*^** | **MW p** ^#^ |
| --- | --- | --- | --- | --- |
| EI vs MI | B cell | 0.126376418 | 1.412571e-12 | 2.477677e-15 |
| EI vs MI | T cell CD4+ | 0.020568211 | 0.050588123 | 0.057913193 |
| EI vs MI | T cell CD8+ | 0.122415601 | 0.00030807 | 1.354652e-10 |
| EI vs MI | Neutrophil | 0.046007358 | 5.294191e-11 | 1.852865e-12 |
| EI vs MI | Macrophage | 0.143229346 | 7.572166e-10 | 4.536059e-11 |
| EI vs MI | Myeloid dendritic cell | 0.194362582 | 7.598411e-15 | 3.958511e-17 |
| EI vs SI | B cell | 0.152054223 | 9.796666e-16 | 2.070226e-18 |
| EI vs SI | T cell CD4+ | 0.03383319 | 0.002858896 | 9.868343e-05 |
| EI vs SI | T cell CD8+ | 0.154435617 | 8.100667e-06 | 1.239301e-17 |
| EI vs SI | Neutrophil | 0.059704271 | 7.281642e-16 | 1.377963e-17 |
| EI vs SI | Macrophage | 0.168566852 | 4.115044e-12 | 1.230219e-15 |
| EI vs SI | Myeloid dendritic cell | 0.26081254 | 2.851264e-21 | 4.552295e-21 |
| MI vs SI | B cell | 0.025677805 | 0.005170142 | 1.059806e-06 |
| MI vs SI | T cell CD4+ | 0.013264979 | 0.098195194 | 0.008388227 |
| MI vs SI | T cell CD8+ | 0.032020016 | 0.000541383 | 1.294312e-05 |
| MI vs SI | Neutrophil | 0.013696912 | 0.000467675 | 1.294312e-05 |
| MI vs SI | Macrophage | 0.025337506 | 0.031489495 | 9.896773e-05 |
| MI vs SI | Myeloid dendritic cell | 0.066449958 | 1.702500e-05 | 8.072030e-09 |

^*^ Welch’s t-test, ^#^ Mann-Whitney U test

**Supplementary Table S12: P-values of Statistical tests performed between tumor immune-subtype (LIHC) for different immune cells**

| **Comparison** | **Feature** | ∆**-Mean** | **t-test p^*^** | **MW p** ^#^ |
| --- | --- | --- | --- | --- |
| EI vs MI | B cell | 0.081324028 | 1.377919e-06 | 1.522463e-13 |
| EI vs MI | T cell CD4+ | 0.028308577 | 0.062439869 | 0.00169877 |
| EI vs MI | T cell CD8+ | 0.134734309 | 3.444754e-09 | 1.794521e-13 |
| EI vs MI | Neutrophil | 0.03333419 | 3.942272e-06 | 2.845287e-09 |
| EI vs MI | Macrophage | 0.061803481 | 0.000117703 | 8.293486e-07 |
| EI vs MI | Myeloid dendritic cell | 0.194826914 | 8.652619e-11 | 2.465731e-15 |
| EI vs SI | B cell | 0.11131608 | 7.468125e-11 | 2.387540e-25 |
| EI vs SI | T cell CD4+ | 0.054131892 | 0.000175259 | 2.089582e-09 |
| EI vs SI | T cell CD8+ | 0.168679228 | 6.345784e-13 | 8.037632e-25 |
| EI vs SI | Neutrophil | 0.05615589 | 6.469414e-15 | 1.138560e-25 |
| EI vs SI | Macrophage | 0.091020981 | 1.839543e-08 | 1.049917e-20 |
| EI vs SI | Myeloid dendritic cell | 0.281369611 | 3.110808e-18 | 1.215402e-29 |
| MI vs SI | B cell | 0.029992052 | 9.609915e-07 | 1.164610e-12 |
| MI vs SI | T cell CD4+ | 0.025823315 | 0.001048498 | 0.00151595 |
| MI vs SI | T cell CD8+ | 0.033944919 | 9.460621e-06 | 1.214222e-07 |
| MI vs SI | Neutrophil | 0.022821699 | 1.480655e-08 | 9.113515e-13 |
| MI vs SI | Macrophage | 0.0292175 | 1.932941e-07 | 1.198717e-07 |
| MI vs SI | Myeloid dendritic cell | 0.086542697 | 5.497097e-14 | 1.117447e-14 |

^*^ Welch’s t-test, ^#^ Mann-Whitney U test

**Supplementary Table S13: P-values of Statistical tests performed between tumor immune-subtype (LUAD) for different immune cells**

| **Comparison** | **Feature** | ∆**-Mean** | **t-test p^*^** | **MW p** ^#^ |
| --- | --- | --- | --- | --- |
| EI vs MI | B cell | 0.064337258 | 1.816480e-05 | 2.066634e-05 |
| EI vs MI | T cell CD4+ | 0.09109058 | 6.649627e-11 | 3.598157e-12 |
| EI vs MI | T cell CD8+ | 0.10686371 | 3.916726e-09 | 5.290828e-10 |
| EI vs MI | Neutrophil | 0.096969847 | 1.166626e-14 | 1.673236e-17 |
| EI vs MI | Macrophage | 0.054111436 | 1.605917e-05 | 1.803818e-05 |
| EI vs MI | Myeloid dendritic cell | 0.293630582 | 5.764524e-24 | 2.545985e-25 |
| EI vs SI | B cell | 0.089107048 | 2.211825e-09 | 5.064749e-11 |
| EI vs SI | T cell CD4+ | 0.123503739 | 2.047459e-18 | 2.390166e-20 |
| EI vs SI | T cell CD8+ | 0.128001494 | 8.262080e-12 | 6.299551e-16 |
| EI vs SI | Neutrophil | 0.121310542 | 2.390987e-19 | 1.000676e-23 |
| EI vs SI | Macrophage | 0.065335909 | 2.055701e-07 | 9.296743e-11 |
| EI vs SI | Myeloid dendritic cell | 0.403960697 | 3.093356e-34 | 3.899765e-27 |
| MI vs SI | B cell | 0.024769789 | 0.004882153 | 0.002729911 |
| MI vs SI | T cell CD4+ | 0.032413159 | 0.000612401 | 1.821040e-05 |
| MI vs SI | T cell CD8+ | 0.021137783 | 0.069700388 | 0.002330002 |
| MI vs SI | Neutrophil | 0.024340695 | 0.000991387 | 1.050245e-07 |
| MI vs SI | Macrophage | 0.011224473 | 0.124863648 | 0.000950664 |
| MI vs SI | Myeloid dendritic cell | 0.110330116 | 1.082579e-06 | 4.840073e-11 |

^*^ Welch’s t-test, ^#^ Mann-Whitney U test

**Supplementary Table S14: P-values of Statistical tests performed between tumor immune-subtype (LUSC) for different immune cells**

| **Comparison** | **Feature** | ∆**-Mean** | **t-test p^*^** | **MW p** ^#^ |
| --- | --- | --- | --- | --- |
| EI vs MI | B cell | 0.034591616 | 0.000680106 | 0.000298254 |
| EI vs MI | T cell CD4+ | 0.054676243 | 3.328445e-07 | 1.670504e-06 |
| EI vs MI | T cell CD8+ | 0.119178349 | 4.185767e-15 | 7.720442e-15 |
| EI vs MI | Neutrophil | 0.083883666 | 4.063320e-21 | 1.277501e-22 |
| EI vs MI | Macrophage | 0.061692014 | 3.813765e-09 | 1.289910e-08 |
| EI vs MI | Myeloid dendritic cell | 0.278970613 | 4.497485e-34 | 2.495564e-30 |
| EI vs SI | B cell | 0.067155253 | 1.225390e-12 | 2.942372e-12 |
| EI vs SI | T cell CD4+ | 0.069023952 | 2.146788e-11 | 1.147654e-10 |
| EI vs SI | T cell CD8+ | 0.202651752 | 1.653709e-36 | 3.643260e-34 |
| EI vs SI | Neutrophil | 0.136329931 | 4.980132e-44 | 4.363439e-40 |
| EI vs SI | Macrophage | 0.084991742 | 9.485258e-17 | 3.447466e-18 |
| EI vs SI | Myeloid dendritic cell | 0.440177234 | 2.612464e-60 | 3.139516e-41 |
| MI vs SI | B cell | 0.032563637 | 2.148262e-05 | 3.078313e-05 |
| MI vs SI | T cell CD4+ | 0.014347709 | 0.043816665 | 0.044053636 |
| MI vs SI | T cell CD8+ | 0.083473403 | 1.838921e-16 | 4.758077e-16 |
| MI vs SI | Neutrophil | 0.052446264 | 7.834736e-19 | 1.287837e-18 |
| MI vs SI | Macrophage | 0.023299728 | 3.052955e-05 | 2.058342e-05 |
| MI vs SI | Myeloid dendritic cell | 0.161206621 | 1.012788e-27 | 4.626791e-25 |

^*^ Welch’s t-test, ^#^ Mann-Whitney U test

**Supplementary Table S15: P-values of Statistical tests performed between tumor immune-subtype (PAAD) for different immune cells**

| **Comparison** | **Feature** | ∆**-Mean** | **t-test p^*^** | **MW p** ^#^ |
| --- | --- | --- | --- | --- |
| EI vs MI | B cell | 0.055675737 | 0.00324628 | 0.002376029 |
| EI vs MI | T cell CD4+ | 0.045606895 | 0.010820892 | 0.005114099 |
| EI vs MI | T cell CD8+ | 0.050245778 | 1.230861e-05 | 8.524765e-06 |
| EI vs MI | Neutrophil | 0.013457367 | 0.00588014 | 0.006804591 |
| EI vs MI | Macrophage | 0.035991857 | 0.001230276 | 0.000227965 |
| EI vs MI | Myeloid dendritic cell | 0.115518171 | 1.066935e-06 | 7.027234e-07 |
| EI vs SI | B cell | 0.078232436 | 6.958671e-05 | 2.180261e-07 |
| EI vs SI | T cell CD4+ | 0.057312076 | 0.001695025 | 7.006123e-06 |
| EI vs SI | T cell CD8+ | 0.079438028 | 3.107386e-11 | 1.909734e-11 |
| EI vs SI | Neutrophil | 0.039464845 | 2.249608e-12 | 1.100265e-11 |
| EI vs SI | Macrophage | 0.034582369 | 0.004602807 | 0.000265605 |
| EI vs SI | Myeloid dendritic cell | 0.233440979 | 1.036733e-15 | 6.261221e-14 |
| MI vs SI | B cell | 0.022556698 | 0.000247763 | 0.001610773 |
| MI vs SI | T cell CD4+ | 0.011705181 | 0.016601948 | 0.010954338 |
| MI vs SI | T cell CD8+ | 0.02919225 | 0.000272377 | 0.000409596 |
| MI vs SI | Neutrophil | 0.026007477 | 1.320662e-08 | 2.105121e-08 |
| MI vs SI | Macrophage | -0.001409488 | 0.892492803 | 0.397066499 |
| MI vs SI | Myeloid dendritic cell | 0.117922808 | 6.675299e-14 | 1.664790e-11 |

^*^ Welch’s t-test, ^#^ Mann-Whitney U test

**Supplementary Table S16: P-values of Statistical tests performed between tumor immune-subtype (PRAD) for different immune cells**

| **Comparison** | **Feature** | ∆**-Mean** | **t-test p^*^** | **MW p** ^#^ |
| --- | --- | --- | --- | --- |
| EI vs MI | B cell | 0.066108534 | 3.856257e-11 | 4.729817e-13 |
| EI vs MI | T cell CD4+ | 0.082797706 | 1.170663e-13 | 8.576410e-18 |
| EI vs MI | T cell CD8+ | 0.072447251 | 1.737115e-11 | 1.111437e-10 |
| EI vs MI | Neutrophil | 0.058563994 | 1.084538e-24 | 3.103691e-26 |
| EI vs MI | Macrophage | 0.060305064 | 1.802450e-13 | 7.836331e-13 |
| EI vs MI | Myeloid dendritic cell | 0.191532104 | 7.121984e-31 | 2.927474e-29 |
| EI vs SI | B cell | 0.099035134 | 2.144212e-21 | 6.799889e-29 |
| EI vs SI | T cell CD4+ | 0.128105198 | 2.274343e-26 | 3.884068e-35 |
| EI vs SI | T cell CD8+ | 0.119353398 | 4.698386e-29 | 1.693694e-25 |
| EI vs SI | Neutrophil | 0.090340823 | 1.442667e-43 | 3.018101e-39 |
| EI vs SI | Macrophage | 0.097760698 | 1.481376e-32 | 2.317259e-31 |
| EI vs SI | Myeloid dendritic cell | 0.315948198 | 1.190449e-56 | 3.229488e-40 |
| MI vs SI | B cell | 0.0329266 | 2.647016e-10 | 7.569840e-10 |
| MI vs SI | T cell CD4+ | 0.045307492 | 2.635845e-15 | 6.075314e-15 |
| MI vs SI | T cell CD8+ | 0.046906147 | 4.832649e-07 | 1.829114e-06 |
| MI vs SI | Neutrophil | 0.03177683 | 4.161074e-22 | 1.005202e-21 |
| MI vs SI | Macrophage | 0.037455635 | 7.003999e-11 | 1.105395e-10 |
| MI vs SI | Myeloid dendritic cell | 0.124416094 | 1.281532e-22 | 4.719379e-21 |

^*^ Welch’s t-test, ^#^ Mann-Whitney U test

**Supplementary Table S17: P-values of Statistical tests performed between tumor immune-subtype (SKCM) for different immune cells**

| **Comparison** | **Feature** | ∆**-Mean** | **t-test p^*^** | **MW p** ^#^ |
| --- | --- | --- | --- | --- |
| EI vs MI | B cell | 0.076731827 | 0.000178046 | 1.799576e-06 |
| EI vs MI | T cell CD4+ | 0.133241623 | 6.083260e-10 | 4.764295e-14 |
| EI vs MI | T cell CD8+ | 0.20665908 | 1.118804e-13 | 2.255628e-16 |
| EI vs MI | Neutrophil | 0.094446871 | 7.952649e-23 | 1.463666e-22 |
| EI vs MI | Macrophage | 0.054670894 | 1.566188e-06 | 7.805394e-09 |
| EI vs MI | Myeloid dendritic cell | 0.351633114 | 2.095967e-27 | 2.342469e-31 |
| EI vs SI | B cell | 0.109217017 | 9.054127e-10 | 2.976332e-14 |
| EI vs SI | T cell CD4+ | 0.185889766 | 6.757220e-18 | 1.117927e-29 |
| EI vs SI | T cell CD8+ | 0.255771591 | 8.672188e-19 | 1.282205e-28 |
| EI vs SI | Neutrophil | 0.13365151 | 4.564066e-37 | 2.085514e-36 |
| EI vs SI | Macrophage | 0.08765407 | 9.918984e-17 | 3.876381e-21 |
| EI vs SI | Myeloid dendritic cell | 0.500856849 | 3.257655e-41 | 2.682363e-40 |
| MI vs SI | B cell | 0.03248519 | 0.009307439 | 0.000117084 |
| MI vs SI | T cell CD4+ | 0.052648142 | 2.680713e-07 | 1.618267e-12 |
| MI vs SI | T cell CD8+ | 0.049112511 | 2.496074e-08 | 1.236022e-07 |
| MI vs SI | Neutrophil | 0.039204639 | 1.156391e-13 | 3.777231e-14 |
| MI vs SI | Macrophage | 0.032983177 | 9.938356e-06 | 3.033146e-04 |
| MI vs SI | Myeloid dendritic cell | 0.149223735 | 8.863664e-26 | 3.044018e-28 |

^*^ Welch’s t-test, ^#^ Mann-Whitney U test

**Supplementary Table S18: P-values of Statistical tests performed between tumor immune-subtype (STAD) for different immune cells**

| **Comparison** | **Feature** | ∆**-Mean** | **t-test p^*^** | **MW p** ^#^ |
| --- | --- | --- | --- | --- |
| EI vs MI | B cell | 0.043733543 | 0.012130165 | 0.018362694 |
| EI vs MI | T cell CD4+ | 0.074834905 | 0.000279304 | 2.107534e-06 |
| EI vs MI | T cell CD8+ | 0.078930577 | 1.801525e-05 | 1.291331e-07 |
| EI vs MI | Neutrophil | 0.045706895 | 5.387858e-08 | 3.210199e-09 |
| EI vs MI | Macrophage | 0.050282792 | 0.000317171 | 0.000717398 |
| EI vs MI | Myeloid dendritic cell | 0.21267483 | 3.050977e-14 | 1.814356e-15 |
| EI vs SI | B cell | 0.083942603 | 3.639514e-07 | 3.979872e-08 |
| EI vs SI | T cell CD4+ | 0.110620827 | 7.046243e-08 | 3.077748e-14 |
| EI vs SI | T cell CD8+ | 0.165283583 | 5.345602e-21 | 4.017552e-26 |
| EI vs SI | Neutrophil | 0.088568864 | 2.312902e-23 | 1.286853e-28 |
| EI vs SI | Macrophage | 0.098200169 | 7.840691e-13 | 2.393077e-18 |
| EI vs SI | Myeloid dendritic cell | 0.391403977 | 1.152960e-36 | 9.088175e-33 |
| MI vs SI | B cell | 0.04020906 | 0.001606093 | 4.179047e-06 |
| MI vs SI | T cell CD4+ | 0.035785922 | 0.003681251 | 1.887313e-05 |
| MI vs SI | T cell CD8+ | 0.086353006 | 1.552184e-10 | 7.726234e-13 |
| MI vs SI | Neutrophil | 0.042861969 | 3.664205e-14 | 1.168457e-15 |
| MI vs SI | Macrophage | 0.047917377 | 2.172103e-09 | 4.457313e-11 |
| MI vs SI | Myeloid dendritic cell | 0.178729147 | 8.953467e-18 | 6.592686e-21 |

^*^ Welch’s t-test, ^#^ Mann-Whitney U test

**Supplementary Table S19: P-values of Statistical tests performed between tumor immune-subtype for T-regulatory cells for different cancers**

| **Cancer** | **Comparison** | **∆-Mean** | **t-test p^*^** | **MW p ^#^** |
| --- | --- | --- | --- | --- |
| BLCA | EI vs MI | -0.00227969 | 5.54e-01 | 9.94e-01 |
| BLCA | EI vs SI | -0.01671478 | 1.40e-03 | 1.07e-01 |
| BLCA | MI vs SI | -0.01443509 | 7.36e-03 | 1.24e-01 |
| BRCA | EI vs MI | 0.00999793 | 4.31e-07 | 5.23e-06 |
| BRCA | EI vs SI | 0.00659255 | 1.85e-03 | 3.71e-04 |
| BRCA | MI vs SI | -0.00340538 | 5.15e-02 | 4.65e-01 |
| CESC | EI vs MI | 0.00183292 | 7.23e-01 | 4.51e-01 |
| CESC | EI vs SI | -0.01103284 | 9.59e-02 | 3.86e-01 |
| CESC | MI vs SI | -0.01286576 | 6.30e-02 | 1.44e-01 |
| COAD | EI vs MI | -0.0066649 | 1.10e-01 | 2.86e-01 |
| COAD | EI vs SI | -0.00630576 | 1.05e-01 | 2.96e-01 |
| COAD | MI vs SI | 0.00035914 | 9.33e-01 | 9.34e-01 |
| ESCA | EI vs MI | 0.00724532 | 2.95e-01 | 4.25e-01 |
| ESCA | EI vs SI | -0.00722067 | 3.87e-01 | 6.25e-01 |
| ESCA | MI vs SI | -0.01446599 | 7.29e-02 | 1.57e-01 |
| GBM | EI vs MI | -0.00361217 | 1.33e-01 | 1.24e-01 |
| GBM | EI vs SI | -0.00109555 | 6.96e-01 | 5.51e-01 |
| GBM | MI vs SI | 0.00251662 | 3.89e-01 | 8.06e-02 |
| HNSC | EI vs MI | 0.00030025 | 9.25e-01 | 3.34e-01 |
| HNSC | EI vs SI | 0.00639837 | 3.05e-02 | 5.42e-04 |
| HNSC | MI vs SI | 0.00609812 | 6.96e-02 | 2.28e-02 |
| KIRC | EI vs MI | 0.00070733 | 7.54e-01 | 3.46e-01 |
| KIRC | EI vs SI | -0.00393584 | 1.21e-01 | 5.90e-01 |
| KIRC | MI vs SI | -0.00464316 | 6.89e-02 | 1.58e-01 |
| KIRP | EI vs MI | -0.00490702 | 8.58e-02 | 2.49e-01 |
| KIRP | EI vs SI | -0.00669239 | 3.90e-02 | 4.84e-01 |
| KIRP | MI vs SI | -0.00178536 | 6.17e-01 | 9.26e-01 |
| LIHC | EI vs MI | 0.00057496 | 8.95e-01 | 4.09e-01 |
| LIHC | EI vs SI | 0.00281571 | 5.87e-01 | 2.12e-02 |
| LIHC | MI vs SI | 0.00224075 | 6.81e-01 | 1.09e-01 |
| LUAD | EI vs MI | -0.00032943 | 9.07e-01 | 6.87e-01 |
| LUAD | EI vs SI | 0.0010601 | 6.97e-01 | 1.62e-01 |
| LUAD | MI vs SI | 0.00138953 | 6.17e-01 | 2.45e-01 |
| LUSC | EI vs MI | 0.00464427 | 2.95e-02 | 2.50e-03 |
| LUSC | EI vs SI | 0.00779789 | 1.22e-04 | 9.80e-07 |
| LUSC | MI vs SI | 0.00315362 | 1.03e-01 | 9.18e-02 |
| PAAD | EI vs MI | 0.00239304 | 6.59e-01 | 8.39e-01 |
| PAAD | EI vs SI | -0.00230894 | 7.39e-01 | 7.91e-01 |
| PAAD | MI vs SI | -0.00470198 | 4.89e-01 | 9.88e-01 |
| PRAD | EI vs MI | 0.00172093 | 4.43e-01 | 1.49e-01 |
| PRAD | EI vs SI | 0.00263136 | 2.56e-01 | 1.21e-01 |
| PRAD | MI vs SI | 0.00091043 | 6.81e-01 | 8.93e-01 |
| SKCM | EI vs MI | -0.00105844 | 7.91e-01 | 8.58e-01 |
| SKCM | EI vs SI | 0.00661738 | 9.76e-02 | 5.53e-03 |
| SKCM | MI vs SI | 0.00767582 | 6.12e-02 | 2.11e-03 |
| STAD | EI vs MI | 0.00430975 | 2.91e-01 | 1.72e-01 |
| STAD | EI vs SI | -0.00674363 | 8.13e-02 | 1.05e-01 |
| STAD | MI vs SI | -0.01105339 | 5.39e-03 | 2.46e-03 |

^*^ Welch’s t-test, ^#^ Mann-Whitney U test
